# Protective and Susceptibility Clusters of Environmental Factors, Gene Expression, Antibody Responses, and Cytokines in Pediatric Atopic Dermatitis: Insights from Multi-Modal Data Integration

**DOI:** 10.64898/2026.01.10.26343854

**Authors:** Damir Zhakparov, Nonhlanhla Lunjani, Marco Schmid, Kathleen Moriarty, Damian Roquero, Anita Dreher, Jeannette I. Heldstab-Kast, Kari C. Nadeau, Cezmi Akdis, Michael Levin, Carol Hlela, Milena Sokolowska, Liam O’Mahony, Katja Baerenfaller

## Abstract

**Background:** Atopic dermatitis (AD) is a chronic skin disease that typically occurs in early childhood. In this cross-sectional case-control study, our objective was to employ machine learning approaches to identify novel clusters of protective or susceptibility features associated with AD.

**Methods and Findings:** We utilised an integrated dataset comprising previously established environmental, cytokine, antibody, and gene expression data from AmaXhosa children, both healthy and with AD, living in either rural or urban settings of South Africa, aged 12-36 months. The applied machine learning methods included the GeneSelectR workflow to identify a subset of relevant genes, the calculation of SHAP values to explain the machine learning output, and the use of DIABLO to integrate the datasets for a comprehensive analysis.

Key findings included the identification of a protective cluster of environmental features primarily found in the rural setting, which were correlated with plasma cytokine levels and with expression of autophagy-related genes. Additionally, we identified AD susceptibility clusters where levels of allergen-specific and total IgE antibodies correlated with the cytokines MCP-4 and TARC. Lastly, we identified an RNA-Seq feature signature specific to the disease endotype.

**Conclusions:** The application of various machine learning methods enabled the identification of significant factors associated with AD in a complex, multi-modular dataset, making the output explainable and potentially informing targeted interventions and improved diagnostic criteria.

## 1. Introduction

Atopic dermatitis (AD) is a complex chronic skin disease characterised by profound itching, and eczematous lesions. It is caused by a complex interplay of several factors, including genetic predisposition, microbial dysbiosis, barrier dysfunction, immune dysregulation, and environmental exposures (1). Typically, AD starts in early childhood, and children with AD often develop food allergies, asthma, or allergic rhinitis, a progression known as the atopic march (2,3). AD affects around 22.6% of children worldwide (4). In urban African centres, the prevalence of AD is rapidly increasing, approximating rates observed in North America and Europe, and it is one of the most common skin disorders in this region (5–7). In South Africa, allergic diseases were shown to be less common in rural areas compared to urban settings (8). Due to differences in environmental and genetic backgrounds, the pathogenesis and clinical presentation of AD vary considerably. For example, whilst AD patients from Tanzania have comparable disease severity as Swiss AD patients, they have different IgE sensitization patterns and serological immune signatures (9). Estimating disease prevalence can also be challenging in different countries. For example, the U.K. criteria for atopic eczema were found to perform poorly in Xhosa communities in South Africa, likely due to translation and cultural issues, and therefore cannot be recommended as a primary outcome measure. Instead, the single parameter of visible flexural eczema performed well for diagnosing AD (10). Also, the number of multi-omic AD studies performed in Africa is lower than in other regions, leading to an underrepresentation bias in understanding the complexity of AD pathogenesis. Thus, we sought to identify factors associated with AD in early childhood in the South African population using systems biology approaches. To do this, we used a variety of machine learning (ML) methods to analyse and integrate data previously published by our groups, including clinical questionnaire data on health and living conditions, cytokine and antibody data, and RNA sequencing (RNA-Seq) data of peripheral blood mononuclear cells (PBMCs) from AmaXhosa children in South Africa with and without AD living in urban and rural areas with the aim to identify distinct features associated with AD. The analysis of the individual datasets, and especially their integration, posed several challenges, as the data are multivariate and complex with a dominant environmental variable, and contain a considerably different number of features in the different datasets. For the questionnaire and antibody data, feature selection using ML was followed by calculation of Shapley additive explanation (SHAP) values to interpret the output of the ML model (11). For the RNA-Seq data, the GeneSelectR workflow (12) was employed to generate a gene sub-list with high classification performance and biological relevance. Finally, integrating all four datasets using Data integration Analysis for biomarker discovery using latent components (DIABLO) (13) allowed us to identify clusters of correlated environmental, cytokine, and RNA-Seq features that could be categorised into protective or susceptibility clusters.

## 2. Materials and Methods

### Study Description and Questionnaires

As detailed in (8,14) this study employed a single-visit, cross-sectional case-control design involving AmaXhosa children in South Africa. The children were between 12-36 months old, lived in a rural or urban environment and were healthy or had a dermatologist diagnosis of moderate to severe AD and fulfilled the UK working party criteria of AD. Exclusion criteria included other considerable health conditions, immune-mediated diseases other than allergic comorbidities and positive stool and serum IgE tests for helminths. A standardised questionnaire was used to obtain data on the environmental exposures and living conditions of the children. In total, we obtained clinical data on patient characteristics of 217 AmaXhosa children (rural_AD: 60, rural_healthy control (HC): 52, urban_AD: 56, urban_HC: 49). For 152 children, we additionally had antibody data, for 149 children we had bulk RNA sequencing (RNA-Seq) data, and for 159 children we had plasma cytokine data (S1 Table, S1 Fig.) The study received Human Research Ethics Committee approval (HREC 451/2014) and was conducted in accordance with the declaration of Helsinki (15). Written informed consent was obtained from parents or guardians of all participants prior to inclusion in the study.

### Allergen Sensitization, Allergen Antibody and Cytokine Measurements

As previously reported in (8), skin prick tests were performed using Alk-Abello (Alk-Abello, Denmark) reagents for sensitization to cow’s milk, egg white, peanut, hazelnut, wheat, soy, fish, raw egg white, fresh cow’s milk and fresh peanut. ImmunoCap assays (Thermo Fisher Scientific, Sweden) were used for quantifying the antibody levels of total IgE, specific IgE against *D. pteronyssinus*, milk, fish, wheat, peanut, soy, hazelnut, egg, and CCD, and specific IgG4 against *D. pteronyssinus*, fish, wheat, peanut, soy, hazelnut, casein, and egg.

The levels of plasma analytes and cytokines were assessed using the Meso Scale Discovery multi-spot assay system and QuickPlex SQ120 platform (Meso Scale Discovery, USA).The cytokine data were subjected to statistical analysis testing for differences between HC and AD groups using a Mann-Whitney test, followed by correction for multiple testing with Bonferroni (S2 Table).

### Machine Learning on Questionnaire and Antibody Data

As reported previously, structured questionnaires were used to collect data on family history of atopy and allergy, medical history, household demographics, early-life exposures and various environmental factors (8). The questionnaire features were pre-processed to exclude features mainly consisting of missing values or limited information, as well as those that were highly imbalanced between the two groups. Features directly associated with AD, such as the SCoring Atopic Dermatitis (SCORAD) score or medication with antihistamines, were marked for exclusion from the analyses, while features with more than two possible entries, such as the kind of fuel used for heating or cooking, were one-hot encoded as separate features. The combined pre-processed questionnaire and antibody data were subjected to statistical analysis to identify differences between HC and AD groups. A hypergeometric test was used for categorical variables, and a Mann-Whitney test was used for numerical features. The results were corrected for multiple testing using the Bonferroni method (S3 Table).

In the ML classification, AD vs HC was set as the target variable. All data were included to build the models using cross-validation as the main aim was not to build a single best model, but to extract the features that contributed most to predictive performance. To conduct the hyperparameter search for the models, the *optuna* framework was used, as it enabled the selection of hyperparameter settings that were most likely to yield a high classification score (16). The hyperparameter search space also included selection of the ML algorithms RandomForestClassifier (17,18), Multi Level Perceptron (MLP) (18,19), Gaussian Naive Bayes (GNB) (18,20) or eXtreme Gradient Boosting (XGBOOST) (21). In running the ML pipeline, missing data were initially imputed using the miceforest package (22), which employs RandomForest to estimate the missing values. This approach was selected because of the uneven distribution of missing values between AD and HC, which could otherwise influence the classification. The features were subsequently scaled, and quasi-constant features were removed using a variance threshold of 0.15. To avoid overfitting, the feature selection procedure was constrained to select a minimum of 5 and a maximum of 30 features. The pipeline was then evaluated for every hyperparameter combination in a 5-fold cross-validation (CV) process, taking the mean accuracy as the classification performance measure because the distribution between AD and HC in the data is nearly equal. After the thorough model building process, 78 models were trained and the best model performance was reached in Model 52 (M52). This best-performing Random Forest model had a cross-validation mean accuracy score of 0.81. After obtaining the CV accuracy scores, all models were refitted on the full dataset with their respective parameter settings to generate the final SHAP (SHapley Additive exPlanations) (11) values to interpret the outputs of the ML models. Inspecting the importance of the different hyperparameter search space features revealed that the choice of model had the greatest impact on the objective function, while scaling and imputation contributed little to the classification (S2 Fig.).

### Feature selection in RNA-Seq data

As described previously in (14), total mRNA was isolated from PBMCs and sequenced using HiSeq 40000. The FASTQ sequencing data were preprocessed using the ARMOR pipeline with integrated differential gene expression (DGE) analysis using edgeR (23). From the original 150 RNA-Seq samples one outlier was identified in the Principal Component Analysis and excluded from analyses when rerunning the pipeline. The final RNA-Seq data were from PBMCs of 149 AmaXhosa children living in urban or rural environments and being healthy (HC) or suffering from atopic dermatitis (AD) (groups: urban_HC = 29; urban_AD = 31; rural_HC = 44; rural_AD = 45). These four groups were set as comparisons in the design matrix of edgeR. Transcripts were considered to be significantly changed between two conditions when they had a false discovery rate (FDR) of less than 0.05 and a fold change (FC) greater than 1.5. Transcripts were considered as changing significantly when they had a false discovery rate (FDR) smaller than 0.05 and a fold change (FC) larger than 1.5.

To select relevant genes for subsequent data integration, we utilised the GeneSelectR R workflow that we recently developed (12), designed to overcome shortcomings associated with classical differential gene expression (DEG) analysis in complex datasets. Initially, the input transcript count data were normalised using a between-sample normalisation approach from the edgeR R package (23). Subsequently, the workflow was used to select informative genes (features) based on ML metrics and their biological relevance across several steps:

(1) A standard ML pipeline for feature selection incorporating four ML methods (Logistic Regression with L1 Penalty (Lasso), Random Forest, Boruta and Univariate filtering (Univariate)), with hyperparameter adjustment was applied.
(2) Features were ranked based on their inbuilt method-specific feature importance scores. Additionally, feature ranking across different iterations was reported.
(3) Genes meeting the filtering criterion to appear in at least 2 out of 10 iterations were assembled into a gene list for every method.
(4) A GO enrichment analysis was conducted on each gene list. GO categories with an adjusted p-value < 0.05 were considered enriched and subsequently subjected to semantic similarity analysis (24).

The comprehensive breakdown of the analysis is available in S2 File, and for a more detailed description of the workflow and its usage, please refer to the github repository: https://github.com/dzhakparov/GeneSelectR.

### Weighted Gene Co-expression Network (WGCNA)

The weighted gene co-expression network analysis (WGCNA) was performed using the official R package, as described in (25). In summary, a correlation matrix between genes in the dataset was calculated to detect co-expression patterns among the genes, and genes with similar patterns were clustered into modules. These gene modules were then correlated with the ML-selected questionnaire features using the cor() function.

### Machine Learning on Integrated Data

To integrate the RF560 RNA-Seq, the combined antibody and questionnaire, and the cytokine datasets, the Data Integration Analysis for Biomarker discovery using Latent cOmponents (DIABLO) method from the mixOmics package was used (13).

In brief, DIABLO is a supervised ML method aimed at decomposing the dataset into components projected into latent space and correlated with the label of interest. Using DIABLO, we observed that the datasets had high pairwise correlation scores, suggesting that there is a common pattern across the data modalities (S3 Fig.). This is also reflected in the latent space projection sample plot, where samples are distinctly grouped based on the labels AD or HC (S4 Fig.).

### Data and Code Availability

The R Markdown file with the full analysis breakdown is available in S2 File and on the GitHub repository (link). The FASTQ sequencing data are available in the ArrayExpress database (http://www.ebi.ac.uk/arrayexpress) under accession number E-MTAB-16535.

## 3. Results

### Machine learning applied to questionnaire and antibody data reveals key variables associated with promotion of atopic dermatitis

Statistical analysis of the information in the pre-processed questionnaire data revealed that from the environmental variables the use of electricity or gas as fuel for cooking differed significantly between HC and AD (S3 Table). To go beyond these univariate tests, we employed various machine learning algorithms with HC vs AD as the target variable in a classification process to identify distinguishing features between these groups. By calculating Jaccard scores to assess the similarities between the feature lists selected by the top three models of each ML algorithm, we found that Random Forest achieved the highest feature stability (S5 Fig.). Inspecting the features that were selected in these three best-performing Random Forest models revealed an overlap of 18 features. These features include total IgE antibody levels, as well as the levels of specific IgE and IgG4 antibodies against house dust mites (sIgE_Der and sIgG4_Der) and specific IgE antibodies against different food. Additional features include exposure to food such as eggs, the type of fuel used for heating or cooking, and the duration of sunlight exposure (S4 Table).

To better explain the model output, SHAP values were calculated, which measure how much each of these features contributes to the prediction of the best-performing model. The SHAP values show that the levels of sIgE antibodies against house dust mites (sIgE_Der) and against egg (sIgE_egg), as well as using electricity or gas as fuel for cooking are the three most important features with the greatest influence (S6 Fig.). While feature importance quantifies the contribution of each feature to the model prediction, it does not specify whether high or low feature values correlate with the prediction of AD. To address this, the influence of individual features on predicting AD is illustrated in Figure 1A. The findings reveal that higher levels of specific IgE against house dust mites, egg, fish, milk, and total IgE are associated with the prediction of AD, as shown in the SHAP dependence plots (Figure 1B, S7-S10 Figs.). Similarly, higher levels of specific IgG4 antibodies against house dust mites were positively associated with the prediction of AD (S11 Fig.), as well as no regular exposure to hen egg (Figure 1A). Of the living environment features, using electricity or gas for cooking positively influences the prediction of AD, whereas not using them contributes towards predicting HC. Conversely, using a paraffin stove or fires outside the house for cooking, or wood or coal for heating contributes towards predicting HC (Figure 1A). Finally, a higher age of first exposure to paracetamol is associated with the prediction of HC. These analyses demonstrate that, although individual features in the questionnaire data may not show significant differences in univariate statistical tests, their combined effect contributes to predicting AD.

**Figure 1:**
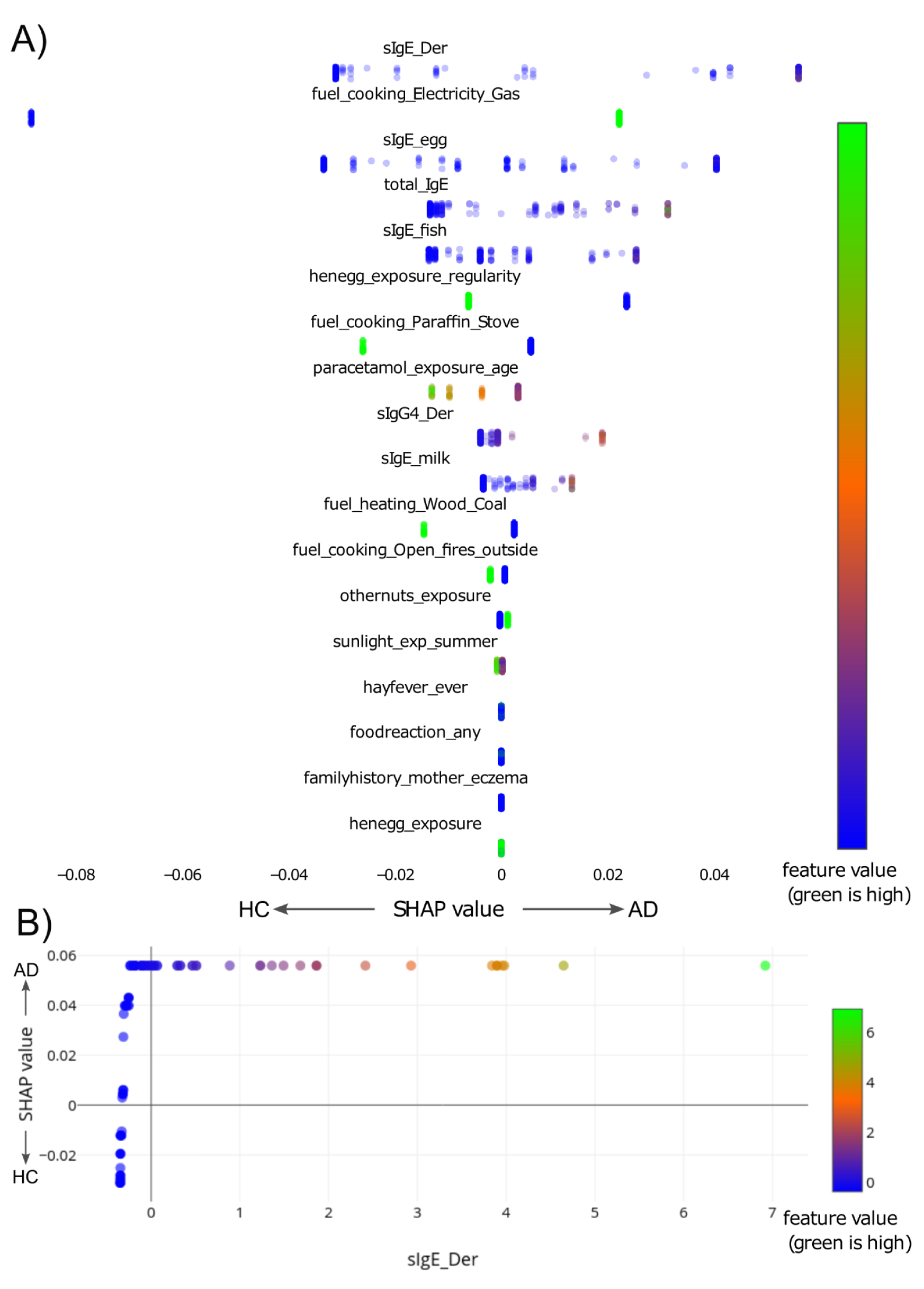
SHAP values of the importance and the impact of the individual features on the output of the best-performing model applied to the combined antibody and questionnaire data. A) The SHAP values of the 18 Random Forest-selected features with the highest impact in target prediction indicate the degree of change towards predicting AD in log odds. Each dot represents a row of data from the questionnaire or antibody data, with the colour of each point indicating the value of the corresponding feature through a colour gradient from green (high value), transitioning through red, to blue (low value). B) SHAP dependence plot for the levels of specific IgE antibodies against house dust mites.

### Adopting a novel ML-based RNA-Seq analysis pipeline revealed a transcript signature of AD

The dataset-wide normalised RNA-Seq data of 149 Xhosa children were first analysed with differential gene expression (DGE) analysis testing for statistical differences between HC and AD, and between rural and urban. As expected from the results of the principal component analysis (S12 Fig.), significantly more transcripts changed between locations than between diagnoses (S5 Table), indicating a strong influence of the environment on the data distribution. For the differences between AD and HC, we found only one gene, Proline Rich Coiled-Coil 2A (PRRC2A), which was present in both the 36 DEGs for the rural samples and the 82 DEGs for the urban samples (S5 Table). In addition, we found no significant correlation in the fold changes between AD and HC for these genes in urban and rural samples. (S13 Fig.). Combined, these findings indicate the limited capability of DGE analysis to identify differential features within a complex dataset where two variables contribute differently to the data distribution.

Considering the univariate nature and limitations of DGE, we used the GeneSelectR workflow (12) to identify a subset of genes distinguishing between AD and HC using ML, as this approach also considers complex nonlinear relationships between features. We employed the four ML-based feature selection methods implemented in GeneSelectR, which are Random Forest (RF), Lasso, Boruta and Univariate Filtering (Univariate). Feature selection based on the inbuilt feature importance resulted in lists with varying numbers of transcripts (RF: 560, Lasso: 372, Univariate: 198, Boruta: 53). The classification performance according to different metrics was similar, with a CV mean score of around 0.7. However, RF scored slightly higher than the other ML methods (S14 Fig.). Calculating three different overlap coefficient metrics implemented in GeneSelectR revealed that there was only minimal overlap and no consensus gene signature shared across the lists (S15 Fig.).

As neither the classification performance metrics nor the overlap coefficients allowed for determining which feature list should be used in downstream analyses, the next step was to evaluate their biological relevance through an overrepresentation analysis of Gene Ontology Biological Process (GOBP) terms. In all the lists, a total of 379 enriched GOBP terms were identified. These functional terms were clustered with binary cut based on their semantic similarity using the topology information from GO. Of the 10 identified clusters, three stood out as more specific to the diagnosis of AD because they were related to immune processes: cluster 1 (lymphocyte development and proliferation), cluster 6 (inflammatory response and oxidative stress), and cluster 7 (metabolism and immune cell processes) (Figure 2A). The 560 features selected with the RF method, termed RF560 list, prominently featured terms from clusters 6 and 7, establishing its significance in AD biomarker identification. In conclusion, the RF-derived list emerged as the most suitable for delineating AD characteristics in the cohort for the subsequent analyses.

**Figure 2.**
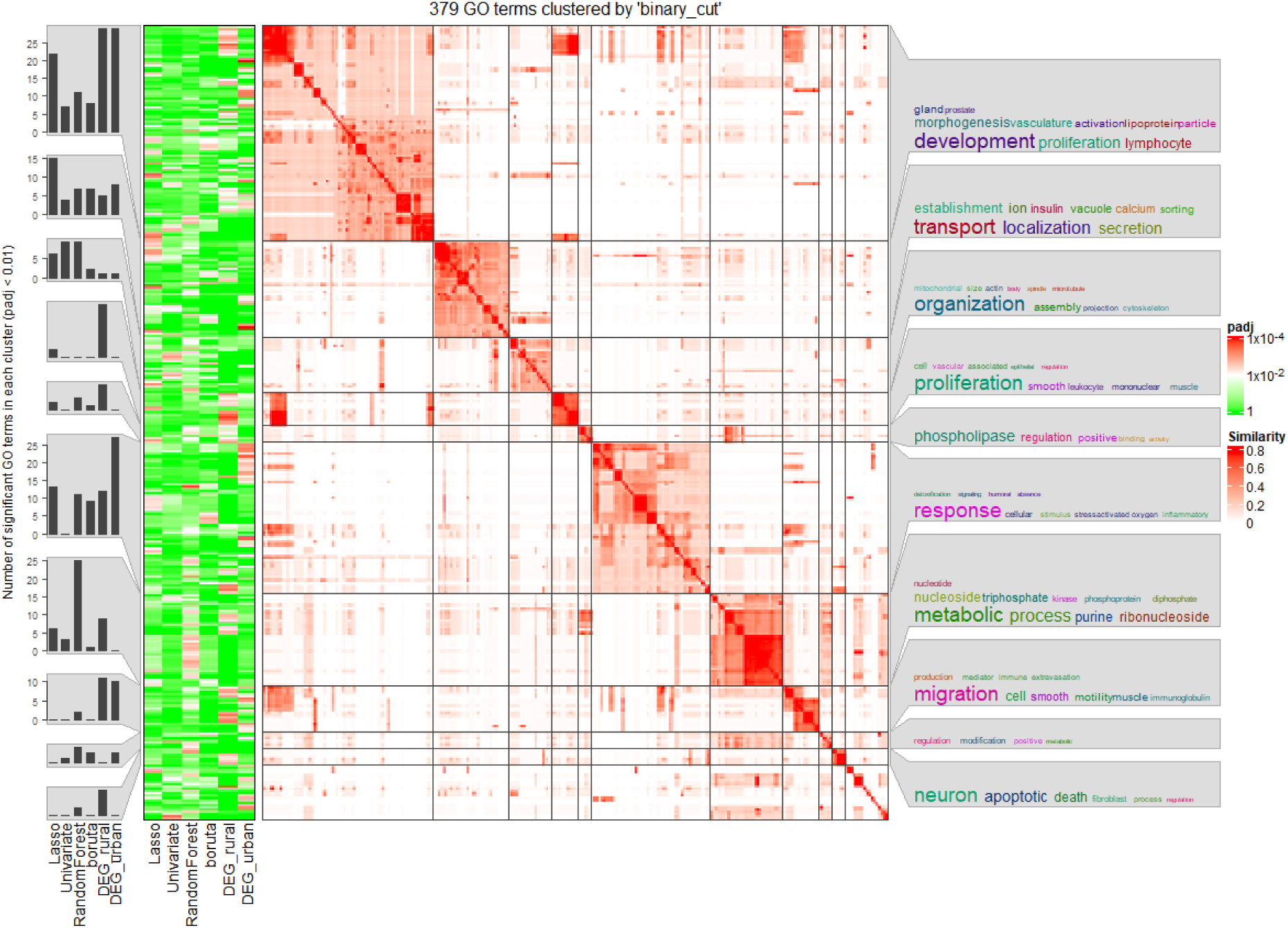
Gene lists associated with AD identified by semantic similarity clustering of enriched GOBP terms. Heatmap of the semantic similarity clustering of the 379 GOBP terms passing an unadjusted p-value threshold of 0.01 in a functional overrepresentation analysis of the feature lists selected by different ML methods and in the DEGs. The composition of each of the 10 clusters is described by the word clouds on the right. At the left, the smaller heatmap shows the significance of the enrichment of the different GOBP terms, with a gradient from red to green, where red indicates a low adjusted p-value, and the histograms demonstrate the number of enriched GOBP terms in each of the clusters.

### Weighted co-expression gene network analysis demonstrates a high correlation of the RF560 list with significant questionnaire and antibody features

The weighted co-expression network analysis (WGCNA) of the genes in the RF560 list revealed 11 clusters of co-expressed genes (S6 Table). These co-expressed gene clusters were correlated with the 18 previously RF-selected questionnaire and antibody features, as well as the two main experimental variables, location (urban vs. rural) and diagnosis (HC vs. AD), to identify significantly associated features. This association can indicate that the gene clusters play a role in influencing these features. Most gene clusters were significantly correlated with location, underscoring the strong influence of this variable on gene expression. In contrast, only the two clusters (the magenta and yellow clusters shown in Figure 3), which were associated with rural location, also showed a significant positive correlation with healthy status. Furthermore, one cluster (the blue cluster) was significantly positively correlated with levels of specific IgE antibodies against milk, one cluster (the black cluster) with any reaction to food, and one cluster (the grey cluster) was significantly negatively correlated with the use of wood or coal as fuel for heating. In summary, the correlation of specific questionnaire and antibody features with specific gene clusters again emphasises the complexity of the dataset.

**Figure 3.**
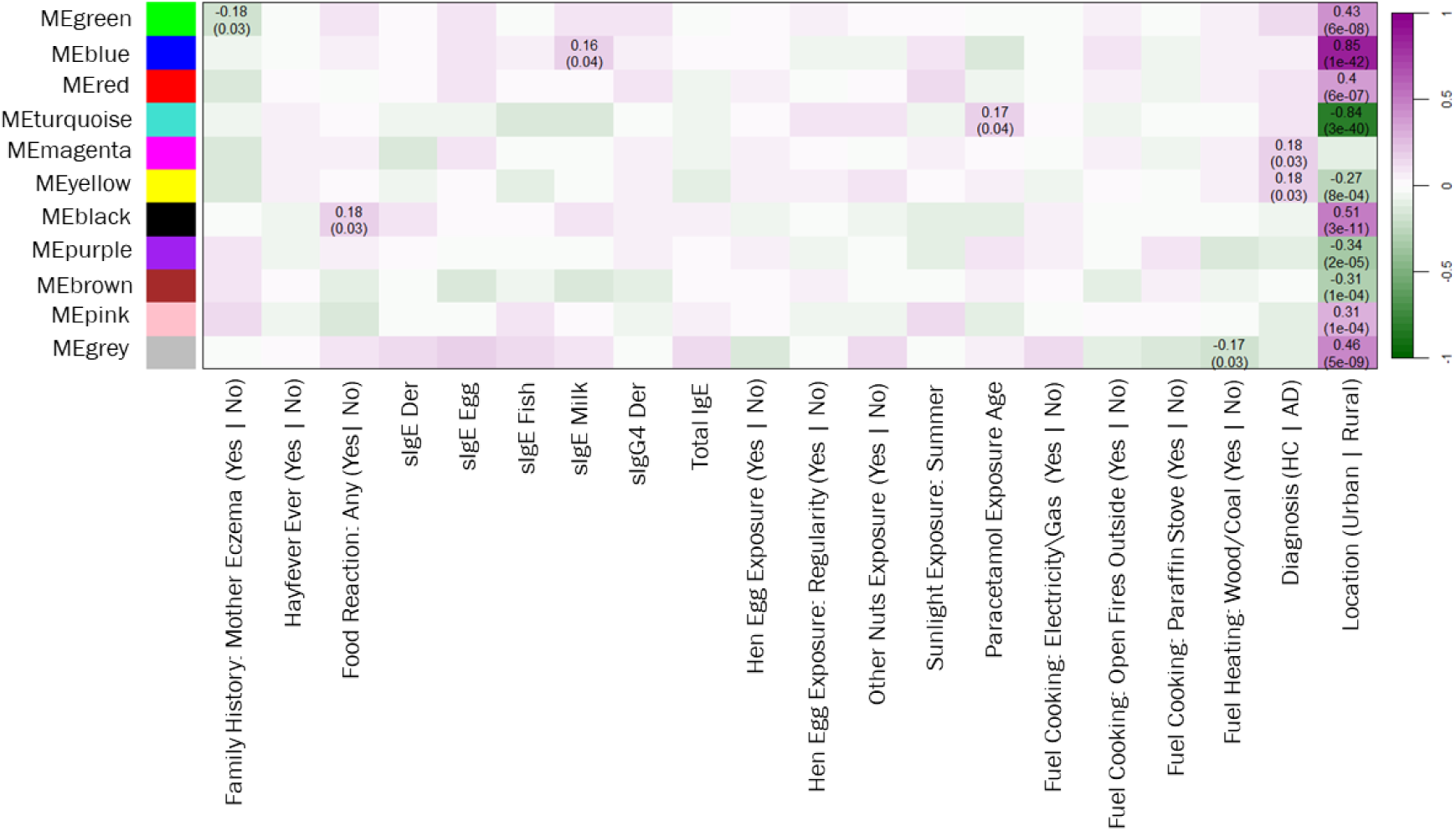
Correlations of co-expressed ML-selected gene clusters with significant questionnaire and antibody features. Heatmap of the correlation between the 11 co-expressed gene clusters identified using WGCNA applied to the RF560 gene list (y-axis) and the 18 questionnaire and antibody features present in the three best RF models plus the major experimental variables location and diagnosis (x-axis). The direction of the correlation is indicated in brackets. For example, Diagnosis (HC | AD) means that the positive correlation of a gene cluster with HC is > 0, while the correlation with AD is < 0. The value of the correlation is indicated with a colour gradient from green to purple, where green indicates a negative correlation and purple indicates a positive correlation. For correlations that passed a p-value threshold of 0.05, the correlation score is indicated along with the p-value in brackets.

### DIABLO integration of all datasets identifies key features related with AD

To further explore features of the four datasets significantly associated with AD, we applied the DIABLO data integration framework (13) (Singh et al., 2019) to investigate relationships among the combined questionnaire and antibody data, the cytokine data, and the RF560-selected transcripts. To determine which samples drive the separation of HC and AD in the latent space projection plot (S4 Fig.), the contributions of variables from the different datasets were visualised on the first two components of the DIABLO model. Since the positive or negative sign of the contribution scores is relevant only in the context of the projection plot, the absolute values of the component contribution scores reflect the importance of each feature (Figure 4).

**Figure 4:**
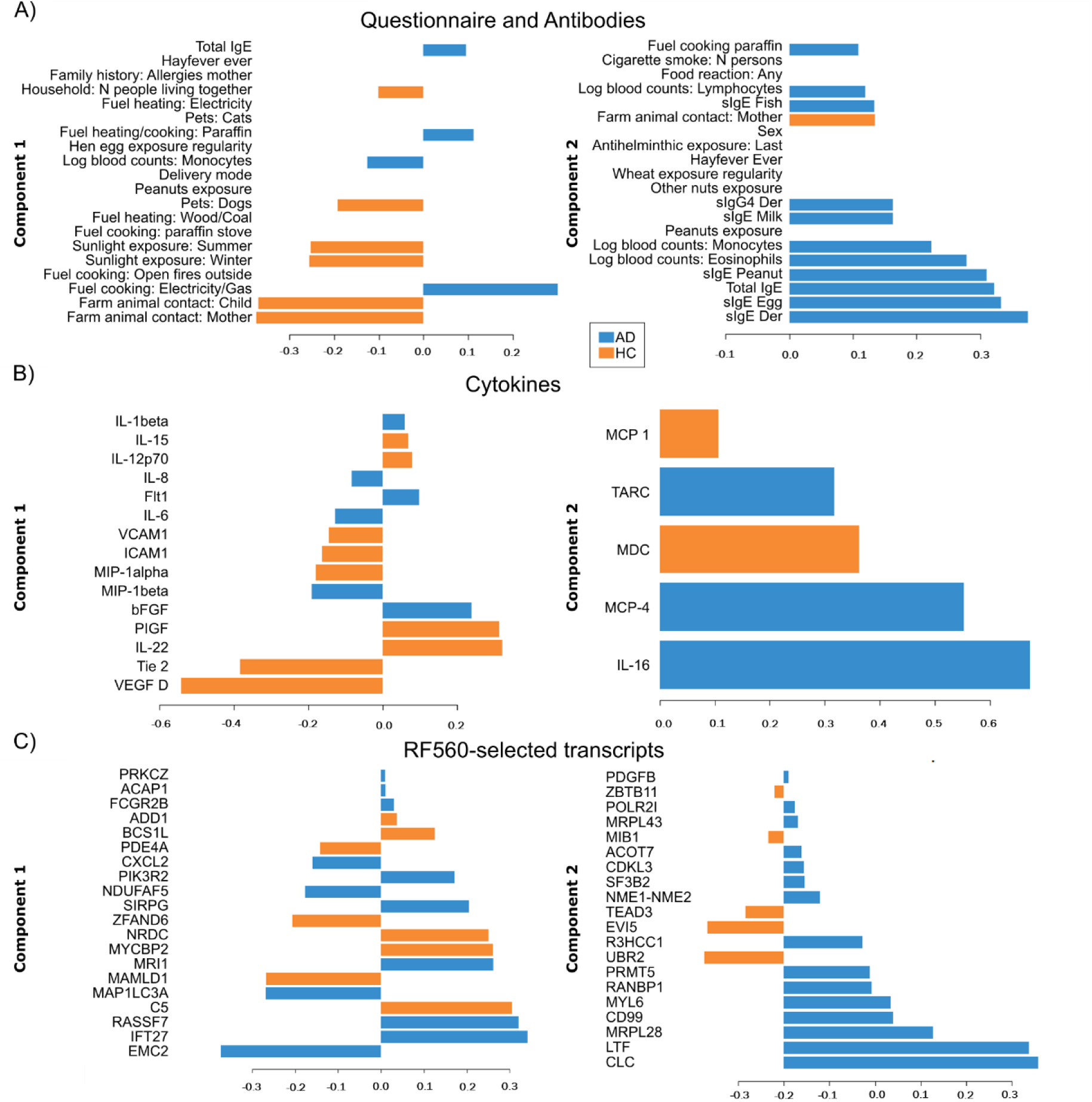
Contributions of variables from the A) combined questionnaire and antibody data, B) the cytokine data, and C) the RF560 selected transcripts to the prediction of AD (blue) or HC (orange) on the first two components of the DIABLO model. The contribution scores of the components are on the x axis, and the feature names are on the y-axis. The higher the absolute value of the contribution score, the more important a feature is. Features without a component score in components 1 or 2 have a tie between the two groups, indicating equal contribution in AD and HC.

In the questionnaire and antibody data, the use of electricity, gas, or paraffin as fuel for cooking or heating, higher counts of monocytes, eosinophils, and lymphocytes, as well as elevated levels of specific and total IgE antibodies contributed to the prediction of AD in the first two components (Figure 4A). The questionnaire features contributing most to the prediction of HC were contact with farm animals of the mother or the child, higher exposure to sunlight in summer and winter, and keeping a dog (Figure 4A). These features are all significantly more prevalent in rural living conditions (8). In the cytokine data, the major contributors to predicting AD in the first two components were IL-1β, IL-8, IL-6, Flt1, MIP-1β, bFGF, TARC, MCP-4, and IL-16 (Figure 1B). The DIABLO model therefore enabled the identification of a greater number of cytokines associated with the diagnosis of AD compared to the univariate statistical test, which identified only Eotaxin-3 and TARC as significantly increased in AD patients compared to HC individuals (S2 Table). Furthermore, DIABLO allowed the identification of transcripts in the RF560 list that contributed most to the prediction of AD or HC. In both components 1 and 2, the major contributors were transcripts predicting AD, including EMC2, IFT27, RASSF7, MAP1LC3A, MRI1, CLC, LTF, MRPL28, CD99, MYL6, RANBP1, and PRMT5. By contrast, the transcripts contributing most to the prediction of HC included C5, MAMLD1, MYCBP2, NRDC, ZFAND6, UBR2, EVI5, and TEAD3 (Figure 4C).

Finally, we used the DIABLO model to investigate how the variables from the four different datasets interact with each other. As depicted in the correlation circle plot in Figure 5A, three distinct clusters of correlated features can be identified, highlighted by the violet, orange, and green areas. The violet cluster (Figure 5B) represents a HC protective cluster. It features the cytokines VGEF D and Tie 2, which contributed most to the prediction of HC in the first component of the DIABLO model for the cytokine data (Figure 4B). These cytokines were correlated with the clinical questionnaire features exposure to sunlight, contact with farm animals, and keeping a dog, which contributed most to the prediction of HC in the first component of the DIABLO model (Figure 4A) and are reported to be protective (26–29). A network analysis of the six correlated features from the RNA-Seq dataset using STRING (30) resulted in the identification of a subcluster comprising ZFAND6 and MAP1LC3A, which was associated with the UniProt keywords autophagy, protein transport and Ubl conjugation pathway (S16 Fig.).

**Figure 5:**
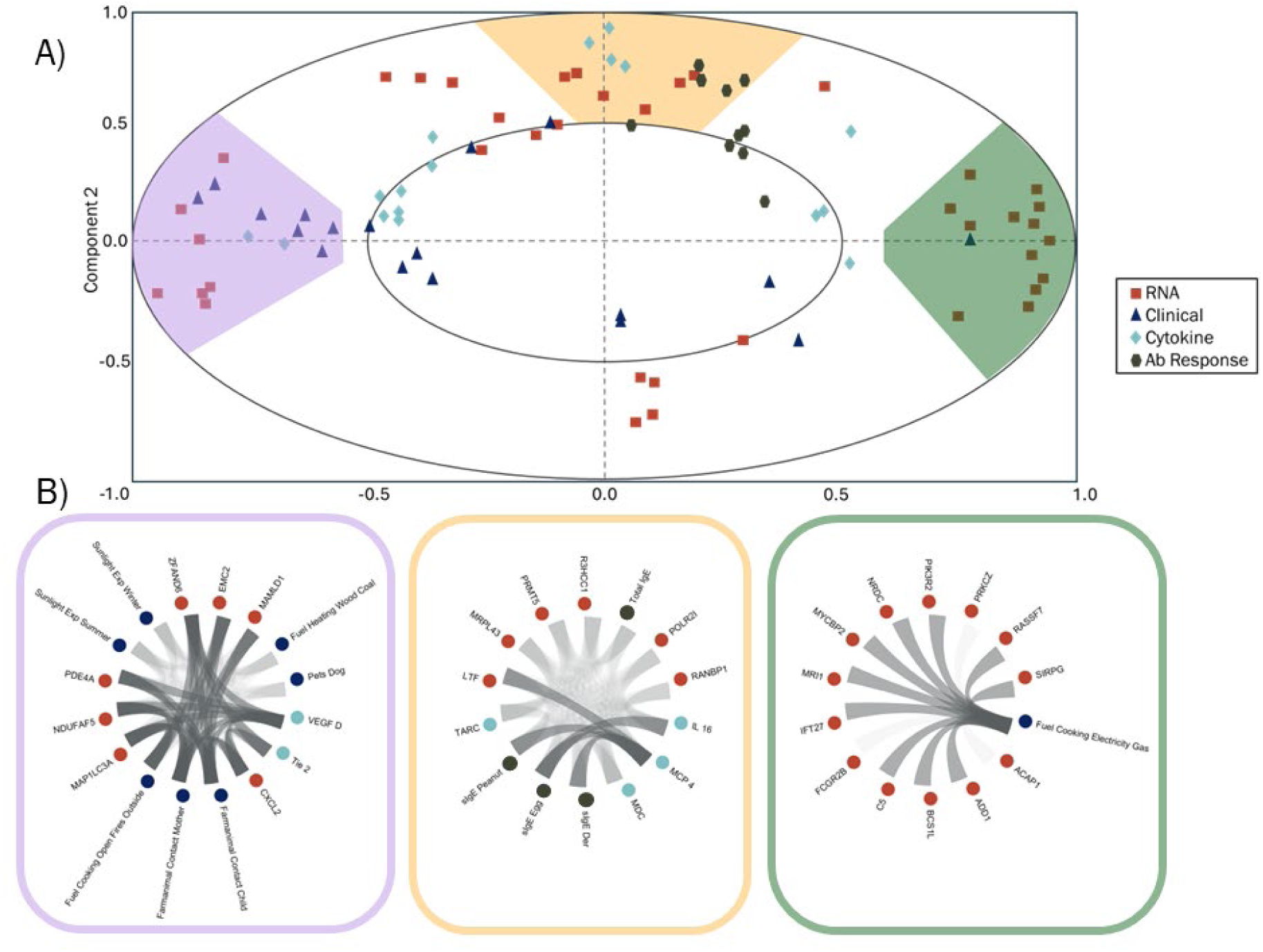
Correlations among features across datasets displayed in a circle plot reveal three distinct clusters. A) Circle plot showing correlations between variables from the RF560-selected transcripts (red squares), the questionnaire (blue triangles), the cytokine (cyan diamonds), and the antibody (black circles) datasets. The angles between two vectors drawn to the variable point from the origin show the correlation score (< 90° = positive correlation, > 90° = negative correlation). The distance from the origin indicates the importance of a variable for the respective component, with the inner circle indicating a correlation score threshold of 0.5. The areas highlighted in violet, orange, or green indicate clusters of correlated variables. B) Circos plots of the correlated features in the violet, orange, or green clusters, respectively.

In contrast, the orange cluster was associated with AD, as it contains the cytokines TARC, MCP4 and IL-16, which are the cytokines contributing to the prediction of AD in the second component of the cytokine DIABLO model (Figure 4B), as well as the levels of total IgE antibodies and of specific IgE antibodies against peanuts, egg and house dust mites (Figure 4A). Submitting the genes of the orange cluster to a network analysis in STRING (30) resulted in three different sub-clusters (S17A Fig.). One of these sub-clusters primarily comprises nuclear proteins involved in transcription and chromatin organisation (S17B Fig.), while another sub-cluster, centred around LTF, shows enrichment for the Reactome pathways *Antimicrobioal peptides* and *Innate immune system* (S17C Fig.).

The green cluster highlighted in the circle plot in Figure 5A was also correlated with AD. It contains the environmental variable to use electricity or gas as fuel for cooking (Figure 5B), which is the variable that contributes most to the prediction of AD in the first component of the DIABLO model of the clinical questionnaire data (Figure 4A). Of the 13 transcript features in the green cluster, the 9 with a fold change of at least 1.08 were considered a signature and subjected to a similarity search against the human mRNASeq dataset in Genevestigator (31) to identify comparisons between conditions or treatments that yield similar transcript signatures. Interestingly, the highest similarity was observed in peripheral blood samples from patients with moderate to severe AD, either untreated or after three months of treatment with Dupilumab, a monoclonal antibody that blocks IL-4 and IL-13 and is used to treat AD. In that study, clinical improvement following Dupilumab administration was accompanied by a decrease in innate immune responses and an increase in B-cell and natural killer cell activation (32). The dataset with the second-highest similarity score was from an asthma study that compared peripheral blood samples collected at baseline and between four and six days after the onset of cold symptoms, without progression to exacerbation (33). These findings underscore the importance of this gene signature in allergic diseases.

## 4. Discussion

The most prominent feature predicting AD in the combined questionnaire and antibody data were the levels of total and allergen-specific IgE antibodies. It is well established that patients with AD often exhibit elevated total and specific IgE levels as a consequence of sensitisation to multiple allergens through a defective epidermal barrier (34). In addition, it was recently shown that skin damage-derived signals alone can be sufficient to initiate humoral immune responses to spatially unlinked antigens (35). Therefore, total and specific IgE are useful biomarkers of sensitisation patterns that may reflect specific AD subtypes associated with atopic comorbidities and could potentially predict the response to targeted therapies, such as treatment with Omalizumab (36–40). This sensitisation pattern might be linked to the eczematous skin lesions that allow easier penetration of allergens (41,42). Damage of the epithelial barrier and the altered skin microbiome in AD (39) might also be associated with the increased expression of LTF, one of the transcripts correlated with IgE antibody levels in a DIABLO susceptibility cluster. LTF is a multifunctional glycoprotein that plays a prominent route in the immune system, and in antimicrobial defence mechanisms (43). The cytokines associated with this susceptibility cluster interestingly comprise TARC and MCP-4, which both contribute to the prediction of AD in component 1 of the clinical questionnaire DIABLO model. Increased levels of TARC and of MCP-4 in the blood of paediatric AD patients has been reported previously (44,45). MCP-4, also known as CCL13, can bind to the receptors CCR1, CCR2, CCR3, CCR5, and CCR11, and is a chemoattractant for monocytes, T cells, immature dendritic cells and eosinophils (46). Corresponding with the increased levels of MCP-4, also the blood monocyte counts were found to be increased in AD and contributing to the prediction of AD in the first component of the clinical questionnaire DIABLO model. The other susceptibility cluster consisted of 13 transcript features and one environmental variable, namely the use of electricity or gas as fuel for cooking. In general, the risk of skin diseases, particularly AD in infants and schoolchildren, increases with higher concentrations of particulate matter (47). Moreover, smoke exposure has been associated with epigenetic modifications at 133 disease-relevant gene loci, increased memory CD8+ T cells, and elevated levels of activation and chemokine receptor biomarkers (48). The identification of this feature in the susceptibility cluster was therefore contrary to expectations, as the use of gas or electricity is generally assumed to generate lower levels of particulate matter than fuels such as wood, coal, or paraffin. However, this finding may reflect underlying differences between the rural cohorts: the rural HC children were from a coastal area, whereas the rural AD children were from a more inland rural region with different lifestyle factors that may predispose individuals to AD. Interestingly, a search of the Genevestigator database (31) for experiments with a similar transcript signature to that found in this susceptibility cluster identified studies investigating AD or asthma. This finding further corroborates the significance and specificity of the identified features for AD.

In contrast, the protective DIABLO cluster includes several environmental exposure variables that are significantly higher in rural areas compared to urban conditions and are protective against AD. These variables include keeping dogs, contact with farm animals, and exposure to sunlight, which may indicate longer periods of time outdoors and exposure to biodiverse environments and increased vitamin D levels (8,14,26–29). The genes in the protective DIABLO cluster that are correlated with these environmental variables comprise ZFAND6, which is a subunit of a TRAF2-cIAP E3 ubiquitin ligase complex and thereby links to the regulation of tumour necrosis factor (TNF)-induced NF-κB signalling (49), and MAP1LC3A that plays a role in the ubiquitin conjugation system and in autophagy, which was shown to influence the pathogenesis of a variety of inflammatory diseases (50). These findings underscore the significant interactions between the immune system and the living environment, which play a crucial role in modulating the risk of atopic sensitisation and AD.

In summary, the use of ML in comprehensively analysing complex datasets including the clinical questionnaire and RNA-Seq datasets, as well as in integrated analysis using DIABLO, allowed for the identification of both known and novel susceptibility and protective factors for AD, as well as unknown interactions between these factors. In analysing the clinical and RNA-Seq data, ML was more efficient in uncovering significant features that differentiate AD from HC, despite the predominant environmental variable. Calculating SHAP values for the questionnaire and antibody data further enhanced the explainability of the ML output. For the RNA-Seq data, identifying the most biologically relevant ML-derived feature list facilitated downstream integration with the questionnaire, antibody and cytokine datasets, which contained considerably fewer features. Correlation analyses ultimately enabled the identification of features contributing to the prediction of HC or AD. This aligns with previous reports showing that multi-omics data integration can uncover more information about implicated molecules and pathways than single-omics approaches (3,51). It also corresponds with the assessment of the 4^th^ Davos Declaration, which emphasises that multidimensional analyses of environmental factors, genetic predisposition, and changes in the skin microbiome are crucial for developing preventive strategies, designing personalised therapies, and understanding geographical variations in AD (1). The strength of ML in analysing complex, multi-modular datasets lies therefore in its ability to reveal intricate patterns and relationships that would otherwise remain undetected.

## Data Availability

The sequencing data are available in the ArrayExpress database under accession number E-MTAB-16535.

http://www.ebi.ac.uk/arrayexpress

## Acknowledgments

This work has been supported by Center for Data Analysis, Visualisation, and Simulation (DAViS), funded by the Swiss Canton of Grisons, and by the Swiss Institute of Allergy and Asthma Research (SIAF). We gratefully acknowledge the SOS-ALL consortium for generating and providing the data used in this study. All data analysed in this work were produced and shared by the consortium.

