## Supporting Information for "Protective and Susceptibility Clusters of Environmental Factors, Gene Expression, Antibody Responses, and Cytokines in Pediatric Atopic Dermatitis: Insights from Multi-Modal Data Integration"

**S1 File. Supporting information file comprising the following supporting tables and figures:**

**S1 Table.** Description of the data.
**S2 Table.** List of the cytokines included in the analyses.
**S3 Table.** List of features in the pre-processed questionnaire data.
**S4 Table.** The occurrence of different features in the top three models of each ML algorithm for the 18 features identified in all three best-performing Random Forest models.
**S5 Table**. Number of significantly changing transcripts in the two comparisons: rural versus urban, and AD versus HC.
**S1 Fig.** Venn diagram showing the overlap of samples with patient information, antibody data, RNA-Seq data, and quantitative cytokine data.
**S2 Fig.** Importance of the hyperparameter space variables model, scaler, number of selected features, imputation using mice, and quasi constant features to the objective function.
**S3 Fig.** Correlation scores between the RNA-Seq, the clinical, and the cytokine datasets.
**S4 Fig.** Projections of samples in the first and second DIABLO components.
**S5 Fig.** Jaccard score for the overlap between the feature lists selected by the best 3 models of each ML algorithm.
**S6 Fig.** Mean SHAP values for the 18 features selected by the Random Forest model M52, applied to the combined questionnaire and antibody data, showing their impact on the target prediction.
**S7 Fig.** SHAP dependence plot for the levels of specific IgE antibodies against hen egg.
**S8 Fig.** SHAP dependence plot for the levels of specific IgE antibodies against fish.
**S9 Fig.** SHAP dependence plot for the levels of specific IgE antibodies against milk.
**S10 Fig.** SHAP dependence plot for the total levels of IgE antibodies.
**S11 Fig.** SHAP dependence plot for the levels of specific IgG4 antibodies against house dust mites.
**S12 Fig.** PCA analysis of the RNA-Seq data with the Principal Components (PCs) 1-3.
**S13 Fig.** Correlation of the fold changes of the DEGs identified in one comparison with the fold changes of these genes in the respective other comparison.
**S14 Fig.** Classification performance metrics of the four different ML methods.
**S15 Fig.** Assessment of the overlap between selected feature lists from the four different ML Methods and the DGE lists using Overlap Coefficient, Jaccard Coefficient, and Sørensen-Dice Coefficient.
**S16 Fig.** Genes in the violet DIABLO cluster subjected to network analysis and k-means subclustering in STRING, highlighting the red subcluster.
**S17 Fig.** Genes in the orange DIABLO cluster subjected to network analysis and k-means subclustering in STRING, highlighting the red and green subclusters.

**S2 File. Markdown document providing an overview of the data analysis carried out on the SOS-ALL project data.**

**Supporting Information S1 File**

**S1 Table. Description of the data**. HC: healthy control, AD: atopic dermatitis

| Feature | samples AD | samples HC | Total |
|---|---|---|---|
| Location rural | 60 | 52 | 112 |
| Location urban | 56 | 49 | 105 |
| **Total questionnaire** | 116 | 101 | **217** |
| Antibody rural | 46 | 39 | 85 |
| Antibody urban | 32 | 35 | 67 |
| **Total Antibody** | 78 | 74 | **152** |
| RNA-Seq rural | 45 | 44 | 89 |
| RNA-Seq urban | 31 | 29 | 60 |
| **Total RNA-Seq** | 76 | 73 | **149** |
| Cytokine rural | 46 | 43 | 89 |
| Cytokine urban | 32 | 38 | 70 |
| **Total Cytokine** | 78 | 81 | **159** |

**S2 Table. List of the cytokines included in the analyses.** The columns 'Values AD' and 'Values HC' indicate the number of non-NA entries and columns 'Distribution AD' and 'Distribution HC' give the median value with the interquartile range for AD and HC, respectively. Statistical significance for the difference between AD and HC was calculated with a Mann-Whitney test, and the resulting p-values were corrected for multiple testing with Bonferroni.

| Cytokine | Values AD | Values HC | Distribution AD | Distribution HC | pvalue | Adjusted pvalue |
|---|---|---|---|---|---|---|
| CRP | 78 | 81 | 4617(1413 - 16096) | 4725 (1560 - 13352) | 0.971 | 1 |
| ICAM.1 | 78 | 81 | 2061 (1693 - 2566) | 2097 (1881 - 2375) | 0.314 | 1 |
| SAA | 78 | 81 | 6545 (1772 - 37614) | 5041 (1579 - 29672) | 0.703 | 1 |
| VCAM.1 | 78 | 81 | 2501 (2122 - 3128) | 2627 (2207 - 3115) | 0.34 | 1 |
| Eotaxin | 78 | 81 | 29.9 (15.4 - 72.1) | 22.3 (13.9 - 59.8) | 0.193 | 1 |
| **Eotaxin.3** | 78 | 80 | 10.0 (6.13 - 25.4) | 4.65 (2.45 - 12.6) | 6.634E-05 | 3.052E-03 |
| IP.10 | 77 | 81 | 74.1 (49.0 - 124) | 64.0 (46.5 - 112) | 0.267 | 1 |
| MCP.1 | 78 | 81 | 35.0 (23.6 - 67.6) | 38.7 (22.9 - 54.0) | 0.56 | 1 |
| MDC | 78 | 81 | 361 (288 - 493) | 355 (295 - 419) | 0.488 | 1 |
| MIP.1alpha | 77 | 81 | 20.4 (8.33 - 52.6) | 17.9 (8.25 - 46.2) | 0.671 | 1 |
| MIP.1beta | 78 | 81 | 75.7 (28.9 - 206) | 45.2 (26.1 - 115) | 5.519E-02 | 1 |
| **TARC** | 78 | 81 | 56.4 (36.8 - 104) | 34.4 (20.7 - 51.8) | 2.336E-06 | 1.075E-04 |
| MCP.4 | 78 | 81 | 44.6 (30.7 - 65.1) | 33.6 (26.3 - 44.6) | 5.202E-03 | 0.239 |
| GM.CSF | 70 | 78 | 0.139 (0.0973 - 0.197) | 0.136 (0.0883 - 0.179) | 0.608 | 1 |
| IL.12.IL.23p40 | 78 | 81 | 71.7 (54.3 - 85.0) | 70.3 (55.7 - 89.9) | 0.44 | 1 |
| IL.15 | 78 | 81 | 0.432 (0.321 - 0.562) | 0.447 (0.343 - 0.580) | 0.486 | 1 |
| IL.16 | 78 | 81 | 74.8 (60.0 - 96.8) | 66.1 (54.4 - 78.0) | 2.480E-02 | 1 |
| IL.17A | 78 | 81 | 3.76 (2.83 - 6.02) | 5.32 (3.36 - 8.49) | 3.220E-03 | 0.148 |
| IL.1alpha | 68 | 69 | 0.250 (0.121 - 0.332) | 0.250 (0.155 - 0.351) | 0.363 | 1 |
| IL.5 | 76 | 79 | 0.349 (0.2209 - 0.552) | 0.334 (0.233 - 0.624) | 0.892 | 1 |
| IL.7 | 78 | 81 | 3.09 (2.57 - 3.86) | 3.05 (2.62 - 3.72) | 0.76 | 1 |
| TNF.beta | 78 | 81 | 0.127 (0.092 - 0.182) | 0.156 (0.102 - 0.212) | 9.270E-02 | 1 |
| VEGF_1 | 78 | 81 | 25.8 (19.8 - 37.6) | 25.1 (15.5 - 37.5) | 0.26 | 1 |
| IFN.gamma | 78 | 81 | 4.97 (2.50 - 11.9) | 3.82 (2.24 - 6.44) | 0.115 | 1 |
| IL.10 | 78 | 81 | 0.370 (0.216 - 0.757) | 0.322 (0.233 - 0.516) | 0.162 | 1 |
| IL.12p70 | 63 | 73 | 0.0638 (0.0355 - 0.160) | 0.0702 (0.0422 - 0.123) | 0.724 | 1 |
| IL.13 | 71 | 71 | 3.23 (1.46 - 6.74) | 2.25 (1.69 - 3.05) | 2.010E-02 | 0.927 |
| IL.1beta | 65 | 69 | 0.169 (0.0442 - 1.13) | 0.0512 (0.0247 - 0.177) | 2.160E-03 | 9.920E-02 |
| IL.2 | 32 | 46 | 0.132 (0.0491 - 4.50) | 0.0986 (0.0596 - 0.197) | 0.366 | 1 |
| IL.4 | 12 | 8 | 2.02 (0.0639 - 3.79) | 0.0304 (0.0106 - 0.0819) | 7.303E-03 | 0.336 |
| IL.6 | 73 | 69 | 2.95 (0.849 - 9.33) | 2.26 (0.869 - 5.932) | 0.321 | 1 |
| IL.8 | 78 | 81 | 32.7 (9.31 - 171) | 24.6 (5.37 - 81.5) | 0.0697 | 1 |
| TNF.alpha | 78 | 80 | 2.10 (1.42 - 3.56) | 1.71 (1.31 - 2.51) | 0.0316 | 1 |
| bFGF | 78 | 81 | 3.58 (1.73 - 32.0) | 3.2 (1.69 - 19.5) | 0.711 | 1 |
| Flt.1 | 78 | 81 | 31.3 (19.2 - 45.1) | 28.4 (19.3 - 47.0) | 0.61 | 1 |
| PlGF | 78 | 81 | 1.42 (0.954 - 32.6) | 1.61 (1.06 - 27.2) | 0.677 | 1 |
| Tie.2 | 77 | 79 | 145 (39.6 - 185) | 146 (25.9 - 182) | 0.743 | 1 |
| VEGF_2 | 78 | 81 | 72.6 (28.3 - 125) | 57.3 (24.1 - 101) | 0.0951 | 1 |
| VEGF.C | 78 | 81 | 35.6 (20.8 - 69.3) | 31.0 (16.6 - 59.9) | 0.318 | 1 |
| VEGF.D | 77 | 79 | 123 (30.1 - 163) | 129 (30.7 - 178) | 0.439 | 1 |
| IL.21 | 18 | 27 | 0.229 (0.141 - 0.346) | 0.307 (0.233 - 0.438) | 0.129 | 1 |
| IL.22 | 78 | 81 | 1.37 (0.723 - 2.21) | 1.66 (0.923 - 2.23) | 0.321 | 1 |
| IL.23 | 31 | 46 | 0.208 (0.148 - 0.533) | 0.270 (0.0918 - 0.435) | 0.655 | 1 |
| IL.27 | 78 | 81 | 285 (230 - 365) | 314 (272 - 419) | 0.0169 | 0.776 |
| IL.31 | 74 | 79 | 0.0684 (0.0472 - 0.1) | 0.0733 (0.0612 - 0.105) | 0.169 | 1 |
| MIP.3alpha | 77 | 81 | 9.87 (6.93 - 14.3) | 9.89 (6.98 - 13.8) | 0.746 | 1 |

**S3 Table**. **List of features in the pre-processed questionnaire data.** The respective data type (num = numerical, cat = categorical) and whether the feature was used in the machine learning (exclude = 0) or not (exclude = 1) are indicated. The columns 'Values AD' and 'Values HC' indicate the number of non-NA entries for numerical features, and the number of positive entries over the number of non-NA entries for categorical features for AD and HC, respectively. Columns 'Distribution AD' and 'Distribution HC' give the median value with the interquartile range for numerical features, and the percentage of positive entries over the number of non-NA entries for categorical features. For categorical features, statistical significance for the difference between AD and HC was calculated with a hypergeometric test, while a Mann-Whitney test was used for numerical features. The resulting p-values were corrected for multiple testing with Bonferroni.

| Feature | Type | Exclude | Values AD | Values HC | Distribution AD | Distribution HC | pvalue | Adjusted pvalue |
|---|---|---|---|---|---|---|---|---|
| enrolment_age | num | 0 | 116 | 101 | 21.5 (16 - 27) | 22 (17 - 29) | 0.44 | 1 |
| gender | cat | 0 | 64 / 115 | 56 / 101 | 55.65% | 55.45% | 0.54 | 1 |
| vaccination_status | cat | 1 | 104 / 114 | 97 / 101 | 91.23% | 96.04% | 0.96 | 1 |
| paracetamol_exposure | cat | 0 | 110 / 114 | 94 / 101 | 96.49% | 93.07% | 0.2 | 1 |
| paracetamol_exposure_age | num | 0 | 109 | 92 | 4 (3 - 6) | 6 (3 - 7.5) | 0.043 | 1 |
| paracetamol_exposure_time | cat | 0 | 74 / 114 | 67 / 100 | 64.91% | 67% | 0.68 | 1 |
| childhood_inf_mumps | cat | 0 | 0 / 114 | 2 / 101 | 0% | 1.98% | 1 | 1 |
| childhood_inf_chickenpox | cat | 0 | 4 / 114 | 7 / 101 | 3.51% | 6.93% | 0.93 | 1 |
| childhood_inf_giandular_fever | cat | 0 | 0 / 114 | 2 / 101 | 0% | 1.98% | 1 | 1 |
| childhood_inf_tuberculosis | cat | 0 | 2 / 114 | 3 / 101 | 1.75% | 2.97% | 0.85 | 1 |
| childhood_inf_other | cat | 0 | 1 / 102 | 2 / 101 | 0.98% | 1.98% | 0.88 | 1 |
| antibiotic_exposure | cat | 0 | 87 / 113 | 75 / 101 | 76.99% | 74.26% | 0.38 | 1 |
| antibiotic_exposure_age | num | 1 | 85 | 75 | 6 (3 - 7) | 6 (3.5 - 8) | 0.28 | 1 |
| antibiotic_exposure_courses | cat | 0 | 24 / 109 | 25 / 99 | 22.02% | 25.25% | 0.76 | 1 |
| antibiotic_exposure_last2months | cat | 0 | 42 / 112 | 33 / 101 | 37.50% | 32.67% | 0.28 | 1 |
| anti_helminthic_exposure | cat | 0 | 95 / 111 | 88 / 101 | 85.59% | 87.13% | 0.7 | 1 |
| anti_helminthic_exposure_age | num | 0 | 94 | 88 | 12 (12 - 12) | 12 (12 - 12) | 0.14 | 1 |
| anti_helminthic_exposure_last | num | 0 | 95 | 88 | 18 (12 - 24) | 18 (12 - 24) | 0.76 | 1 |
| anti_helminthic_exposure_yearly | cat | 0 | 76 / 111 | 75 / 101 | 68.47% | 74.26% | 0.86 | 1 |
| delivery_mode vaginal | cat | 0 | 75 / 114 | 70 / 101 | 65.79% | 69.31% | 0.76 | 1 |
| sunlight_exp_winter | num | 0 | 76 | 62 | 2 (1 - 4) | 3.5 (2 - 5) | 0.0049 | 0.61 |
| sunlight_exp_summer | num | 0 | 73 | 55 | 3 (2 - 6) | 6 (3.5 - 8) | 0.0014 | 0.18 |
| peanuts_mother_exposure_regularity | cat | 0 | 73 / 111 | 55 / 100 | 65.77% | 55% | 0.072 | 1 |
| peanuts_mother_exposure_avoidance | cat | 0 | 12 / 111 | 16 / 99 | 10.81% | 16.16% | 0.91 | 1 |
| breastfeeding_ever | cat | 0 | 92 / 115 | 86 / 101 | 80% | 85.15% | 0.88 | 1 |
| breastfeeding_uptoage_age_complete | num | 0 | 104 | 89 | 3 (0.5 - 6) | 3 (1 - 6) | 0.61 | 1 |
| breastfeeding_uptoage_age_stop | num | 0 | 106 | 92 | 6 (1 - 12) | 6 (2 - 12) | 0.92 | 1 |
| formula_age_introduction | num | 0 | 106 | 99 | 3 (0 - 6) | 3 (0 - 6) | 0.92 | 1 |
| solidfood_age_introduction | num | 0 | 115 | 101 | 5 (4 - 6) | 6 (4 - 6) | 0.46 | 1 |
| solidfood_which_first | cat | 1 | NA | NA | NA | NA | NA | NA |
| peanuts_exposure | cat | 0 | 71 / 114 | 60 / 101 | 62.28% | 59.41% | 0.39 | 1 |
| peanuts_exposure_age_first | num | 1 | 70 | 60 | 12 (10 - 18) | 12 (9 - 19) | 0.84 | 1 |
| peanuts_exposure_regularity | cat | 1 | 60 / 71 | 45 / 61 | 84.51% | 73.77% | 0.095 | 1 |
| othernuts_exposure | cat | 0 | 36 / 114 | 14 / 101 | 31.58% | 13.86% | 0.0016 | 0.2 |
| othernuts_exposure_age_first | num | 1 | 36 | 14 | 12 (11 - 23.5) | 12 (12 - 14) | 0.37 | 1 |
| othernuts_exposure_regularity | cat | 1 | 32 / 36 | Dec-14 | 88.89% | 85.71% | 0.55 | 1 |
| dairyproducts_exposure | cat | 0 | 112 / 114 | 101 / 101 | 98.25% | 100% | 1 | 1 |
| dairyproducts_exposure_age_first | num | 0 | 112 | 101 | 7 (6 - 9.5) | 7 (6 - 9) | 0.61 | 1 |
| dairyproducts_exposure_regularity | cat | 0 | 109 / 112 | 100 / 101 | 97.32% | 99.01% | 0.93 | 1 |
| cowmilkformula_exposure | cat | 0 | 108 / 114 | 97 / 101 | 94.74% | 96.04% | 0.78 | 1 |
| cowmilkformula_exposure_age_first | num | 0 | 99 | 86 | 3 (0 - 6) | 3 (1 - 6) | 0.63 | 1 |
| cowmilkformula_exposure_regularity | cat | 0 | 53 / 107 | 41 / 97 | 49.53% | 42.27% | 0.18 | 1 |
| henegg_exposure | cat | 0 | 94 / 114 | 94 / 101 | 82.46% | 93.07% | 0.1 | 1 |
| henegg_exposure_age_first | num | 0 | 92 | 94 | 12 (8 - 12.5) | 12 (8 - 12) | 0.74 | 1 |
| henegg_exposure_regularity | cat | 0 | 63 / 93 | 88 / 95 | 67.74% | 92.63% | 1 | 1 |

| Feature | Type | Exclude | Values AD | Values HC | Distribution AD | Distribution HC | pvalue | Adjusted pvalue |
|---|---|---|---|---|---|---|---|---|
| wheat_exposure | cat | 0 | 108 / 114 | 100 / 101 | 94.74% | 99.01% | 0.98 | 1 |
| wheat_exposure_age_first | num | 0 | 108 | 100 | 9 (6 - 12) | 10 (6 - 12) | 0.33 | 1 |
| wheat_exposure_regularity | cat | 0 | 99 / 108 | 98 / 100 | 91.67% | 98% | 0.99 | 1 |
| fish_exposure | cat | 0 | 77 / 114 | 80 / 101 | 67.54% | 79.21% | 0.98 | 1 |
| fish_exposure_age_first | num | 1 | 78 | 80 | 12 (12 - 18) | 12 (12 - 17) | 0.83 | 1 |
| fish_exposure_regularity | cat | 1 | 76 / 78 | 77 / 81 | 97.44% | 95.06% | 0.36 | 1 |
| asthma_ever | cat | 0 | 1 / 116 | 1 / 101 | 0.86% | 0.99% | 0.78 | 1 |
| hayfever_ever | cat | 0 | 20 / 116 | 4 / 101 | 17.24% | 3.96% | 0.0014 | 0.17 |
| eczema_ever | cat | 1 | 116 / 116 | 4 / 101 | 100% | 3.96% | 1.21E-57 | 1.49E-55 |
| eczema_age | num | 1 | 116 | 4 | 5 (3 - 7) | 2 (0.75 - 13.5) | 0.28 | 1 |
| eczema_whodiagnosed | cat | 1 | NA | NA | NA | NA | NA | NA |
| medication_any | cat | 1 | 1 / 114 | 1 / 101 | 0.88% | 0.99% | 0.78 | 1 |
| medication_nasalcorticosteroid | cat | 1 | 5 / 114 | 1 / 101 | 4.39% | 0.99% | 0.14 | 1 |
| medication_antihistamines | cat | 1 | 99 / 114 | 14 / 101 | 86.84% | 13.86% | 5.06E-29 | 6.23E-27 |
| medication_steroidcreams | cat | 1 | 109 / 114 | 2 / 101 | 95.61% | 1.98% | 2.90E-52 | 3.57E-50 |
| medication_oral_which | cat | 1 | NA | NA | NA | NA | NA | NA |
| medication_antihistamines_last | cat | 1 | NA | NA | NA | NA | NA | NA |
| medicalhistory_comorbidities | cat | 1 | 1 / 114 | 1 / 101 | 0.88% | 0.99% | 0.78 | 1 |
| household_peoplelivingtogether | num | 0 | 114 | 101 | 5 (4 - 7) | 5 (4 - 8) | 0.13 | 1 |
| household_otherolderchildren | num | 0 | 113 | 101 | 1 (0 - 2) | 1 (0 - 2) | 0.2 | 1 |
| household_otheryoungerchildren | num | 0 | 114 | 101 | 0 (0 - 0) | 0 (0 - 0) | 0.61 | 1 |
| parental_education_level | cat | 1 | NA | NA | NA | NA | NA | NA |
| parental_income | cat | 1 | NA | NA | NA | NA | NA | NA |
| pets_cat | cat | 0 | 18 / 114 | 26 / 101 | 15.79% | 25.74% | 0.975911 | 1 |
| pets_dog | cat | 0 | 50 / 114 | 57 / 101 | 43.86% | 56.44% | 0.976121 | 1 |
| farmanimal_contact_child | cat | 0 | 44 / 113 | 52 / 101 | 38.94% | 51.49% | 0.976238 | 1 |
| farmanimal_contact_mother | cat | 0 | 49 / 113 | 51 / 100 | 43.36% | 51% | 0.894775 | 1 |
| cigarettesmoke_numberofpersons | num | 0 | 111 | 101 | 0 (0 - 0.5) | 0 (0 - 1) | 0.12 | 1 |
| cigarettesmoke_duringpregnancy | cat | 0 | 1 / 112 | 4 / 101 | 0.89% | 3.96% | 0.98 | 1 |
| am_height | num | 1 | 116 | 101 | 81 (77 - 85.5) | 84 (79 - 88) | 0.055 | 0.67 |
| am_weight | num | 1 | 116 | 101 | 11.9 (10.5 - 13.1) | 12.2 (10.8 - 13.6) | 0.34 | 1 |
| am_abd_girth | num | 1 | 113 | 97 | 45 (43 - 48) | 44 (41 - 46) | 0.0057 | 0.7 |
| am_skinfold | num | 1 | 112 | 98 | 11 (9 - 12.5) | 10 (8 - 11) | 2.79E-05 | 0.0034 |
| sp_egg | num | 0 | 87 | 101 | 0 (0 - 0) | 0 (0 - 0) | 1.49E-05 | 0.0018 |
| sp_milk | num | 0 | 109 | 101 | 0 (0 - 0) | 0 (0 - 0) | 0.095 | 1 |
| sp_soy | num | 0 | 109 | 101 | 0 (0 - 0) | 0 (0 - 0) | 0.095 | 1 |
| sp_wheat | num | 0 | 110 | 101 | 0 (0 - 0) | 0 (0 - 0) | 0.018 | 1 |
| sp_fish | num | 0 | 109 | 101 | 0 (0 - 0) | 0 (0 - 0) | NA | NaN |
| sp_peanut | num | 0 | 96 | 101 | 0 (0 - 0) | 0 (0 - 0) | 0.00026 | 0.032 |
| sp_hazelnut | num | 0 | 107 | 101 | 0 (0 - 0) | 0 (0 - 0) | 0.092 | 1 |
| sp_fresh_peanut | num | 0 | 98 | 100 | 0 (0 - 0) | 0 (0 - 0) | 0.0011 | 0.13 |
| sp_fresh_egg | num | 0 | 88 | 101 | 0 (0 - 10) | 0 (0 - 0) | 3.58E-12 | 4.40E-10 |
| sp_fresh_milk | num | 0 | 105 | 101 | 0 (0 - 0) | 0 (0 - 0) | 0.027 | 1 |

| Feature | Type | Exclude | Values AD | Values HC | Distribution AD | Distribution HC | pvalue | Adjusted pvalue |
|---|---|---|---|---|---|---|---|---|
| sIgE_Der | num | 0 | 78 | 74 | 7.9 (0.24 - 31) | 0.086 (0.072 - 0.12) | 8.64E-15 | 1.06E-12 |
| sIgE_milk | num | 0 | 78 | 74 | 0.31 (0.11 - 1.66) | 0.15 (0.051 - 0.30) | 0.0003 | 0.037 |
| sIgE_fish | num | 0 | 78 | 74 | 0.031 (0.022- 0.059) | 0.020 (0.011 - 0.028) | 1.58E-07 | 1.95E-05 |
| sIgE_wheat | num | 0 | 78 | 74 | 0.15 (0.096 - 0.43) | 0.090 (0.080 - 0.11) | 2.02E-09 | 2.49E-07 |
| sIgE_peanut | num | 0 | 78 | 74 | 0.24 (0.073 - 0.83) | 0.061 (0.049 - 0.073) | 1.56E-13 | 1.92E-11 |
| sIgE_soy | num | 0 | 78 | 74 | 0.084 (0.042 - 0.29) | 0.036 (0.027- 0.044) | 8.17E-10 | 1.00E-07 |
| sIgE_hazelnut | num | 0 | 78 | 74 | 0.051 (0.032 - 0.10) | 0.028 (0.022 - 0.034) | 5.82E-10 | 7.15E-08 |
| sIgE_egg | num | 0 | 78 | 74 | 1.02 (0.17 - 4.13) | 0.045 (0.030 - 0.11) | 6.44E-17 | 7.92E-15 |
| sIgE_CCD | num | 0 | 78 | 74 | 0.028 (0.018 - 0.054) | 0.017 (0.011 - 0.027) | 3.40E-06 | 0.00042 |
| sIgG4_Der | num | 0 | 78 | 74 | 17.1 (11.5 - 31.9) | 11.2 (8.08- 15.8) | 5.64E-06 | 0.00069 |
| sIgG4_fish | num | 0 | 78 | 74 | 0.14 (0.001 - 1.03) | 0.001 (0.001 - 1.20) | 0.84 | 1 |
| sIgG4_wheat | num | 0 | 78 | 74 | 16.2 (12.1 - 32.2) | 13.1 (8.98 - 27.3) | 0.02 | 1 |
| sIgG4_peanut | num | 0 | 78 | 74 | 5.79 (3.48 - 14.6) | 4.51 (2.17 - 6.83) | 0.012 | 1 |
| sIgG4_soy | num | 0 | 78 | 74 | 5.36 (3.19 - 12.3) | 4.26 (2.40 - 6.13) | 0.042 | 1 |
| sIgG4_hazelnut | num | 0 | 78 | 74 | 4.30 (3.02 - 6.34) | 3.27 (1.69 - 5.03) | 0.027 | 1 |
| sIgG4_casein | num | 0 | 78 | 74 | 93.7 (51.1 - 339) | 47.6 (23.8 - 182) | 0.0084 | 1 |
| sIgG4_egg | num | 0 | 78 | 74 | 76.7 (18.4 - 508) | 13.40 (5.11 - 407) | 0.0097 | 1 |
| total_IgE | num | 0 | 78 | 74 | 257 (83.9 - 760) | 51.2 (24.7 - 133) | 1.98E-09 | 2.44E-07 |
| scorad | num | 1 | 15 | 0 | 44 (39 - 56) | NA | NA | NA |
| bsa | num | 1 | 113 | 0 | 11.5 (6 - 22.5) | NA | NA | NA |
| stool_katokatz | cat | 0 | NA | NA | NA | NA | NA | NA |
| country | cat | 0 | NA | NA | NA | NA | NA | NA |
| foodreaction_any | cat | 0 | 15 / 114 | 2 / 101 | 13.16% | 1.98% | 0.0018 | 0.22 |
| immun_number | num | 0 | 113 | 100 | 17 (15 - 17) | 17 (15 - 17) | 0.25 | 1 |
| familyhistory_mother_eczema | cat | 0 | 9 / 113 | 1 / 101 | 7.96% | 0.99% | 0.015 | 1 |
| familyhistory_mother_other_allergic_disease | cat | 0 | 20 / 113 | 9 / 101 | 17.70% | 8.91% | 0.046 | 1 |
| familyhistory_father_eczema | cat | 0 | 5 / 112 | 0 / 97 | 4.46% | 0% | 0.042 | 1 |
| familyhistory_father_other_allergic_disease | cat | 0 | 16 / 112 | 3 / 97 | 14.29% | 3.09% | 0.0039 | 0.48 |
| familyhistory_sibling_eczema | cat | 1 | 6 / 45 | 0 / 45 | 13.33% | 0% | 0.013 | 1 |
| familyhistory_sibling_other_allergic_disease | cat | 1 | 6 / 45 | 2 / 45 | 13.33% | 4.44% | 0.13 | 1 |
| fuel_cooking_Electricity_Gas | cat | 0 | 112 / 114 | 60 / 100 | 98.25% | 60% | 1.26E-13 | 1.55E-11 |
| fuel_cooking_Paraffin_Stove | cat | 0 | 6 / 114 | 31 / 100 | 5.26% | 31% | 1 | 1 |
| fuel_cooking_Open_fires_inside | cat | 0 | 2 / 114 | 0 / 100 | 1.75% | 0% | 0.28 | 1 |
| fuel_cooking_Open_fires_outside | cat | 0 | 15 / 114 | 35 / 100 | 13.16% | 35% | 1 | 1 |
| fuel_heating_Electricity | cat | 0 | 25 / 113 | 14 / 101 | 22.12% | 13.86% | 0.082 | 1 |
| fuel_heating_Gas | cat | 0 | 4 / 113 | 3 / 101 | 3.54% | 2.97% | 0.56 | 1 |
| fuel_cooking_heating_Paraffin | cat | 0 | 80 / 113 | 57 / 101 | 70.80% | 56.44% | 0.021 | 1 |
| fuel_heating_Wood_Coal | cat | 0 | 4 / 113 | 25 / 101 | 3.54% | 24.75% | 1 | 1 |
| fuel_heating_Other | cat | 0 | 3 / 113 | 7 / 101 | 2.65% | 6.93% | 0.97 | 1 |
| log_blood_counts_wcc | num | 0 | 100 | 87 | 2.40(2.22 - 2.54) | 2.31(2.13- 2.55) | 0.093 | 1 |
| log_blood_counts_hb | num | 0 | 97 | 82 | 2.40 (2.36- 2.45) | 2.40 (2.33 - 2.46) | 0.89 | 1 |
| log_blood_counts_plts | num | 0 | 96 | 82 | 5.97 (5.77- 6.18) | 5.97 (5.78- 6.16) | 0.55 | 1 |
| log_blood_counts_neutrophils | num | 0 | 94 | 79 | 1.24 (0.97 - 1.65) | 1.26 (0.88 - 1.59) | 0.96 | 1 |
| log_blood_counts_monocytes | num | 0 | 95 | 79 | 0.039 (-0.27 - 0.30) | -0.051(-0.44 - 0.17) | 0.026 | 1 |
| log_blood_counts_lymphocytes | num | 0 | 94 | 79 | 1.76 (1.52- 1.96) | 1.63 (1.42 - 1.91) | 0.08 | 1 |
| log_blood_counts_eosinophils | num | 0 | 3 | 7 | -0.58 (-1.08 - -0.10) | -0.92 (-1.50 - -0.34) | 0.022 | 1 |

**S4 Table. The occurrence of different features in the top three models of each ML algorithm for the 18 features identified in all three best-performing Random Forest models.**

| Feature | Sum | GNB first_3 trial 20 | GNB first_3 trial 28 | GNB first_3 trial 46 | MLP first_3 trial 0 | MLP first_3 trial 11 | MLP first_3 trial 14 | RF first_3 trial 52 | RF first_3 trial 73 | RF first_3 trial 77 | XGBoost first_3 trial 29 | XGBoost first_3 trial 34 | XGBoost first_3 trial 48 |
|---|---|---|---|---|---|---|---|---|---|---|---|---|---|
| fuel_heating_Wood_Coal | 12 | 1 | 1 | 1 | 1 | 1 | 1 | 1 | 1 | 1 | 1 | 1 | 1 |
| total_IgE | 12 | 1 | 1 | 1 | 1 | 1 | 1 | 1 | 1 | 1 | 1 | 1 | 1 |
| fuel_cooking_Electricity_Gas | 12 | 1 | 1 | 1 | 1 | 1 | 1 | 1 | 1 | 1 | 1 | 1 | 1 |
| fuel_cooking_Paraffin_Stove | 12 | 1 | 1 | 1 | 1 | 1 | 1 | 1 | 1 | 1 | 1 | 1 | 1 |
| fuel_cooking_Open_fires_outside | 12 | 1 | 1 | 1 | 1 | 1 | 1 | 1 | 1 | 1 | 1 | 1 | 1 |
| sIgE_Der | 12 | 1 | 1 | 1 | 1 | 1 | 1 | 1 | 1 | 1 | 1 | 1 | 1 |
| sIgE_fish | 12 | 1 | 1 | 1 | 1 | 1 | 1 | 1 | 1 | 1 | 1 | 1 | 1 |
| sIgG4_Der | 12 | 1 | 1 | 1 | 1 | 1 | 1 | 1 | 1 | 1 | 1 | 1 | 1 |
| henegg_exposure_regularity | 12 | 1 | 1 | 1 | 1 | 1 | 1 | 1 | 1 | 1 | 1 | 1 | 1 |
| hayfever_ever | 12 | 1 | 1 | 1 | 1 | 1 | 1 | 1 | 1 | 1 | 1 | 1 | 1 |
| sIgE_milk | 11 | 1 | 1 | 1 | 1 | 1 | 1 | 1 | 1 | 1 | 1 | 1 | 0 |
| paracetamol_exposure_age | 11 | 1 | 1 | 1 | 1 | 1 | 1 | 1 | 1 | 1 | 1 | 1 | 0 |
| othernuts_exposure | 11 | 1 | 1 | 1 | 1 | 1 | 1 | 1 | 1 | 1 | 1 | 1 | 0 |
| foodreaction_any | 11 | 1 | 1 | 1 | 1 | 1 | 1 | 1 | 1 | 1 | 1 | 1 | 0 |
| sunlight_exp_summer | 10 | 1 | 0 | 1 | 1 | 1 | 1 | 1 | 1 | 1 | 1 | 1 | 0 |
| familyhistory_mother_eczema | 10 | 1 | 0 | 1 | 1 | 1 | 1 | 1 | 1 | 1 | 1 | 1 | 0 |
| henegg_exposure | 10 | 1 | 0 | 1 | 1 | 1 | 1 | 1 | 1 | 1 | 1 | 1 | 0 |
| sIgE_egg | 10 | 1 | 0 | 1 | 1 | 1 | 1 | 1 | 1 | 1 | 1 | 1 | 0 |

**S5 Table. Number of significantly changing transcripts in the two comparisons: rural versus urban, and AD versus HC.** Number of transcripts with FDR < 0.05 and abs(fold change) > 1.5 in the respective comparison; HC: healthy control, AD: atopic dermatitis

| | Higher in rural | Higher in urban | Total |
|---|---|---|---|
| Rural-AD vs Urban-AD | 2293 | 2406 | 4699 |
| Rural-HC vs Urban-HC | 1898 | 1933 | 3831 |
| Overlap | 1568 | 1571 | 3139 |
| | Higher in AD | Higher in HC | |
| Rural-AD vs Rural-HC | 9 | 27 | 36 |
| Urban-AD vs Urban-HC | 54 | 28 | 82 |
| Overlap | 1 | 0 | 1 |

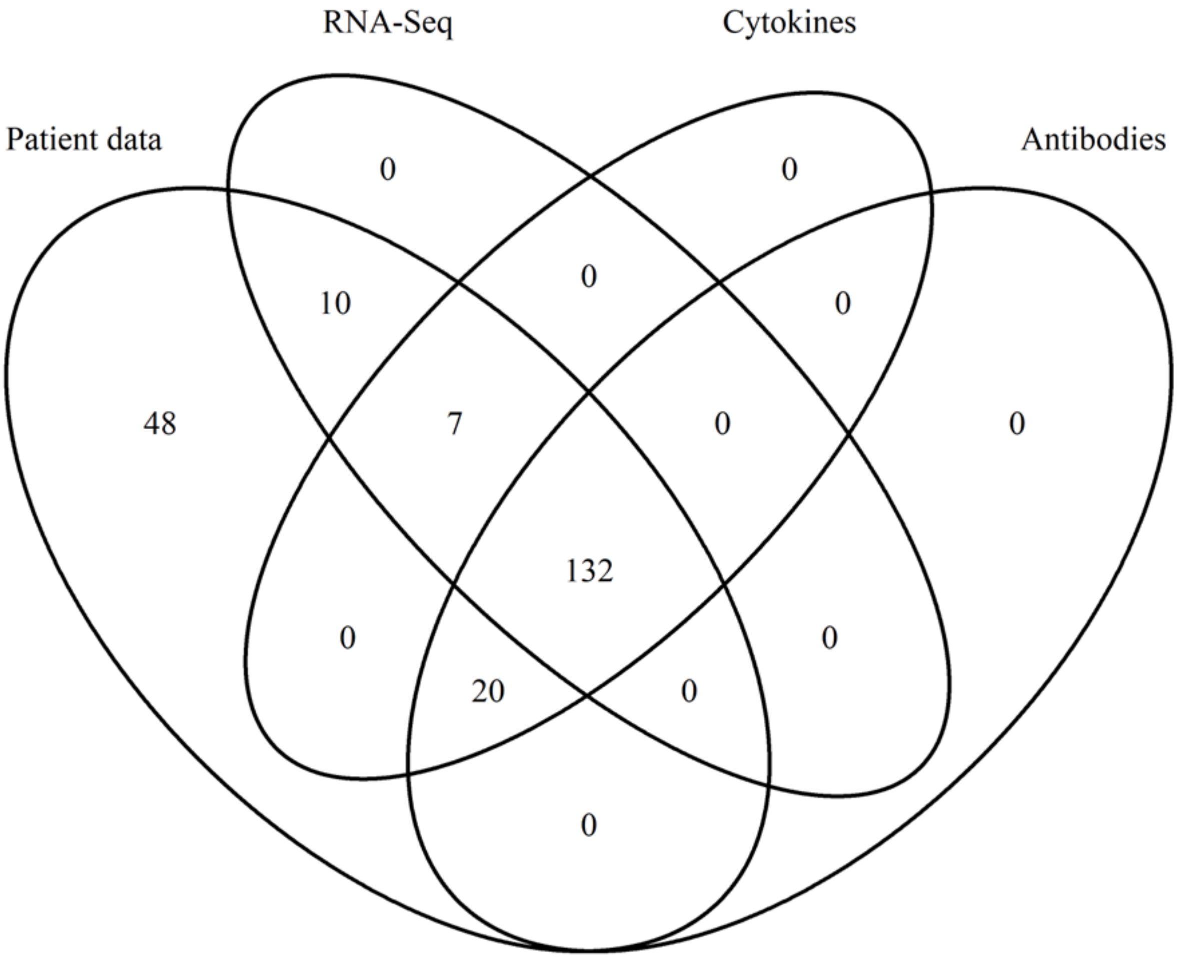


**S1 Fig.** Venn diagram showing the overlap of samples with patient information, antibody data, RNA-Seq data, and quantitative cytokine data.

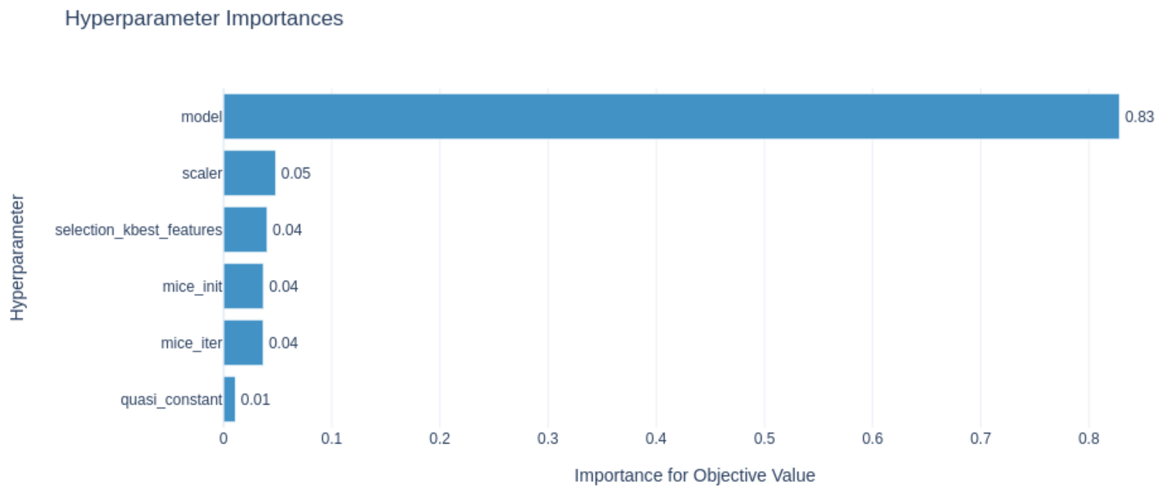


**S2 Fig.** Importance of the hyperparameter space variables model, scaler, number of selected features, imputation using mice, and quasi constant features to the objective function.

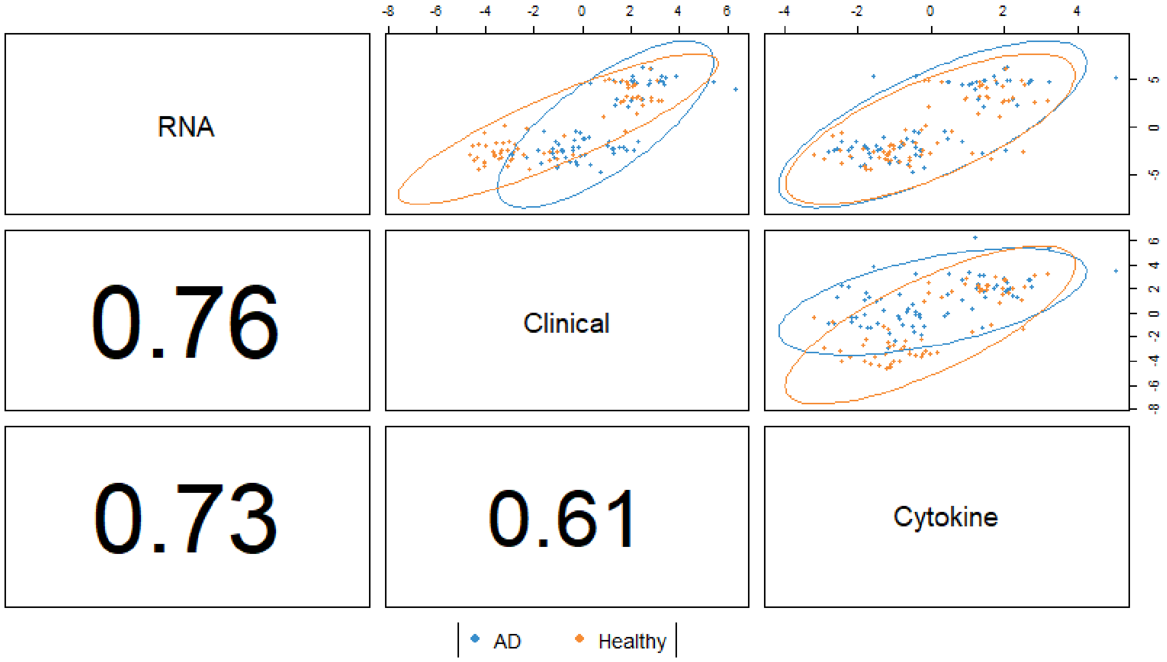


**S3 Fig.** Correlation scores between the RNA-Seq, the clinical, and the cytokine datasets (HC samples in orange, AD samples in blue). The clinical dataset corresponds to the combined questionnaire and antibody data.

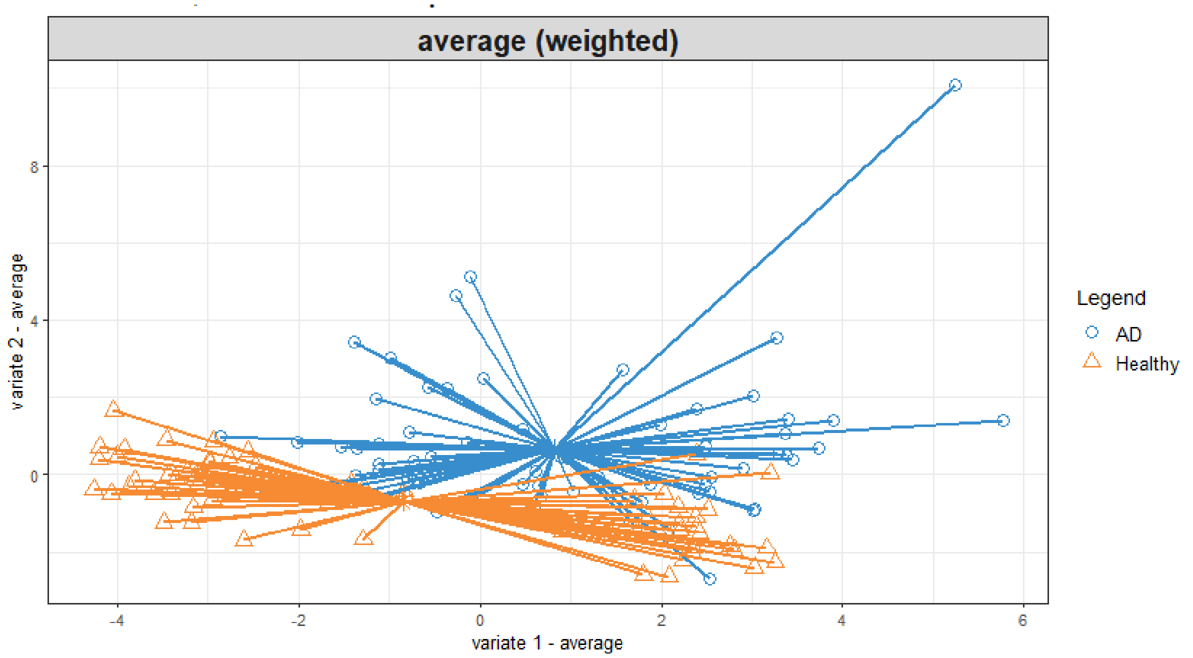


**S4 Fig.** Projections of samples in the first and second DIABLO components (HC samples in orange, AD samples in blue).

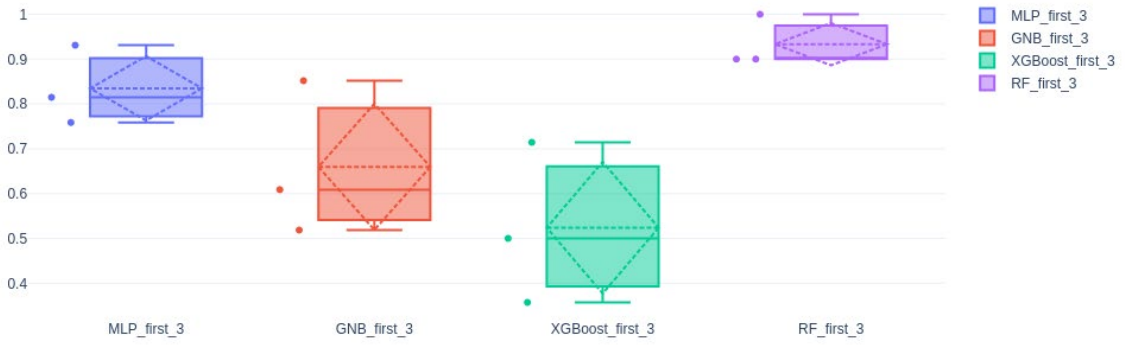


**S5 Fig.** Jaccard score for the overlap between the feature lists selected by the best 3 models of each ML algorithm.

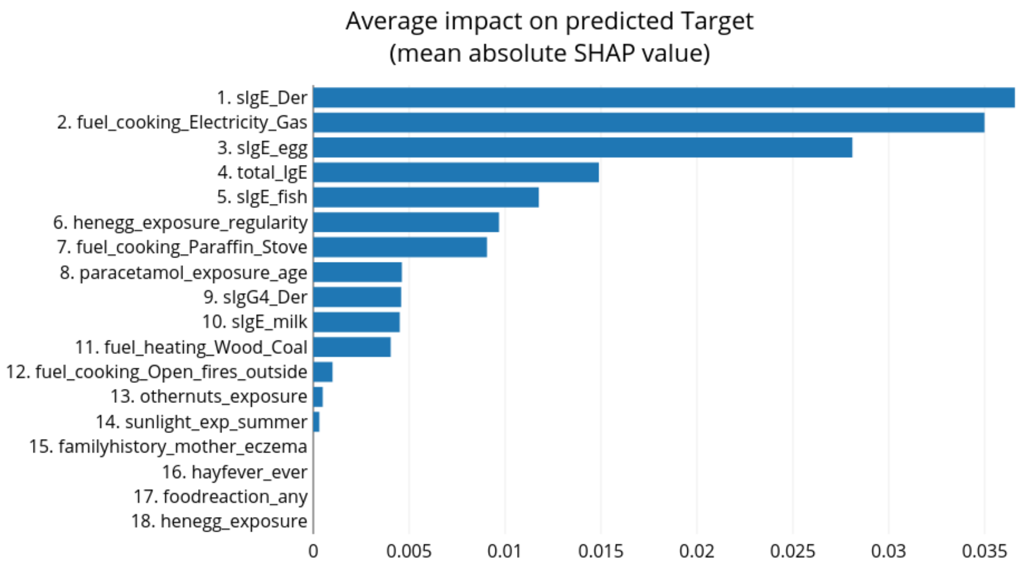


**S6 Fig.** Mean SHAP values for the 18 features selected by the Random Forest model M52, applied to the combined questionnaire and antibody data, showing their impact on the target prediction.

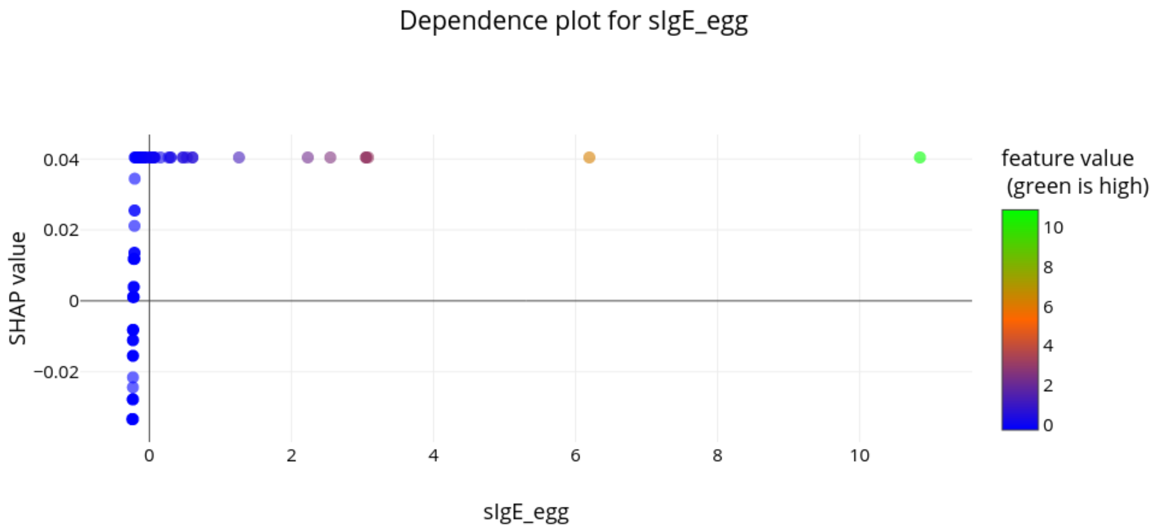


**S7 Fig.** SHAP dependence plot for the levels of specific IgE antibodies against hen egg.

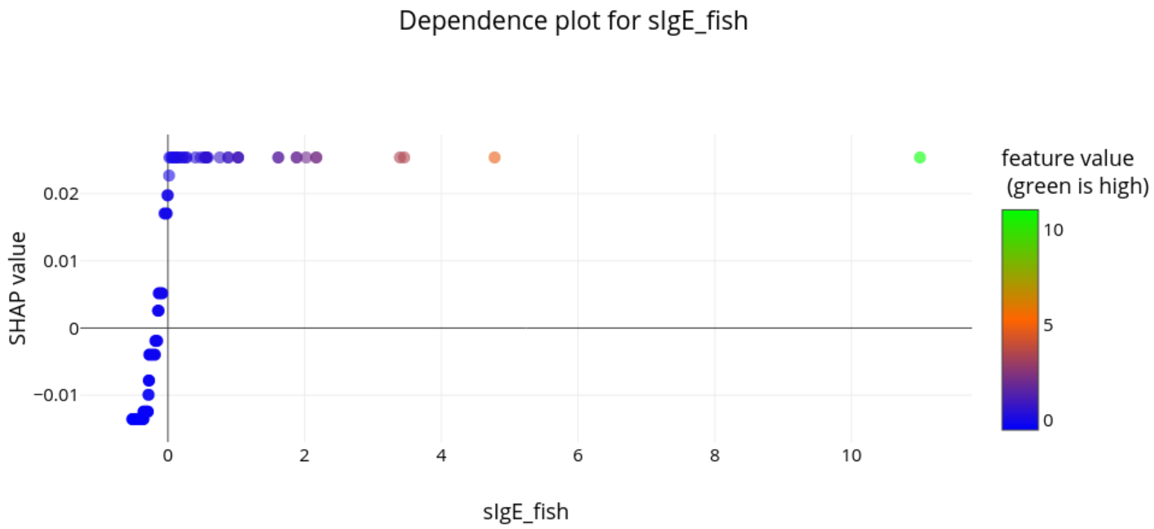


**S8 Fig.** SHAP dependence plot for the levels of specific IgE antibodies against fish.

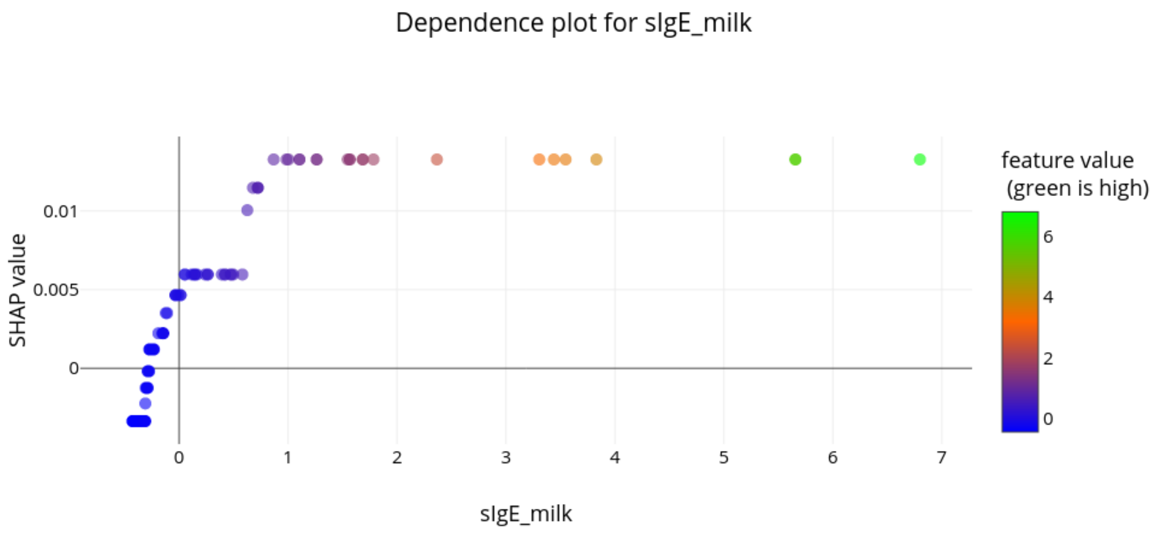


**S9 Fig.** SHAP dependence plot for the levels of specific IgE antibodies against milk.

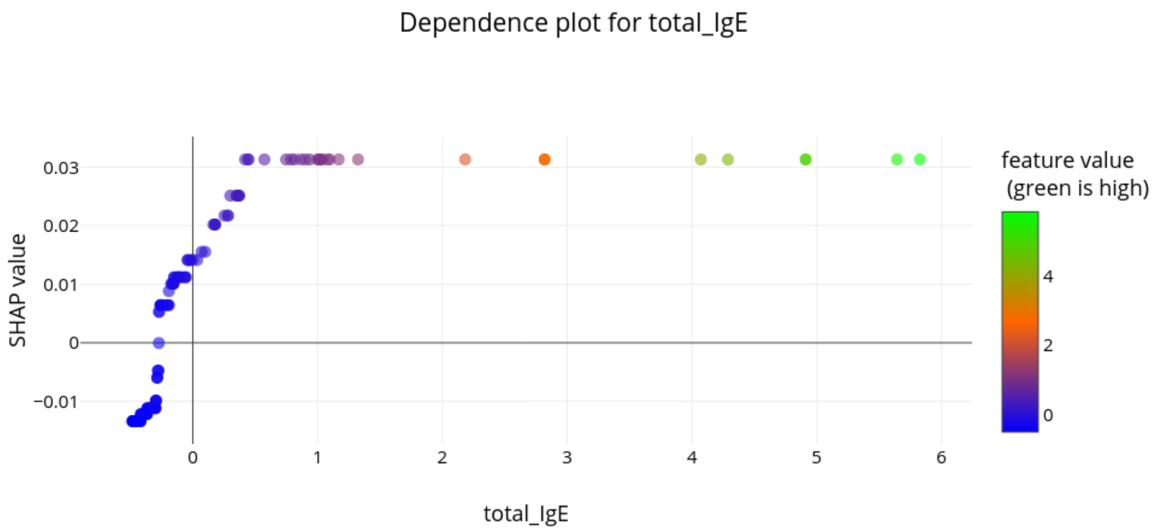


**S10 Fig.** SHAP dependence plot for the total levels of IgE antibodies.

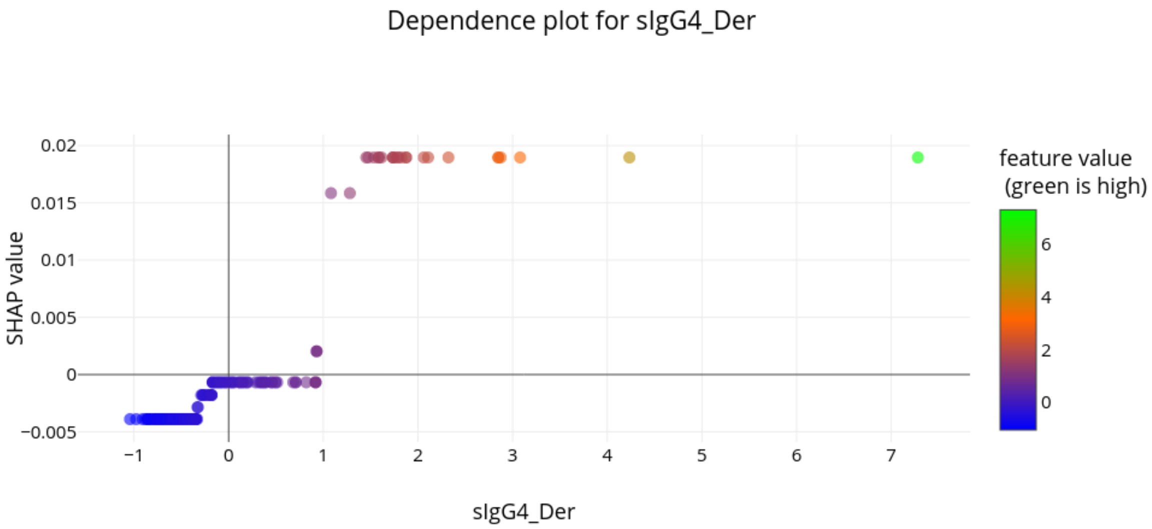


**S11 Fig.** SHAP dependence plot for the levels of specific IgG4 antibodies against house dust mites.

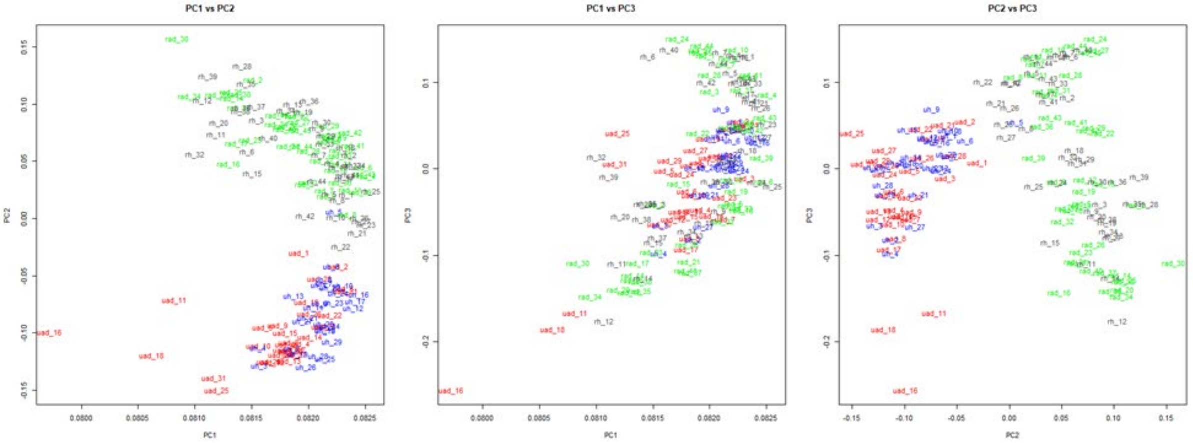


**S12 Fig.** PCA analysis of the RNA-Seq data with the Principal Components (PCs) 1-3 (green: rural – AD; grey: rural – HC; red: urban – AD; blue: urban – HC)

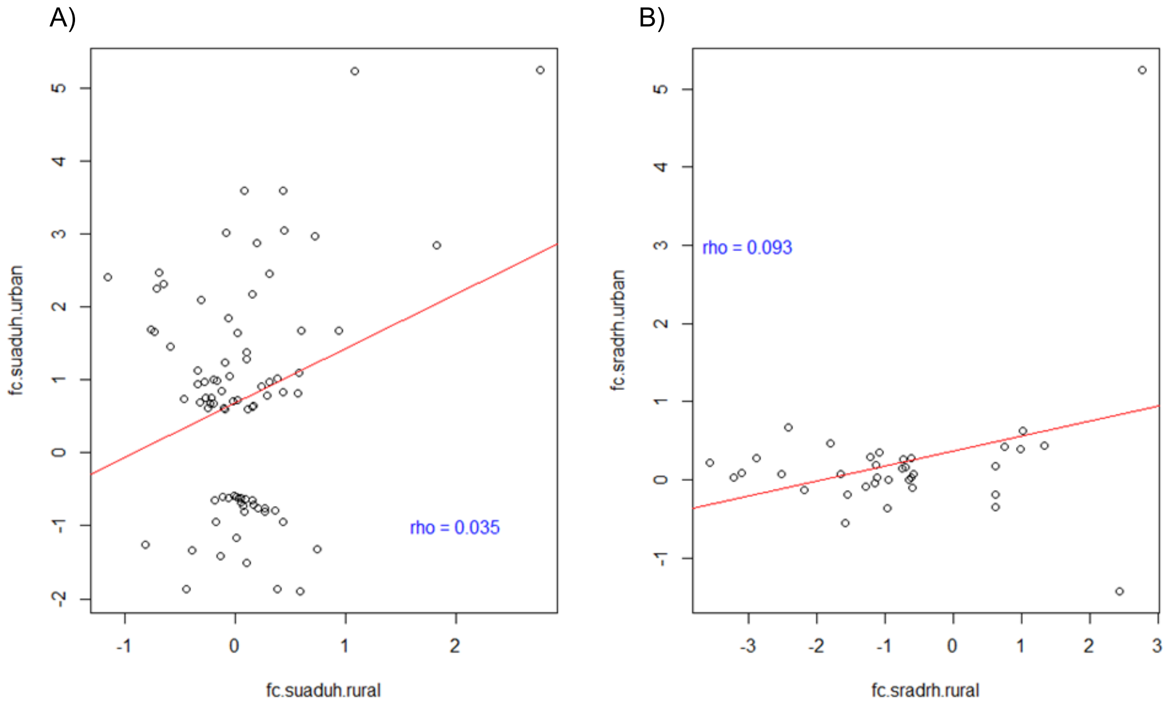


**S13 Fig. Correlation of the fold changes of the DEGs identified in one comparison with the fold changes of these genes in the respective other comparison**. A) Correlation of the fold changes of the genes identified with FDR < 0.05 and abs(fold change) > log2(1.5) in the comparison AD versus HC in urban samples. B) Correlation of the fold changes of the genes identified with FDR < 0.05 and abs(fold change) > log2(1.5) in the comparison AD versus HC in rural samples.

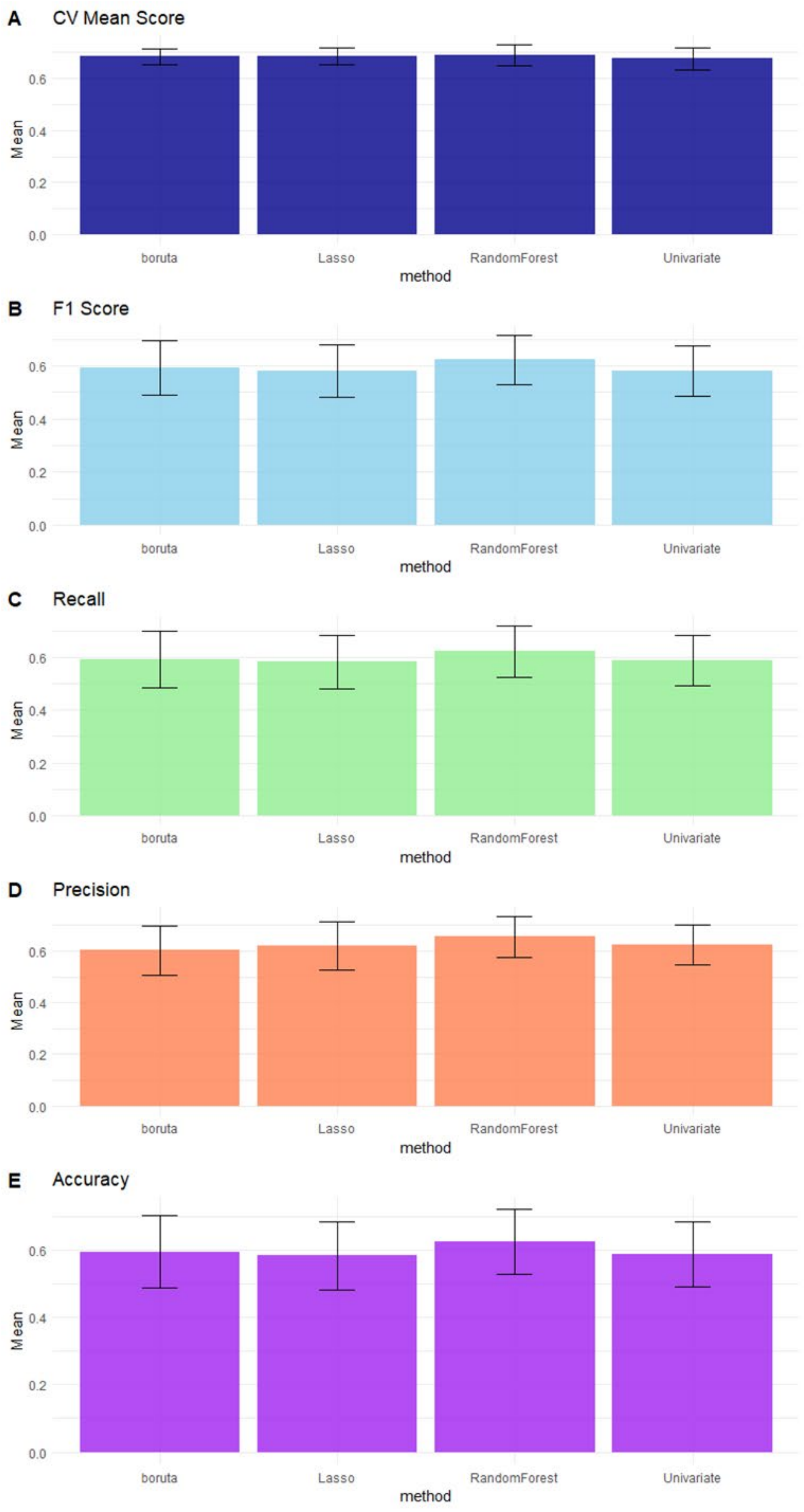


**S14 Fig.** Classification performance metrics of the four different ML methods.

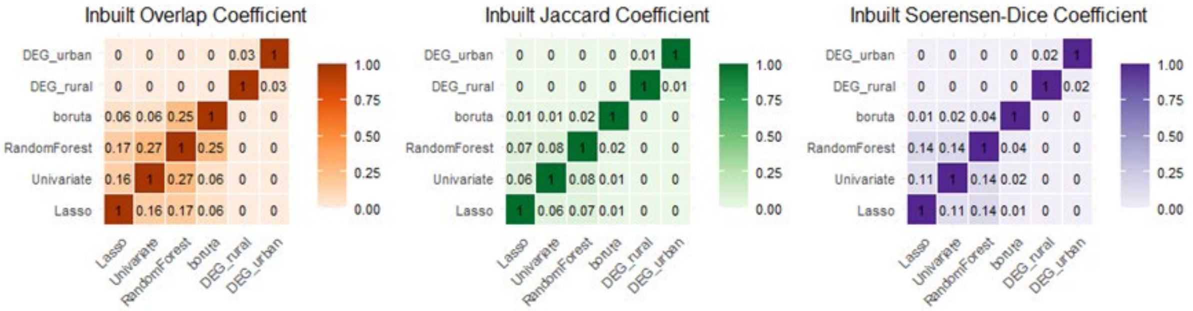


**S15 Fig.** Assessment of the overlap between selected feature lists from the four different ML Methods and the DGE lists using Overlap Coefficient, Jaccard Coefficient, and Sørensen-Dice Coefficient.

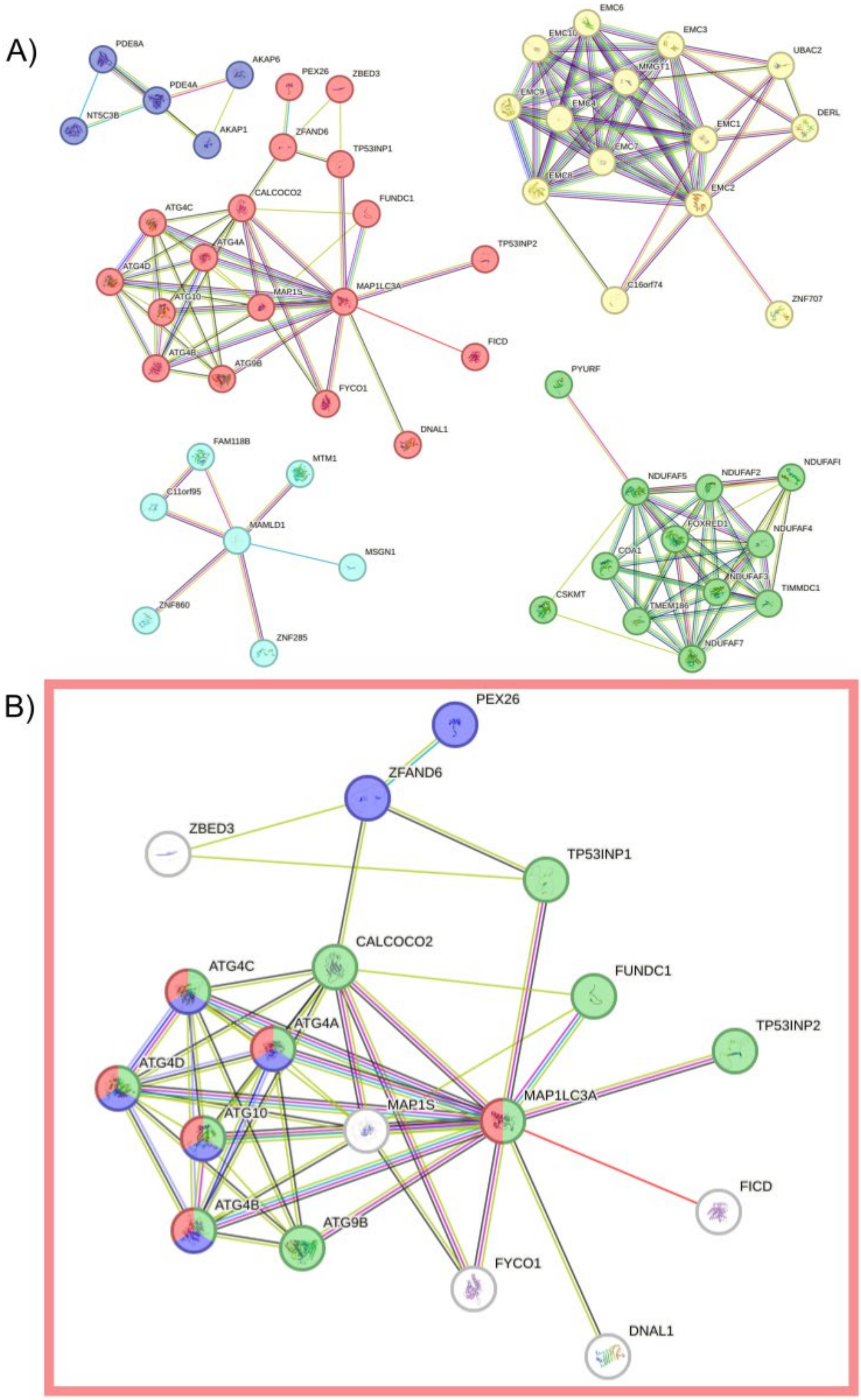


**S16 Fig. Genes in the violet DIABLO cluster subjected to network analysis and k-means subclustering in STRING, highlighting the red subcluster.** A) The genes from the violet DIABLO cluster subjected to a network analysis in STRING (Szklarczyk et al., 2023), followed by a k-means clustering with the distinct subclusters coloured in red, violet, cyan, yellow, and green. B) The red subcluster from A) features ZFAND6 and MAP1LC3A. The nodes are coloured according to their UniProt annotated keywords *Autophagy* (green), *Protein transport* (blue), and *Ubl conjugation pathway* (red).

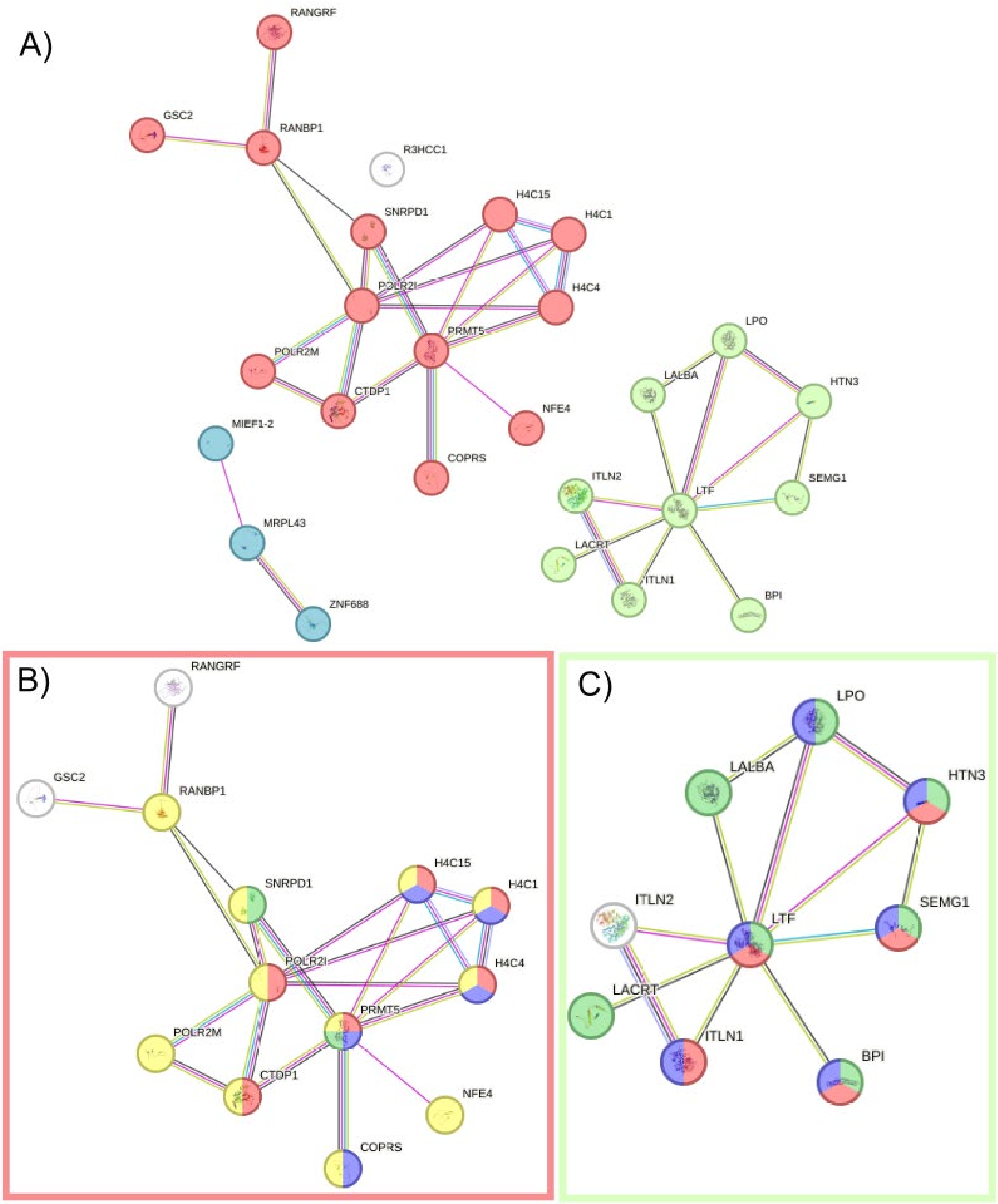


**S17 Fig. Genes in the orange DIABLO cluster subjected to network analysis and k-means subclustering in STRING, highlighting the red and green subclusters.** A) The genes from the orange DIABLO cluster subjected to a network analysis in STRING (Szklarczyk et al., 2023) with the interaction sources text mining, experiments, databases and co-expression with a maximum of 20 interactors in the 1st shell, followed by a k-means clustering with the distinct subclusters coloured in red, blue, and green. B) The nodes in the red subcluster from A) are coloured according to the functional enrichment terms: Subcellular localization: *Nucleus* (yellow); GO Biological Process: *Transcription, DNA-templated* (red) and *Chromatin organization* (purple); Local network cluster: *Methylosome* (green). C) The node in the green subcluster from A) are coloured according to the functional enrichment terms: GO Biological Process: *defense response to bacterium* (green); Reactome pathways: *Antimicrobial peptides* (red) and *Innate immune system* (purple).

### S2 File

#### SOS-ALL Data Analysis

Damir Zhakparov

2026-01-09

#### Table of Contents


**Introduction** 1
**1. Loading the packages required for the analyses** 2
**2. GeneSelectR Procedure** 2
  **2.1 Data Preparation and running GeneSelectR** 2
  2.2 Loading and processing the results of the differential gene expression (DGE) analysis 4
  **2.3 GeneSelectR Results Visualization** 6
    **2.3.1 Machine Learning Metrics Visualization** 6
    **2.2.2 Feature Importance Scoring** 8
  **2.3 Gene Ontology Enrichment Analysis** 16
**3. Data Integration with mixOmics** 26
  **3.1 Input Data Preparation** 27
    **3.1.1 Reading Input Data** 27
  **3.2 Data Integration with DIABLO** 28
    **3.2.1 Preparing the Data** 28
    **3.2. 2 Optimal Component Selection** 29
    **3.2.3 Feature Selection Tuning** 30
    **3.2.4 Final Model Fitting** 30
  **3.3 Results Inspection** 31
    **3.3.1 Variable Selection** 31
    **3.3.2 Visualization of DIABLO Components** 35
    **3.3.3 Individual and Variable Plots** 36
    **3.3.4 Loadings Analysis** 38


#### Introduction

This Markdown document provides an overview of the data analysis carried out on the SOS-ALL Project data, which focuses on studying Atopic Dermatitis (AD) in 149 African children that are either healthy or suffer from AD and live either in a rural or urban environment. Our primary objective is to identify common factors contributing to AD in both urban and rural environments, with the key variable in the dataset being the distinction between rural and urban settings. The analysis encompasses the following steps:

1) Examination of the differential gene expression (DEG) results;

2) Implementation of the GeneSelectR workflow on our dataset;

3) Integration of the different data types using the DIABLO method from the mixOmics R package.

#### 1. Loading the packages required for the analyses

```
library(dplyr)
library(stringr)
library(glue)
library(GeneSelectR)
library(mixOmics)
```

#### 2. GeneSelectR Procedure

GeneSelectR is a Machine Learning (ML)-powered workflow that aims to identify a subset of genes in transcriptomic datasets that have both computational and biological relevance.

##### 2.1 Data Preparation and running GeneSelectR

The initial step in the analysis involves loading the dataset and preparing it to be processed with GeneSelectR:

```
##############################################################################
###
# 1. Load in the data
##############################################################################
###
work_dir <- dirname(getwd())
data_dir <- file.path(work_dir, 'raw-data')
out_dir <- file.path(work_dir, 'results')

print(data_dir)

## [1] "C:/Users/damir/Documents/Projects/sosall/raw-data"

exp <- read.csv(file.path(data_dir, 'rna_normalized_logcpm.csv'))
meta <- read.csv(file.path(data_dir, 'rna_metadata.csv'))

exp <- as.data.frame(t(exp))
colnames(exp) <- exp[1,]
exp <- exp[-1,]
exp$SampleID <- rownames(exp)

# split treatment into two columns
```

```r
meta[,c('location', 'diagnosis')] <- str_split_fixed(meta[['treatment']],'_',
n=Inf)

# join the meta data with the exp dataframe
exp <- inner_join(exp, meta, by = c('SampleID' = 'X'))
rownames(exp) <- exp$SampleID
exp <- exp %>%
   dplyr::select(-SampleID) %>%
   dplyr::select(-names) %>%
   dplyr::select(-type) %>%
   dplyr::select(-location) %>%
   dplyr::select(-treatment)

X <- exp %>% dplyr::select(-diagnosis)
y <- exp['diagnosis']
y <- as.factor(y$diagnosis)
y <- as.integer(y)
```

Then, we can run the main GeneSelectR function specifying the following parameters:

- X_train - the training data array
- y_train - vector with the training labels
- njobs - number of cores to use for calculations, in this case -1 stands for using all available cores
- n_splits - number of train/test splits to assess the data splitting variance
- max_features - maximum number of features to pick
- calculate_permutation_importance - flag whether to calculate permutation feature importance

These parameters were used as a trade-off between calculation time and optimal algorithm performance.

```r
all_selection  <- GeneSelectR::GeneSelectR(X_train = X,
                                           y_train = y,
                                           njobs = -1L,
                                           n_iter = 100L,
                                           n_splits = 5L,
                                           max_features = 200L,
                                           calculate_permutation_importance =
TRUE)



Specification of the GeneSelectR output:

- ‘best_pipeline’: A named list containing parameters of the best performer
pipeline.

‘cv_results’: Results of cross-validation for each pipeline.

‘inbuilt_feature_importance’: Aggregated inbuilt feature importance scores.
```

```
'test_metrics': If the perform_split parameter was set to TRUE, a dataframe
of test metrics for each pipeline is returned.

-'cv_mean_score': A dataframe that summarizes the mean cross-validation
scores.

'permutation_importance': If permutation importance was calculated, its mean
values are returned.
```

#### 2.2 Loading and processing the results of the differential gene expression (DGE) analysis

To compare the ML-selected features with those resulting from the differential gene expression (DGE) analysis, we load the previously obtained DGE analysis results:

```r
deg.urban <- read.csv(file.path(data_dir,'treatU_AD_vs_U_H.csv'), header =
TRUE) # urban HC vs urban AD; 82 DEGs

deg.rural <- read.csv(file.path(data_dir,'treatR_AD_vs_R_H.csv'), header =
TRUE) # rural HC vs rural AD; 36 DEGs
```

The differentially expressed genes (DEGs) were obtained with edgeR and contain typical output columns from this method such as log fold change (logFC), false discovery rate (FDR) etc.

We are only interested in differentially expressed genes (DEGs) that have passed a significance (FDR < 0.05) and log fold change threshold (abs(logFC) > log2(1.5)). To filter them out we will use a helper function:

```r
#' Subset differentially expressed genes (DEGs)
#'
#' This function subsets a dataframe for differentially expressed genes based
on
#' specific criteria. It checks for the presence of column 'X1' or 'X',
subsets
#' the dataframe based on log fold change and FDR thresholds, and extracts
gene ids.
#'
#' @param df A dataframe containing gene expression data with potential
columns 'X1' or 'X',
#'   'gene_name', 'logFC', 'logCPM', 'F', 'PValue', 'FDR', and
'mlog10PValue'.
#' @return A dataframe with columns for the identifier (either 'X1' or 'X'),
ensembl_id,
#'   and gene_name, filtered by the specified logFC and FDR thresholds.
#' @examples
#' # Assuming `data` is your dataframe with the necessary columns
#' subsetted_genes <- subset_DEGs(data)
#'
```

```r
#' @importFrom stringr str_split_fixed
#' @importFrom dplyr select, filter
subset_DEGs <- function(df) {
  require(stringr)
  require(dplyr)

  # check which column exists, 'X1' or 'X'
  col <- ifelse('X1' %in% colnames(df), 'X1', ifelse('X' %in% colnames(df),
'X', stop("Neither 'X1' nor 'X' found in column names.")))

  # subset DEGs
  filtered_df <- df[,c(col,'gene_name',"logFC", "logCPM", "F",
"PValue","FDR","mlog10PValue")]
  filtered_df <- filtered_df[which((df[,"logFC"] > log2(1.5) | df[,"logFC"] <
-log2(1.5)) & df[,"FDR"] <0.05),]

  # extract gene ids
  result_df <- filtered_df
  result_df[,c('ensembl_id', 'gene_name')] <-
str_split_fixed(result_df[[col]],'__', n=Inf)
  result_df <- result_df %>% dplyr::select(col, ensembl_id, gene_name)

  return(result_df)
}
```

And then apply the function to our DGE analysis results:

```r
deg.urban <- subset_DEGs(deg.urban)

## Warning: Using an external vector in selections was deprecated in
tidyselect 1.1.0.
## ℹ Please use `all_of()` or `any_of()` instead.
##   # Was:
##   data %>% select(col)
##
##   # Now:
##   data %>% select(all_of(col))
##
## See <https://tidyselect.r-lib.org/reference/faq-external-vector.html>.
## This warning is displayed once every 8 hours.
## Call `lifecycle::last_lifecycle_warnings()` to see where this warning was
## generated.

deg.rural <- subset_DEGs(deg.rural)
```

We also need to clean gene names in our GeneSelectR results object for subsequent steps:

```r
# Remove the gene symbol from the feature names
all_selection@inbuilt_feature_importance <-
lapply(all_selection@inbuilt_feature_importance, function(x) {x$feature <-
sub("__.+$", "", x$feature); x})
```

```r
all_selection@permutation_importance <-
lapply(all_selection@permutation_importance, function(x) {x$feature <-
sub("__.+$", "", x$feature); x})
```

#### 2.3 GeneSelectR Results Visualization

##### 2.3.1 Machine Learning Metrics Visualization

First, we will inspect Machine Learning (ML) performance on our data by calling the plot_metrics function():

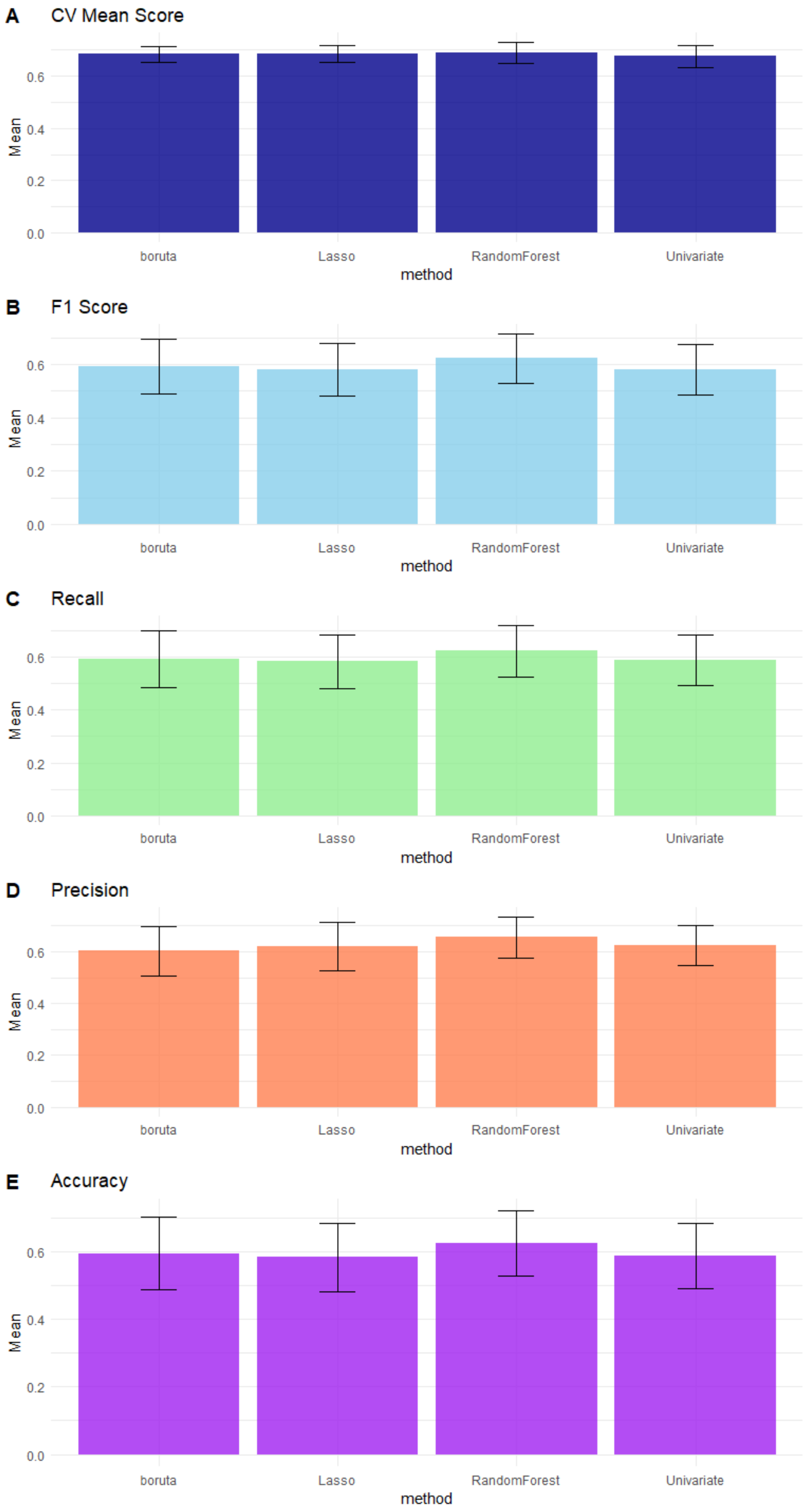
A
CV Mean Score
B
F1 Score
C
Recall
D
Precision
E
Accuracy
Mean
method
boruta
Lasso
RandomForest
Univariate
0.0
0.2
0.4
0.6

The overall performance seems comparable across the four different methods, with Random Forest showing marginally better performance. Relying solely on performance metrics makes it therefore inconclusive to determine the superior method. Thus, additional analyses are required.

##### 2.2.2 Feature Importance Scoring

Before we proceed it's important to filter out inconsistent features (e.g. features that show up less than certain number across different iterations):

Here we define two helper functions (please refer to the GeneSelectR documentation for the dataframe structure):

```r
#' Filter rows based on the amount of missing data in specified columns
#'
#' This function filters rows in a dataframe based on the proportion of
missing
#' data (NA values) in columns starting from the 4th column to the last
column.
#' Rows are kept if the proportion of missing data in these columns is less
#' than or equal to the specified threshold.
#'
#' @param data A dataframe containing the data to be filtered.
#' @param threshold A numeric value between 0 and 1 indicating the maximum
#' allowed proportion of missing data for a row to be kept. Default is 0.8.
#'
#' @return A dataframe with rows filtered based on the specified missing data
threshold.
filter_rank_columns <- function(data, threshold = 0.8) {
  keep <- rowMeans(is.na(data[, 4:ncol(data)])) <= threshold
  return(data[keep, ])
}


#' Sort a dataframe by descending order of a specific column
#'
#' This function sorts a dataframe in descending order based on the values of
#' the 'mean_importance' column. It is intended to organize dataframes by the
#' importance of variables, assuming 'mean_importance' represents some form
of
#' statistical or model-derived importance metric.
#'
#' @param df A dataframe that includes a 'mean_importance' column.
#'
#' @return A dataframe sorted in descending order by the 'mean_importance'
column.
sort_df <- function(df) {
  df_sorted <- df[order(-df$mean_importance), ]
  return(df_sorted)
}
```

For each ML algorithm, we will keep those genes that appear in at least 2 out of 10 iterations:

```
#80%
# permutation importance
all_selection@inbuilt_feature_importance <-
lapply(all_selection@inbuilt_feature_importance, filter_rank_columns,
threshold = 0.8)
all_selection@inbuilt_feature_importance <-
lapply(all_selection@inbuilt_feature_importance, sort_df)

# permutation importance
all_selection@permutation_importance <-
lapply(all_selection@permutation_importance, filter_rank_columns, threshold =
0.8)
all_selection@permutation_importance <-
lapply(all_selection@permutation_importance, sort_df)
```

On the selection step we calculated method specific and permutation based feature importance scores. To visualize top 10 features for every method and both ways of feature importance estimation we will run the following line:

```
# feature importance
all_selection_importance <-
GeneSelectR::plot_feature_importance(all_selection, top_n_features = 10)
all_selection_importance

## $Lasso
```

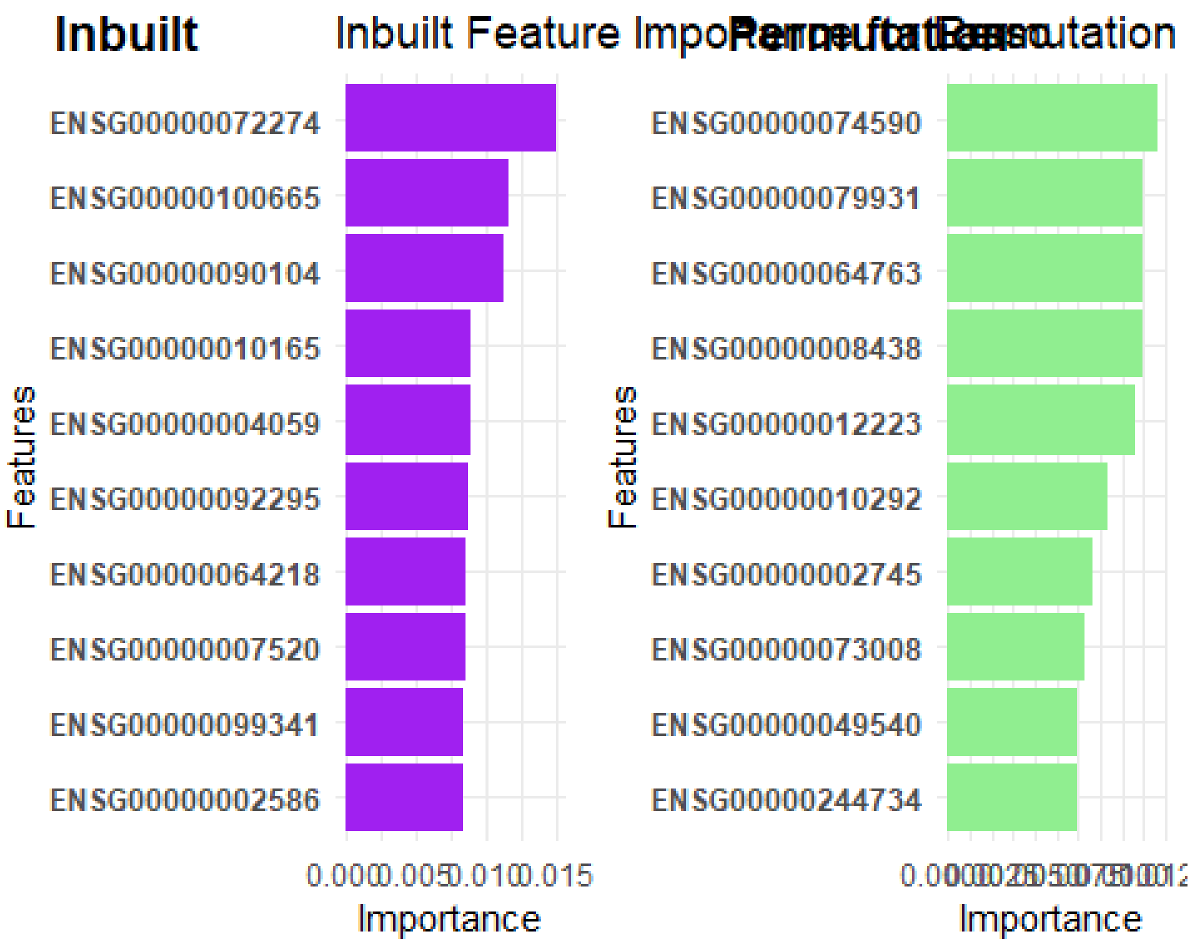
Inbuilt
Inbuilt Feature Importance
Permutation Based
Permutation
Features
ENSG00000072274
ENSG00000100665
ENSG00000090104
ENSG00000010165
ENSG00000004059
ENSG00000092295
ENSG00000064218
ENSG00000007520
ENSG00000099341
ENSG00000002586
0.000
0.005
0.010
0.015
Importance
Features
ENSG00000074590
ENSG00000079931
ENSG00000064763
ENSG00000008438
ENSG00000012223
ENSG00000010292
ENSG00000002745
ENSG00000073008
ENSG00000049540
ENSG00000244734
Importance

```
##
## $Univariate
```

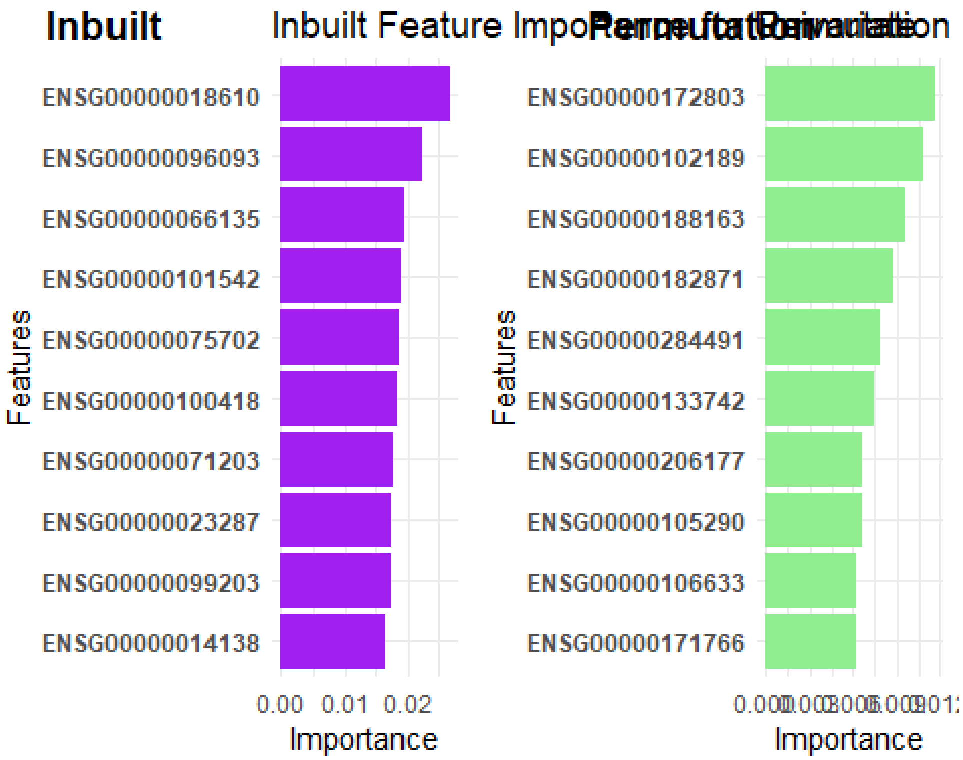


```
##
## $RandomForest
```

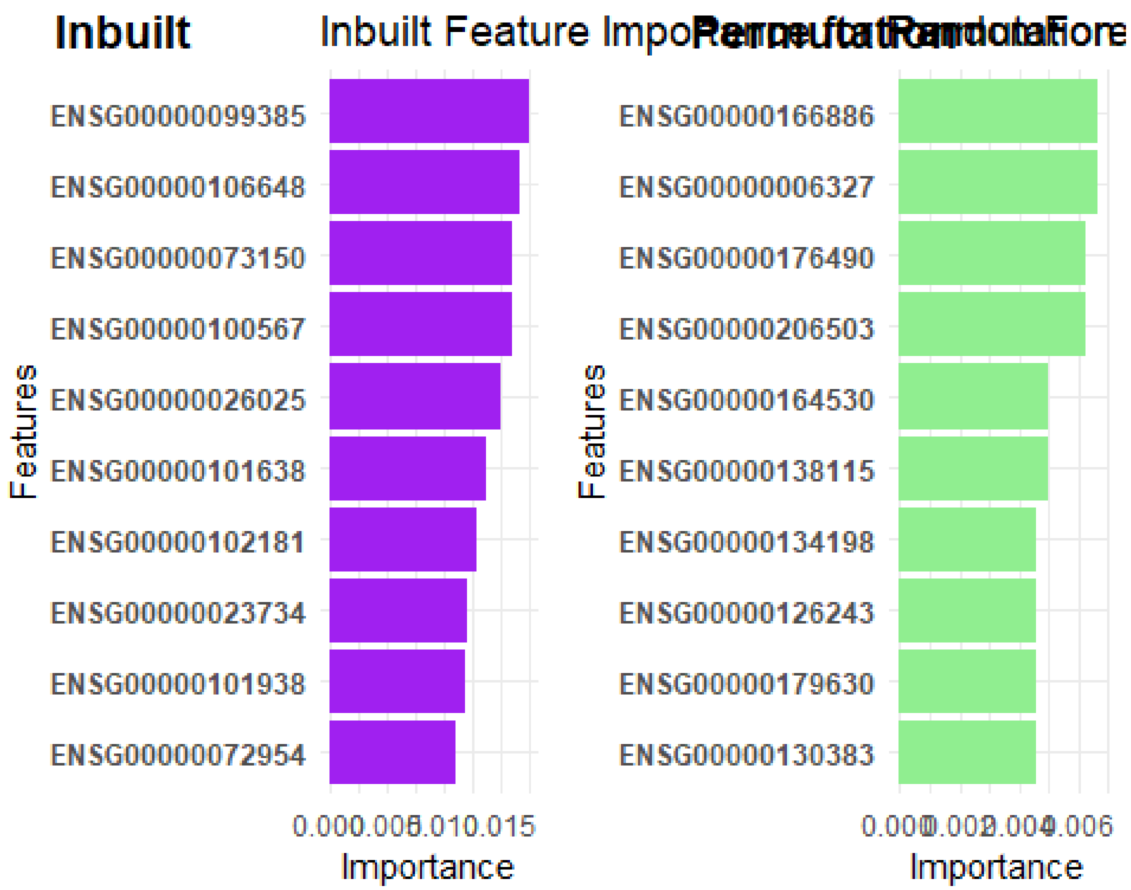
Inbuilt
Inbuilt Feature Impo
Permutation Random Fore
ENSG00000099385
ENSG00000106648
ENSG00000073150
ENSG00000100567
ENSG00000026025
ENSG00000101638
ENSG00000102181
ENSG00000023734
ENSG00000101938
ENSG00000072954
Features
0.000 0.005 0.010 0.015
Importance
ENSG00000166886
ENSG00000006327
ENSG00000176490
ENSG00000206503
ENSG00000164530
ENSG00000138115
ENSG00000134198
ENSG00000126243
ENSG00000179630
ENSG00000130383
Features
0.000 0.002 0.004 0.006
Importance

```
##
## $boruta
```

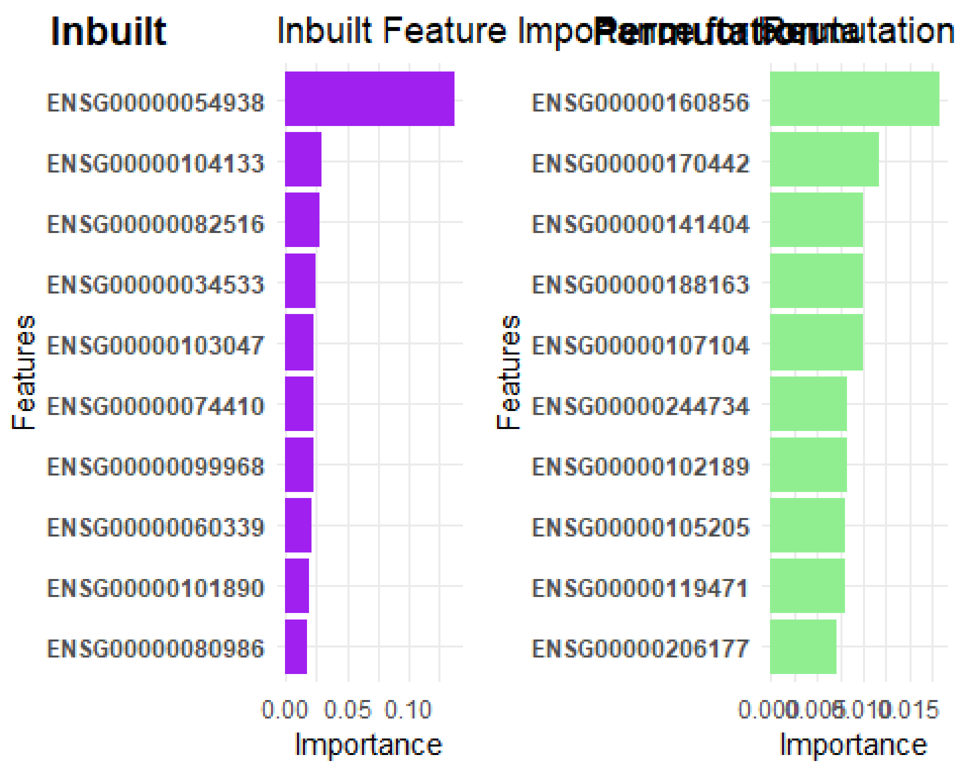


In addition to that, we can also inspect whether there is an overlap between feature selection lists produced by different methods as well as with differential gene expression lists:

```
# get background list (list of all genes that were used in the selection
process)
background <- colnames(X)

# remove gene symbols and only keep ENSEMBL IDs
background <- sub("__.+$", "", background)

# Overlap calculation
custom_all <- list(background = background,
                   DEG_rural = as.character(deg.rural$X),
                   DEG_urban = as.character(deg.urban$X1))

all_selection_overlap <-
GeneSelectR::calculate_overlap_coefficients(all_selection,

custom_lists = list(DEG_rural = as.character(deg.rural$X),

DEG_urban = as.character(deg.urban$X1)))
GeneSelectR::plot_overlap_heatmaps(all_selection_overlap)
```

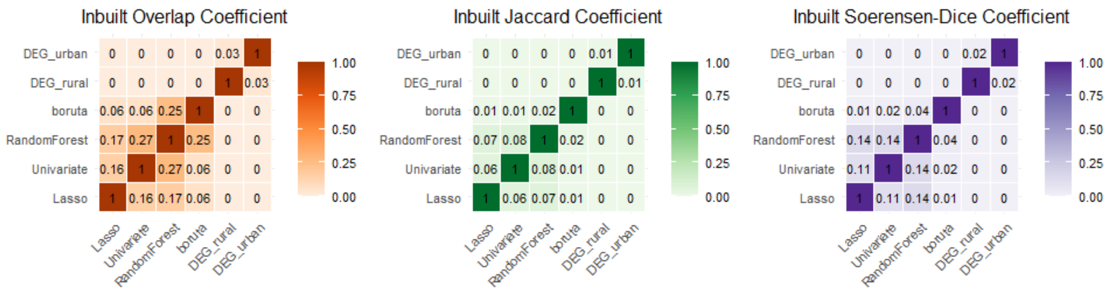


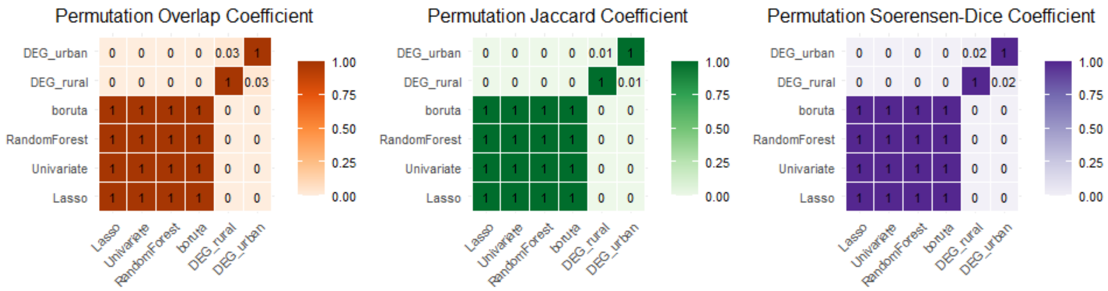


We don't observe too much overlap between either of the gene lists. As for permutation importance lists, the reason why the lists are identical is that all features are preserved at every iteration. Therefore, overlap analysis is not suitable for this method.

In addition to heatmaps, overlap can be also visualized with upset plot:

```
plot_upset(all_selection, custom_lists = list(DEG_rural = 
as.character(deg.rural$X), DEG_urban = as.character(deg.urban$X1)))

## $inbuilt_importance
```

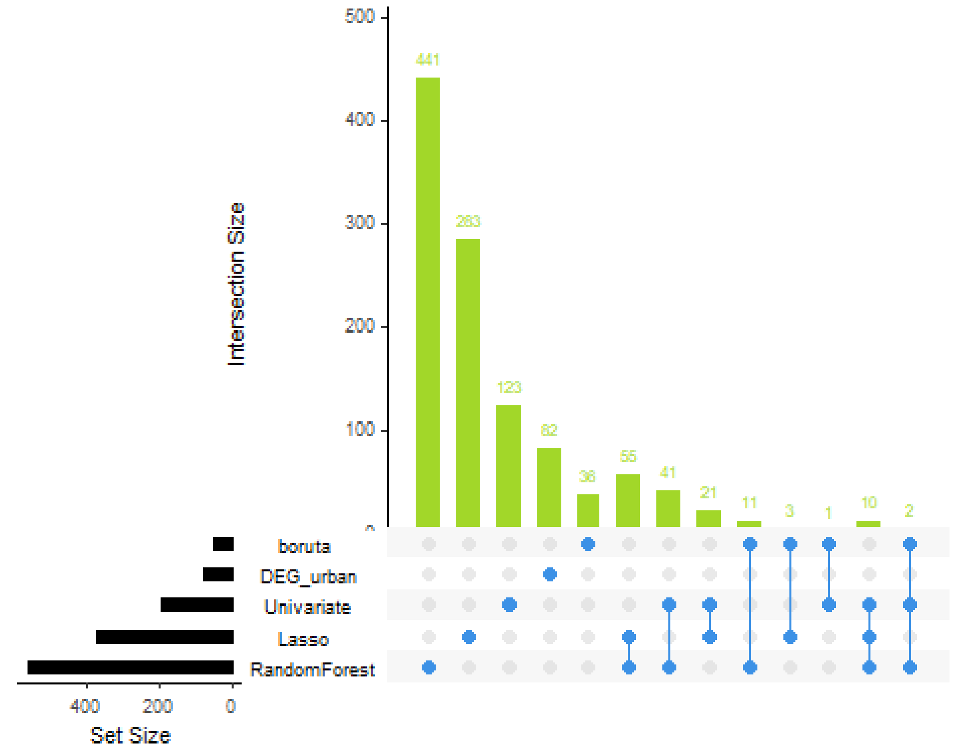


```
##
## $permutation_importance
```

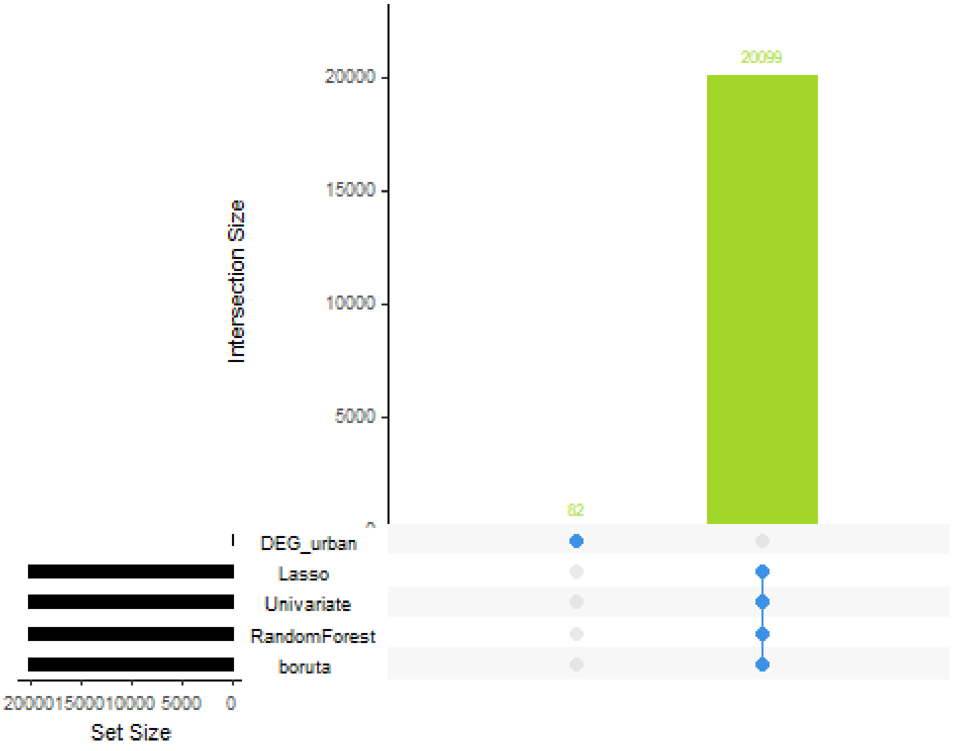

Since inbult feature importance scoring method yields more manageable lists, we will proceed with features that were deemed important according to their inbuilt feature importance scores.

##### 2.3 Gene Ontology Enrichment Analysis

In order to estimate biological relevance of feature lists we will proceed with gene ontology enrichment (GO) analysis. We will mine Biological Process ontology (GOBP).
We create annotation db object to retrieve gene identifiers:

```
# create annotation db object
ah <- AnnotationHub::AnnotationHub()
human_ens <- AnnotationHub::query(ah, c("Homo sapiens", "EnsDb"))
human_ens <- human_ens[['AH98047']]

## loading from cache

## require("ensembldb")

annotations_ahb <- ensembldb::genes(human_ens, return.type = "data.frame")
%>%
  dplyr::select(gene_id,gene_name,entrezid,gene_biotype)
```

We also need to clean up ENSEMBL IDs in DEG lists:

```
# remove the gene symbols from custom lists
custom_all <- lapply(custom_all, function (x) {x <- sub("__.+$", "", x); x})
```

To annotate genes in feature selection lists we will run the helper function from GeneSelectR:

```
# annotate the gene lists
all_selection.annot <- GeneSelectR::annotate_gene_lists(all_selection,
                                                        custom_lists =
custom_all,
                                                        annotations_ahb =
annotations_ahb,
                                                        format = 'ENSEMBL')

## Warning in clusterProfiler::bitr(original_ids, fromType = "ENSEMBL",
toType =
## c("SYMBOL", : 0.27% of input gene IDs are fail to map...

## Warning in clusterProfiler::bitr(original_ids, fromType = "ENSEMBL",
toType =
## c("SYMBOL", : 0.18% of input gene IDs are fail to map...

## Warning in clusterProfiler::bitr(original_ids, fromType = "ENSEMBL",
toType =
## c("SYMBOL", : 2.97% of input gene IDs are fail to map...

## Warning in clusterProfiler::bitr(original_ids, fromType = "ENSEMBL",
```

```
toType =
## c("SYMBOL", : 2.97% of input gene IDs are fail to map...

## Warning in clusterProfiler::bitr(original_ids, fromType = "ENSEMBL",
toType =
## c("SYMBOL", : 2.97% of input gene IDs are fail to map...

## Warning in clusterProfiler::bitr(original_ids, fromType = "ENSEMBL",
toType =
## c("SYMBOL", : 2.97% of input gene IDs are fail to map...

## Warning in clusterProfiler::bitr(original_ids, fromType = "ENSEMBL",
toType =
## c("SYMBOL", : 2.97% of input gene IDs are fail to map...

## Warning in clusterProfiler::bitr(original_ids, fromType = "ENSEMBL",
toType =
## c("SYMBOL", : 2.97% of input gene IDs are fail to map...
```

With annotated feature selection lists we can then run GOBP enrichment for both inbuilt and permutation importance based lists:

```
all_selection.GO_permutation <-
GeneSelectR::GO_enrichment_analysis(all_selection.annot,
                                                        list_type =
'permutation',
                                                        background =
all_selection.annot@permutation$background@ENSEMBL,
                                                        keyType = 'ENSEMBL')

## Performing GO Enrichment analysis for the:Lasso

## Performing GO Enrichment analysis for the:Univariate

## Performing GO Enrichment analysis for the:RandomForest

## Performing GO Enrichment analysis for the:boruta

## Performing GO Enrichment analysis for the:DEG_rural

## Performing GO Enrichment analysis for the:DEG_urban

all_selection.GO_inbuilt <-
GeneSelectR::GO_enrichment_analysis(all_selection.annot,
                                                        list_type =
'inbuilt',
                                                        background =
all_selection.annot@inbuilt$background@ENSEMBL,
                                                        keyType = 'ENSEMBL')

## Performing GO Enrichment analysis for the:Lasso
```

```
## Performing GO Enrichment analysis for the:Univariate

## Performing GO Enrichment analysis for the:RandomForest

## Performing GO Enrichment analysis for the:boruta

## Performing GO Enrichment analysis for the:DEG_rural

## Performing GO Enrichment analysis for the:DEG_urban
```

We can visualize the fraction of terms of interest in our list that stem from a common ancestor node that is related to some process. For example, we can take two parent terms that are related to immune system processes (GO:0002376) and defense against bacteria (GO:0042742), two terms that are of a big interest in the context of AD.

```
# fractions of terms of interest
# GO:0002376 immune system process
# defense against bacteria  GO:0042742
all_selection_immune <-
compute_GO_child_term_metrics(all_selection.GO_inbuilt,
                              GO_terms = c('GO:0002376', 'GO:0042742'),
                              ontology = 'BP',
                              plot = TRUE)
```

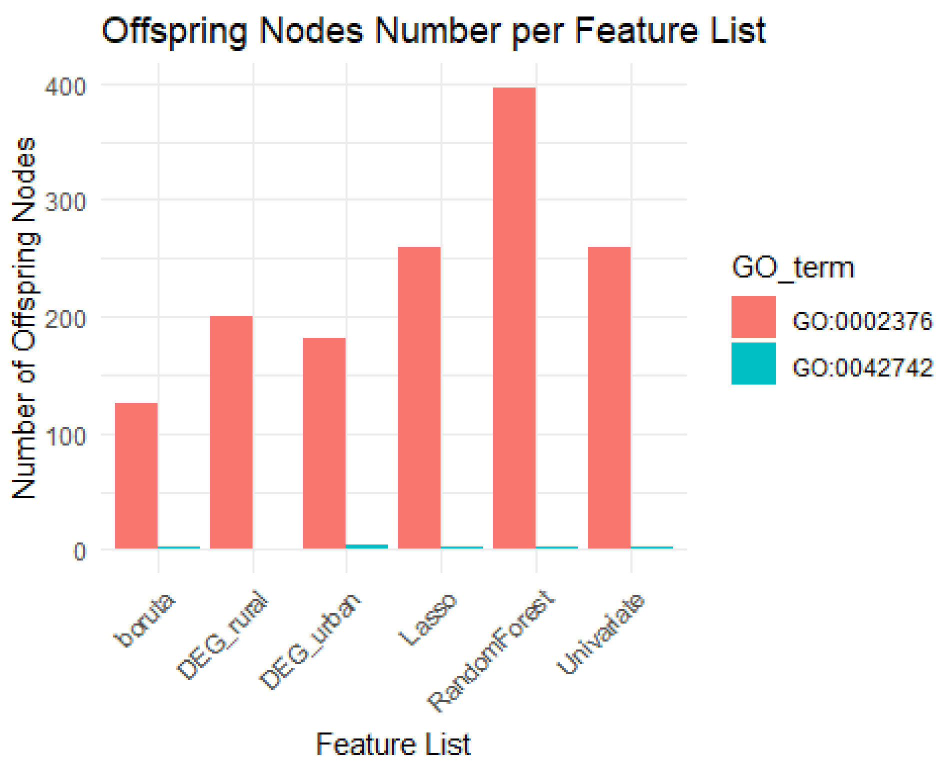


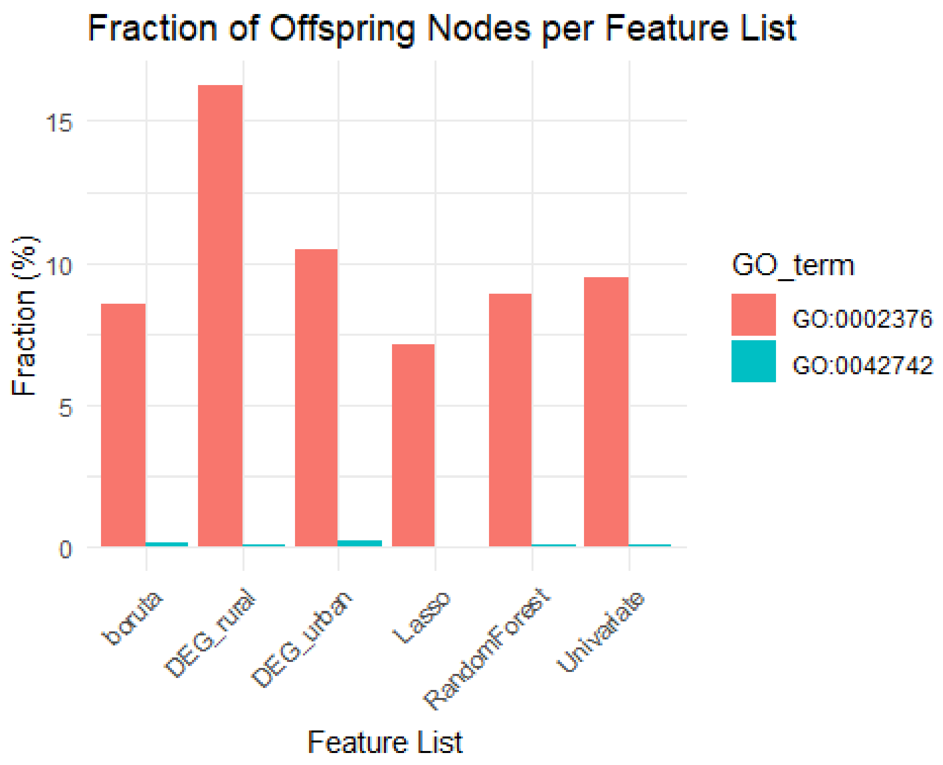


In the inbult feature selection list …

```
# convert to a list of dataframes
all_selection.GO_permutation <- list(Lasso  =
```

```
all_selection.GO_permutation$Lasso@result,
                         Univariate  =
all_selection.GO_permutation$Univariate@result,
                         boruta  =
all_selection.GO_permutation$boruta@result,
                         RandomForest  =
all_selection.GO_permutation$RandomForest@result,
                         DEG_rural =
all_selection.GO_permutation$DEG_rural@result,
                         DEG_urban =
all_selection.GO_permutation$DEG_urban@result)

all_selection.GO_inbuilt <- list(Lasso  =
all_selection.GO_inbuilt$Lasso@result,
                         Univariate  =
all_selection.GO_inbuilt$Univariate@result,
                         boruta  = all_selection.GO_inbuilt$boruta@result,
                         RandomForest  =
all_selection.GO_inbuilt$RandomForest@result,
                         DEG_rural =
all_selection.GO_inbuilt$DEG_rural@result,
                         DEG_urban =
all_selection.GO_inbuilt$DEG_urban@result)

#' Create a Dot Plot for Significant Gene Ontology (GO) Terms
#'
#' This function generates a dot plot visualizing significant Gene Ontology
(GO) terms
#' based on their p-values and q-values. It highlights the top 30 significant
GO terms
#' by arranging them in ascending order of their q-values. Each dot's size
represents the
#' gene count associated with the GO term, and its color indicates the
significance level
#' (-log10 transformed p-value).
#'
#' @param df A dataframe containing Gene Ontology (GO) term data, including
columns for
#'   `pvalue`, `qvalue`, `Count`, and `Description`.
#' @param title A character string specifying the title of the plot. Default
is an empty string.
#'
#' @return A ggplot object representing the dot plot of significant GO terms.
The plot is not
#'   displayed until it is printed or explicitly displayed in an R
environment that supports
#'   graphical output (e.g., RStudio).
#'
#' @examples
#' # Assuming `go_data` is a dataframe with columns: `pvalue`, `qvalue`,
```

```
`Count`, `Description`
#' go_data <- data.frame(
#'   Description = c("GO:0008150", "GO:0007049", "GO:0009987"),
#'   pvalue = c(0.01, 0.02, 0.04),
#'   qvalue = c(0.05, 0.07, 0.06),
#'   Count = c(100, 150, 120)
#' )
#' dotplot <- create_dotplot(go_data, title = "Top 30 Significant GO Terms")
#' print(dotplot) # To display the plot
#'
#' @import ggplot2
#'
create_dotplot <- function(df, title = '') {
  require(ggplot2)
  # Create a subset of the dataframe with significant GO terms (e.g.,
p.adjust < 0.05)
  df_sig <- df[df$pvalue < 0.05, ]

  # Arrange the dataframe by q-value (smaller values at the top)
  df_sig <- df_sig[order(df_sig$qvalue), ]
  df_sig <- df_sig[1:30,]
  # Create a dotplot
  ggplot(df_sig, aes(x = -log10(pvalue), y = reorder(Description, -qvalue)))
+
    geom_point(aes(size = Count, color = -log10(pvalue))) +
    scale_color_gradient(low = "blue", high = "red") +
    theme_minimal() +
    theme(axis.text.x = element_text(angle = 90, hjust = 1)) +
    labs(x = "GO Term", y = "-log10(p-value)", title = title,
         size = "Gene Count", color = "-log10(p-value)")
}

create_dotplot(all_selection.GO_inbuilt$Lasso, title = 'Lasso')
```

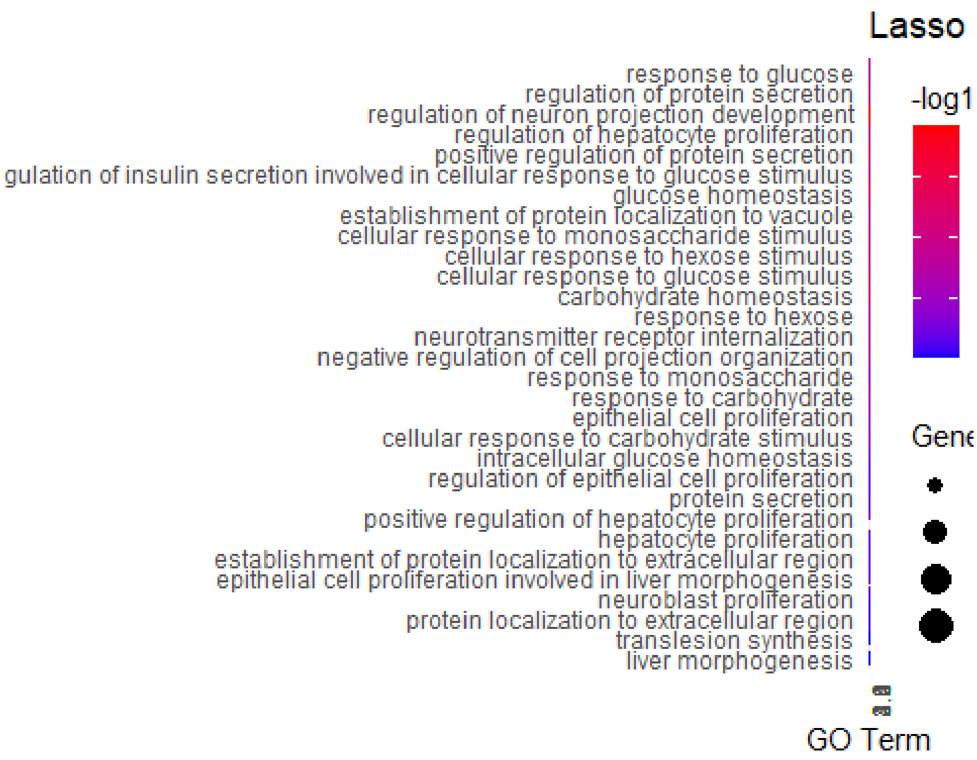


```r
create_dotplot(all_selection.GO_inbuilt$Univariate, title = 'Univariate')
```

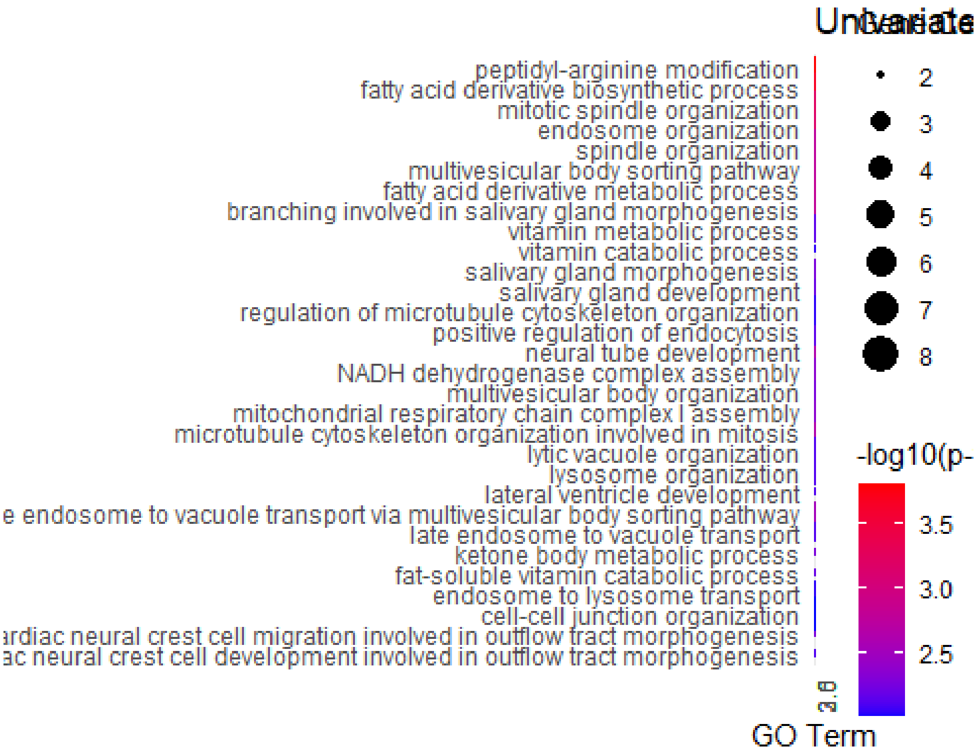


```r
create_dotplot(all_selection.GO_inbuilt$boruta, title = 'boruta')
```

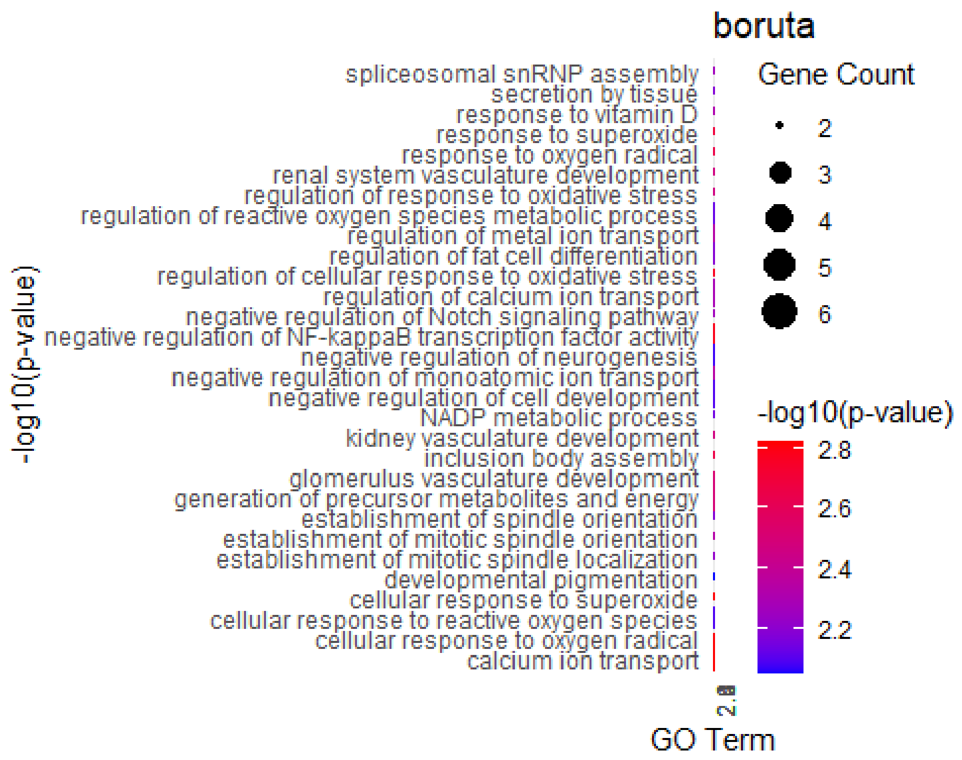


```
create_dotplot(all_selection.GO_inbuilt$RandomForest, title = 'RandomForest')
```

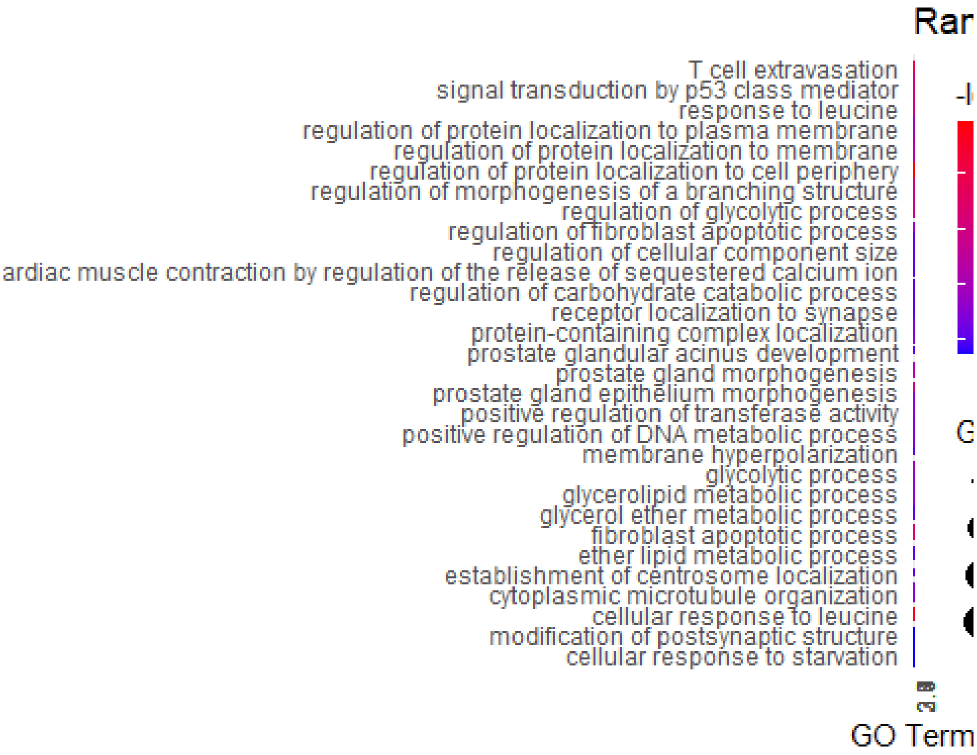


```
create_dotplot(all_selection.GO_inbuilt$DEG_rural, title = 'DEG_rural')
```

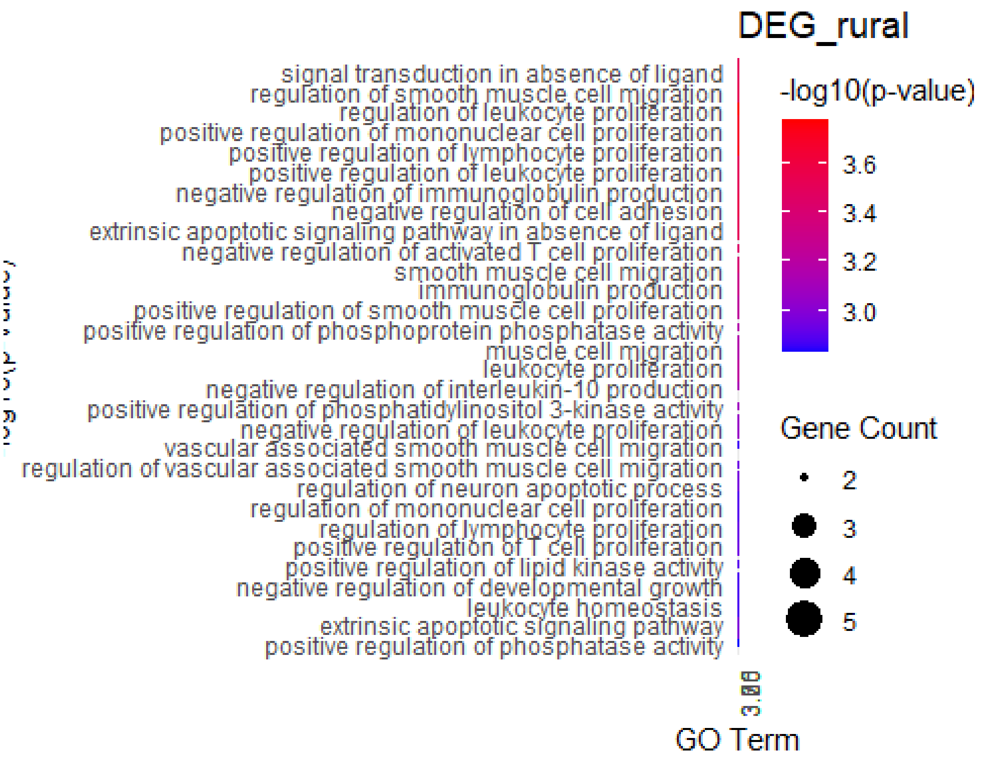


```r
create_dotplot(all_selection.GO_inbuilt$DEG_urban, title = 'DEG_urban')
```

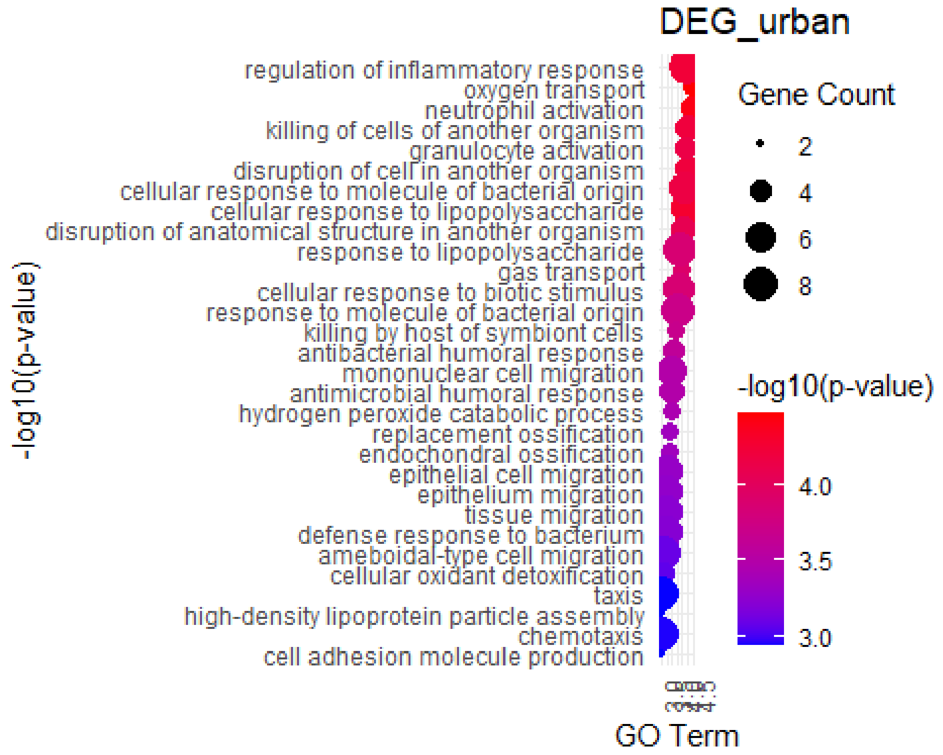

```
hmap =
simplifyEnrichment::simplifyGOFromMultipleLists(all_selection.GO_inbuilt,
                                              padj_column = 'pvalue',
                                              padj_cutoff = 0.01,
                                              ont = 'BP',
                                              measure = 'Lin',
                                              method = 'kmeans')

## Use column 'ID' as `go_id_column`.

## Loading required namespace: gridtext

## 382/5376 GO IDs left for clustering.

## Cluster 382 terms by 'kmeans'... 13 clusters, used 1.334591 secs.
## 'magick' package is suggested to install to give better rasterization.
##
## Set `ht_opt$message = FALSE` to turn off this message.
## Perform keywords enrichment for 13 GO lists...
## 'magick' package is suggested to install to give better rasterization.
##
## Set `ht_opt$message = FALSE` to turn off this message.
```

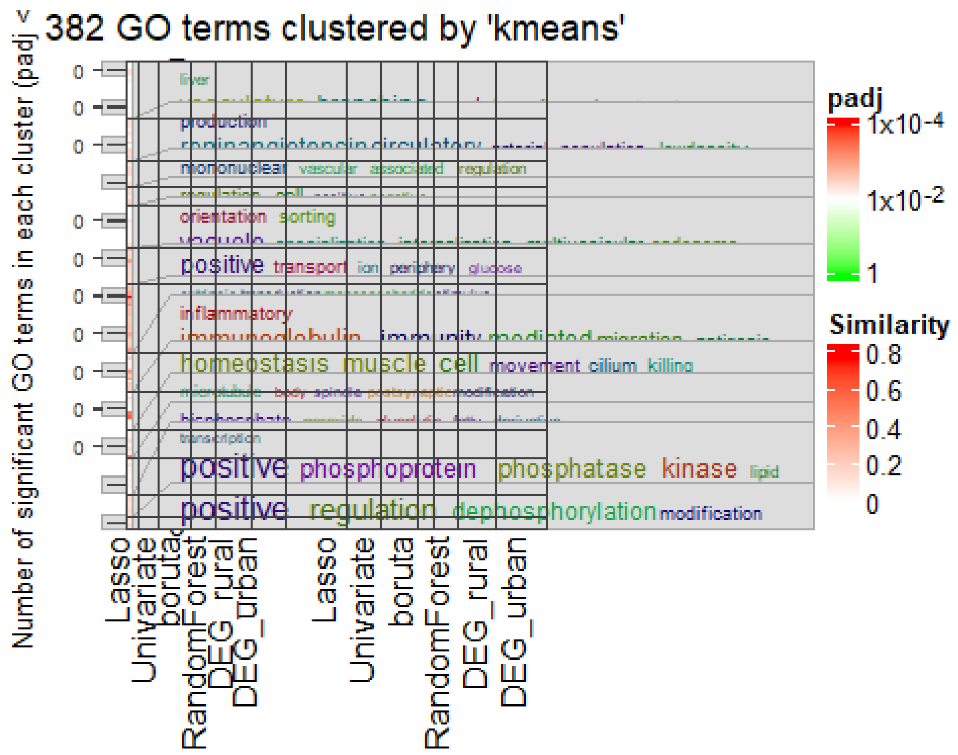


```
hmap =
simplifyEnrichment::simplifyGOFromMultipleLists(all_selection.GO_permutation,
                                              padj_column = 'pvalue',
                                              padj_cutoff = 0.01,
                                              ont = 'BP',
```

```
                                    measure = 'Lin',
                                    method = 'kmeans')

## Use column 'ID' as `go_id_column`.

## 170/6277 GO IDs left for clustering.

## Cluster 170 terms by 'kmeans'... 9 clusters, used 0.07781792 secs.
## 'magick' package is suggested to install to give better rasterization.
##
## Set `ht_opt$message = FALSE` to turn off this message.
## Perform keywords enrichment for 9 GO lists...
## 'magick' package is suggested to install to give better rasterization.
##
## Set `ht_opt$message = FALSE` to turn off this message.
```

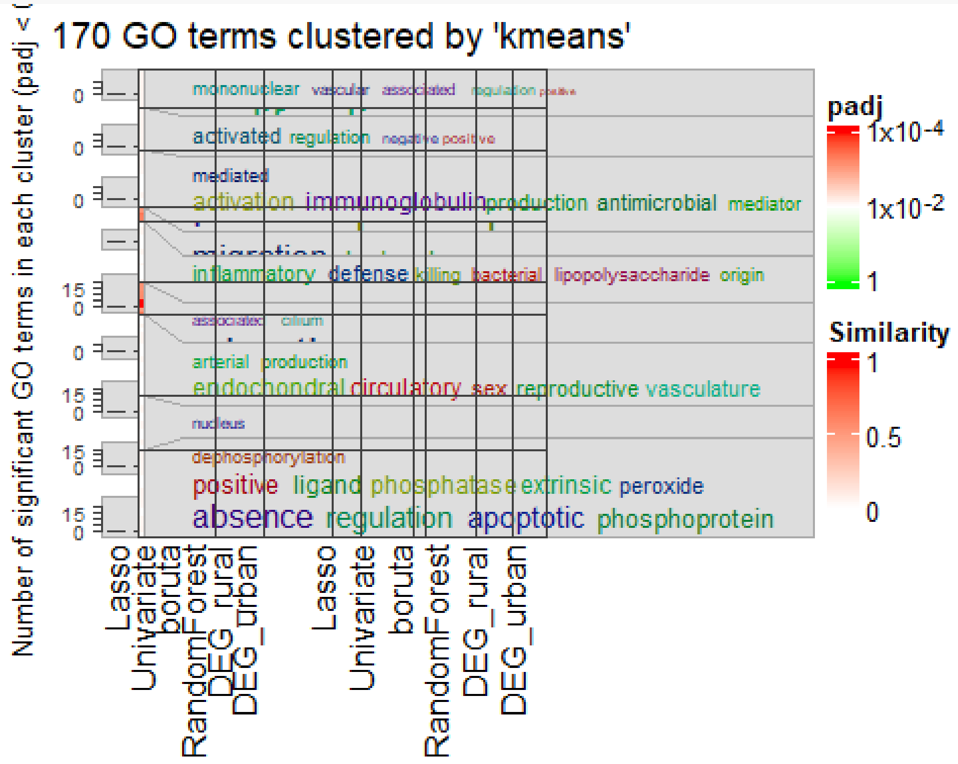


#### 3. Data Integration with mixOmics

mixOmics is a comprehensive R package designed for the exploration and integration of biological datasets through a variety of multivariate methods, including DIABLO (Data Integration Analysis for Biomarker discovery using Latent cOmponents). DIABLO is tailored for integrating multiple types of data (e.g., RNAseq, cytokine expression, and clinical questionnaire data) to uncover correlations between these datasets and a response variable, facilitating the identification of potential biomarkers.

#### 3.1 Input Data Preparation

##### 3.1.1 Reading Input Data

Our first step involves loading the RNAseq, clinical, and cytokine expression datasets. This step is foundational, ensuring that all subsequent analyses are based on accurately imported and formatted data. The RNAseq data (X) has already been prepared, while the clinical (Y) and cytokine (Z) data are read from CSV and text files, respectively. Special attention is paid to handling missing values, which are identified and managed appropriately to maintain data integrity.

```
# X - RNAseq dataframe has been obtained in the previous section
Y <-
read.csv(file.path(data_dir,'sosall_imputed_values','no_sp','41_imputed_value
s.csv'), sep = ',', na.strings = 'NA') #Clinical
Z <- read.csv(file.path(data_dir,'sos_all_cytokines.txt'), sep = '\t',
na.strings = c('NaN','NaN63')) # cytokines
```

We then load a list of 560 genes identified by a Random Forest analysis as being significant. This gene list serves as a filter to focus the RNAseq dataset on these key features, aligning the analysis with previously identified biological signals of interest.

```
rf560 <- read.csv(file = file.path(out_dir,'2023-07-
31','RandomForest560.txt'), sep = '\t', header = FALSE, skip = 1, col.names =
c('index','feature'))

feature_list <- rf560$feature
```

The RNAseq dataframe is subset to include only the genes present in the Random Forest list. This step narrows down the dataset to the most relevant features for the analysis, potentially enhancing the model’s performance and interpretability. Additionally, gene names are cleaned to remove unnecessary prefixes and standardize naming conventions.

```
X <- X[,colnames(X) %in% feature_list]
names(X) <- gsub("^ENSG[[:alnum:]]+__", "", names(X))
rownames(X) <- toupper(rownames(X))
```

To ensure the integration process proceeds smoothly, we take several steps to harmonize the data:

- Numerical Conversion: All values across datasets are converted to numerical format, as required by mixOmics for proper analysis.
- Patient ID Alignment: The datasets are checked and adjusted to ensure that only samples present across all datasets are included in the analysis. This step avoids mismatches and potential biases stemming from incomplete data.
- Ordering of Samples: The samples are ordered consistently across the datasets to prevent any misalignment during the integration process.
- Matrix Conversion: Finally, the datasets are converted into matrix format, which is necessary for the subsequent analysis steps in mixOmics.

```
Y <- Y %>% mutate_if(is.character, as.numeric)
X <- X %>% mutate_if(is.character, as.numeric)
Z <- Z %>% mutate_if(is.character, as.numeric)
```

```
## Warning: There were 2 warnings in `mutate()`.
## The first warning was:
## ℹ In argument: `IL.1beta = .Primitive("as.double")(IL.1beta)`.
## Caused by warning:
## ! NAs introduced by coercion
## ℹ Run `dplyr::last_dplyr_warnings()` to see the 1 remaining warning.
```

```
# subset the dataframe to have only shared samples
Z <- Z[tolower(rownames(Z)) %in% tolower(meta$X), ]
Y <- Y[tolower(rownames(Y)) %in% tolower(rownames(Z)), ]
X <- X[tolower(rownames(X)) %in% tolower(rownames(Z)), ]


# order X and Y to have them in the same order
Y = Y[order(match(rownames(Y), rownames(X))),]
X = X[order(match(rownames(X), rownames(Y))),]
Z = Z[order(match(rownames(Z), rownames(Y))),]

rownames(X) <- toupper(rownames(X))
rownames(Y) <- toupper(rownames(Y))
rownames(Z) <- toupper(rownames(Z))


# convert to matrices
X <- as.matrix(X)
Y <- as.matrix(Y)
Z <- as.matrix(Z)
```

#### 3.2 Data Integration with DIABLO

##### 3.2.1 Preparing the Data

You begin by constructing a consolidated list (X.train_full) containing the three datasets: RNA, Clinical, and Cytokine. This integration enables a holistic analysis, considering multiple biological dimensions. Additionally, the diagnosis labels are extracted and converted into a factor variable (Y.train_full), ensuring they are appropriately handled in the modeling process:

```
X.train_full <- list(RNA = X,
                     Clinical = Y,
                     Cytokine = Z)

Y.train_full <- as.factor(meta[toupper(meta$X) %in% rownames(Z),"diagnosis"])
```

##### 3.2. 2 Optimal Component Selection

To identify the most effective number of components for the model, we employ Leave-One-Out Cross-Validation (LOOCV) on a model with up to 10 components. This step is crucial for model optimization, as it helps determine the complexity required to best capture the underlying data structure without overfitting. The choice of the optimal number of components is informed by plotting the LOOCV results, which provides a visual assessment of model performance across different component counts:

```
##############################################################################
###
# Three datasets
##############################################################################
###
diablo.sosall_full <- block.splsda(X.train_full, Y.train_full, ncomp = 10,
near.zero.var = TRUE)
cv.diablo.sosall <- perf(diablo.sosall_full,
                         validation = 'loo',
                         progressBar = TRUE,
                         auc = TRUE)
```

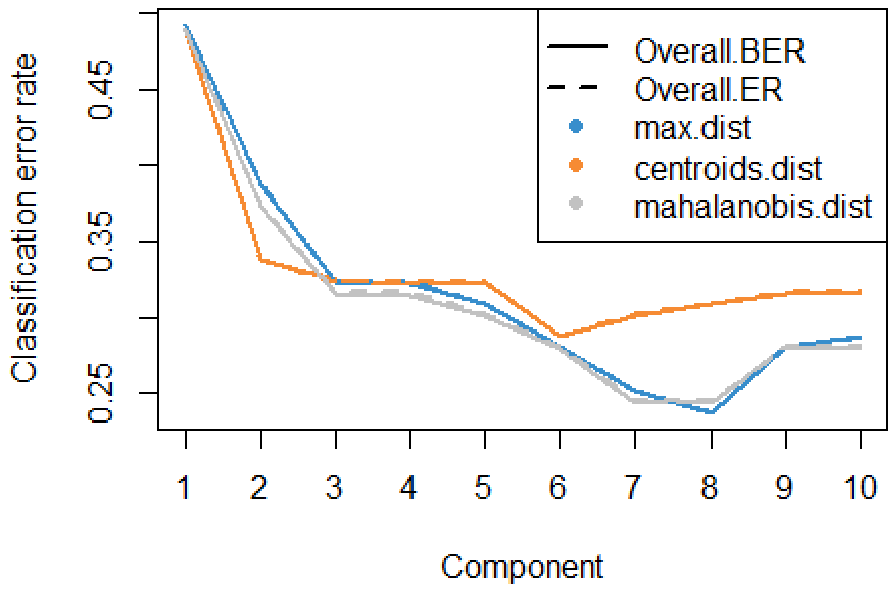


We want to maximize the distance between data points so we use max.dist parameters to determine an optimal number of components for the model. According to the plot the optimal number of components for our model is therefore 3.

##### 3.2.3 Feature Selection Tuning

The next step involves tuning the model to decide how many features to select from each dataset. By setting up a fully connected design matrix, you ensure that all datasets are considered both independently and in relation to the response variable. The tune.block.splsda function iterates over a predefined grid of feature counts (keepX.grid), assessing model performance through M-fold cross-validation. This process not only refines the model by selecting the most informative features but also tailors the integration to highlight relevant biological signals:

```
design <- matrix(1, ncol = length(X.train_full), nrow = length(X.train_full),
                 dimnames = list(names(X.train_full), names(X.train_full)))
diag(design) <- 0

keepX.grid <- list(RNA = c(20, 25, 30, 50, 100),
                   Clinical = c(3, 5, 10, 12, 15, 20, 25, 30, 50, 70),
                   Cytokine = c(5, 10, 15, 20, 30, 40, 45))

tune.diablo.sosall <- tune.block.splsda(X.train_full,
                                        Y.train_full,
                                        ncomp = ncomp,
                                        test.keepX = keepX.grid,
                                        validation = 'Mfold',
                                        folds = 10,
                                        nrepeat = 100,
                                        max.iter = 1000,
                                        design = design,
                                        dist = 'max.dist',
                                        scheme = 'factorial',
                                        progressBar = TRUE,
                                        near.zero.var = TRUE,
                                        BPPARAM =
BiocParallel::SnowParam(workers = 12)) #parallel::detectCores()-1))
keep.features <- tune.diablo.sosall$choice.keepX
```

##### 3.2.4 Final Model Fitting

With the optimal number of components and feature selection determined, you fit the final DIABLO model. This model is expected to leverage the strengths of each dataset, offering a nuanced understanding of the diagnostic labels.

```
#### fit the model with defined parameters
diablo.sosall_full <- block.splsda(X.train_full,
                              Y.train_full,
                              ncomp = ncomp,
                              keepX = keep.features,
                              near.zero.var = TRUE,
                              design = design)
```

#### 3.3 Results Inspection

##### 3.3.1 Variable Selection

By examining the variables selected in the first component across the three datasets, you gain insights into the most discriminative features driving the model. This step is instrumental in understanding the biological relevance of each dataset in the context of the diagnosis:

```
selectVar(diablo.sosall_full, block = 'RNA', comp = 1)
```

```
## $RNA
## $RNA$name
##  [1] "EMC2"     "IFT27"    "RASSF7"   "C5"       "MAP1LC3A" "MAMLD1"
##  [7] "MRI1"     "MYCBP2"   "NRDC"     "ZFAND6"   "SIRPG"    "NDUFAF5"
## [13] "PIK3R2"   "CXCL2"    "PDE4A"    "BCS1L"    "ADD1"     "FCGR2B"
## [19] "ACAP1"    "PRKCZ"
##
## $RNA$value
##             value.var
## EMC2     -0.372436188
## IFT27     0.340569061
## RASSF7    0.319714441
## C5        0.304846949
## MAP1LC3A -0.268674246
## MAMLD1   -0.266800670
## MRI1      0.261335781
## MYCBP2    0.259821097
## NRDC      0.250176396
## ZFAND6   -0.206094486
## SIRPG     0.205073288
## NDUFAF5  -0.176281942
## PIK3R2    0.170960275
## CXCL2    -0.159200691
## PDE4A    -0.142176013
## BCS1L     0.124641377
## ADD1      0.036951809
## FCGR2B    0.029949776
## ACAP1     0.010482508
## PRKCZ     0.009775259
##
##
## $comp
## [1] 1
```

```
selectVar(diablo.sosall_full, block = 'Clinical', comp = 1)
```

```
## $Clinical
## $Clinical$name
##  [1] "farmanimal_contact_mother"
```

```
##  [2] "farmanimal_contact_child"
##  [3] "fuel_cooking_Electricity_Gas"
##  [4] "fuel_cooking_Open_fires_outside"
##  [5] "sunlight_exp_winter"
##  [6] "sunlight_exp_summer"
##  [7] "fuel_cooking_Paraffin_Stove"
##  [8] "fuel_heating_Wood_Coal"
##  [9] "pets_dog"
## [10] "peanuts_exposure"
## [11] "delivery_mode"
## [12] "log_blood_counts_monocytes"
## [13] "henegg_exposure_regularity"
## [14] "fuel_cooking_heating_Paraffin"
## [15] "pets_cat"
## [16] "fuel_heating_Electricity"
## [17] "household_peoplelivingtogether"
## [18] "familyhistory_mother_other_allergic_disease"
## [19] "hayfever_ever"
## [20] "total_IgE"
## [21] "sIgG4_fish"
## [22] "wheat_exposure_regularity"
## [23] "sIgE_Der"
## [24] "cigarettesmoke_numberofpersons"
## [25] "household_otheryoungerchildren"
## [26] "household_otherolderchildren"
## [27] "paracetamol_exposure_time"
## [28] "sIgE_fish"
## [29] "log_blood_counts_plts"
## [30] "sIgE_hazelnut"
## [31] "anti_helminthic_exposure"
## [32] "sIgE_peanut"
## [33] "sIgG4_casein"
## [34] "solidfood_age_introduction"
## [35] "sIgE_CCD"
## [36] "paracetamol_exposure_age"
## [37] "othernuts_exposure"
## [38] "sIgE_soy"
## [39] "log_blood_counts_eosinophils"
## [40] "sIgG4_Der"
## [41] "antibiotic_exposure_last2months"
## [42] "fish_exposure"
## [43] "enrolment_age"
## [44] "henegg_exposure_age_first"
## [45] "breastfeeding_ever"
## [46] "peanuts_mother_exposure_avoidance"
## [47] "sIgE_egg"
## [48] "dairyproducts_exposure_age_first"
## [49] "fuel_heating_Other"
## [50] "sIgG4_peanut"
## [51] "immun_number"
```

```
## [52] "anti_helminthic_exposure_last"
## [53] "cowmilkformula_exposure"
## [54] "antibiotic_exposure"
## [55] "sIgG4_hazelnut"
## [56] "familyhistory_father_other_allergic_disease"
## [57] "henegg_exposure"
## [58] "formula_age_introduction"
## [59] "breastfeeding_uptoage_age_complete"
## [60] "log_blood_counts_hb"
## [61] "log_blood_counts_lymphocytes"
## [62] "gender"
## [63] "sIgG4_egg"
## [64] "peanuts_mother_exposure_regularity"
## [65] "cowmilkformula_exposure_regularity"
## [66] "anti_helminthic_exposure_age"
## [67] "anti_helminthic_exposure_yearly"
## [68] "sIgE_milk"
## [69] "sIgE_wheat"
## [70] "cowmilkformula_exposure_age_first"
##
## $Clinical$value
##                                               value.var
## farmanimal_contact_mother                  -0.373672881
## farmanimal_contact_child                   -0.368285038
## fuel_cooking_Electricity_Gas                0.299133882
## fuel_cooking_Open_fires_outside            -0.292248108
## sunlight_exp_winter                        -0.255020664
## sunlight_exp_summer                        -0.251980621
## fuel_cooking_Paraffin_Stove                -0.233614218
## fuel_heating_Wood_Coal                     -0.229311314
## pets_dog                                   -0.191435541
## peanuts_exposure                            0.161328971
## delivery_mode                               0.147419463
## log_blood_counts_monocytes                 -0.124912412
## henegg_exposure_regularity                 -0.124618884
## fuel_cooking_heating_Paraffin               0.111561451
## pets_cat                                   -0.108269492
## fuel_heating_Electricity                    0.104746352
## household_peoplelivingtogether             -0.101538841
## familyhistory_mother_other_allergic_disease  0.097907550
## hayfever_ever                               0.096488392
## total_IgE                                   0.095380497
## sIgG4_fish                                  0.093057765
## wheat_exposure_regularity                   0.088287925
## sIgE_Der                                    0.086370742
## cigarettesmoke_numberofpersons              0.082991702
## household_otheryoungerchildren             -0.077674077
## household_otherolderchildren               -0.076385133
## paracetamol_exposure_time                   0.070698858
## sIgE_fish                                   0.070524792
```

```
## log_blood_counts_plts                              0.068026098
## sIgE_hazelnut                                      0.067942316
## anti_helminthic_exposure                           0.067237724
## sIgE_peanut                                        0.063126771
## sIgG4_casein                                       0.060997082
## solidfood_age_introduction                        -0.060196960
## sIgE_CCD                                           0.059849293
## paracetamol_exposure_age                           0.057468967
## othernuts_exposure                                 0.056944256
## sIgE_soy                                           0.056397305
## log_blood_counts_eosinophils                      -0.055162546
## sIgG4_Der                                          0.052318765
## antibiotic_exposure_last2months                    0.050410833
## fish_exposure                                      0.050317589
## enrolment_age                                      0.050144845
## henegg_exposure_age_first                          0.046491990
## breastfeeding_ever                                -0.045003432
## peanuts_mother_exposure_avoidance                 -0.044852863
## sIgE_egg                                           0.041245781
## dairyproducts_exposure_age_first                   0.038952104
## fuel_heating_Other                                -0.038003622
## sIgG4_peanut                                       0.037453848
## immun_number                                      -0.035727417
## anti_helminthic_exposure_last                      0.034400670
## cowmilkformula_exposure                            0.033572465
## antibiotic_exposure                               -0.033407737
## sIgG4_hazelnut                                     0.030305701
## familyhistory_father_other_allergic_disease        0.028850813
## henegg_exposure                                   -0.025409383
## formula_age_introduction                          -0.022148658
## breastfeeding_uptoage_age_complete                -0.020160231
## log_blood_counts_hb                               -0.014051939
## log_blood_counts_lymphocytes                       0.010774246
## gender                                             0.008548880
## sIgG4_egg                                         -0.007594799
## peanuts_mother_exposure_regularity                -0.005029687
## cowmilkformula_exposure_regularity                -0.002925952
## anti_helminthic_exposure_age                       0.002908559
## anti_helminthic_exposure_yearly                   -0.002277732
## sIgE_milk                                         -0.001849621
## sIgE_wheat                                        -0.001509870
## cowmilkformula_exposure_age_first                  0.001485494
##
##
## $comp
## [1] 1
```

```
selectVar(diablo.sosall_full, block = 'Cytokine', comp = 1)
```

```
## $Cytokine
## $Cytokine$name
##  [1] "VEGF.D"     "Tie.2"      "PlGF"       "IL.22"      "bFGF"
##  [6] "MIP.1beta"  "MIP.1alpha" "ICAM.1"     "VCAM.1"     "IL.6"
## [11] "Flt.1"      "IL.8"       "IL.12p70"   "IL.15"      "IL.1beta"
##
## $Cytokine$value
##              value.var
## VEGF.D     -0.62382738
## Tie.2      -0.38259064
## PlGF        0.38183716
## IL.22       0.31300121
## bFGF        0.23966738
## MIP.1beta  -0.19087976
## MIP.1alpha -0.17873182
## ICAM.1     -0.16246671
## VCAM.1     -0.14373614
## IL.6       -0.12707211
## Flt.1       0.09866342
## IL.8       -0.08367894
## IL.12p70    0.07996811
## IL.15       0.06995760
## IL.1beta    0.06037711
##
##
## $comp
## [1] 1
```

Explain the plots here.

##### 3.3.2 Visualization of DIABLO Components

The plotDiablo function visualizes the relationships between the components and the datasets, offering a graphical representation of how each data type contributes to the model. This visualization aids in interpreting the model’s structure and the interplay between different data sources.

```r
plotDiablo(diablo.sosall_full, ncomp = 1)
```

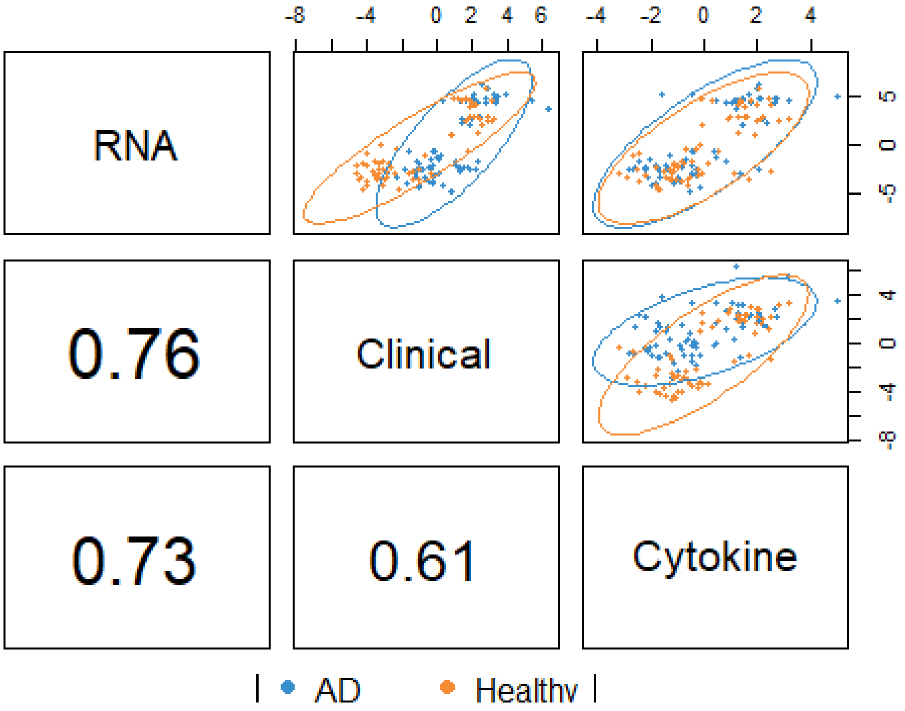


##### 3.3.3 Individual and Variable Plots

Further visualizations, including plots of individuals (plotIndiv) and variables (plotVar), provide a deeper dive into the model's behavior. These plots reveal patterns, groupings, and outliers within the integrated analysis, offering valuable insights into the data's underlying structure.

```
plotIndiv(diablo.sosall_full,
          ind.names = FALSE,
          study = 'global',
          style = 'ggplot2',
          legend = TRUE,
          centroid = TRUE,
          star = TRUE,
          title = 'SOSALL, DIABLO comp 1-2',
          block = 'weighted.average',
          ellipse = FALSE,
          ellipse.level = 0.5)
```

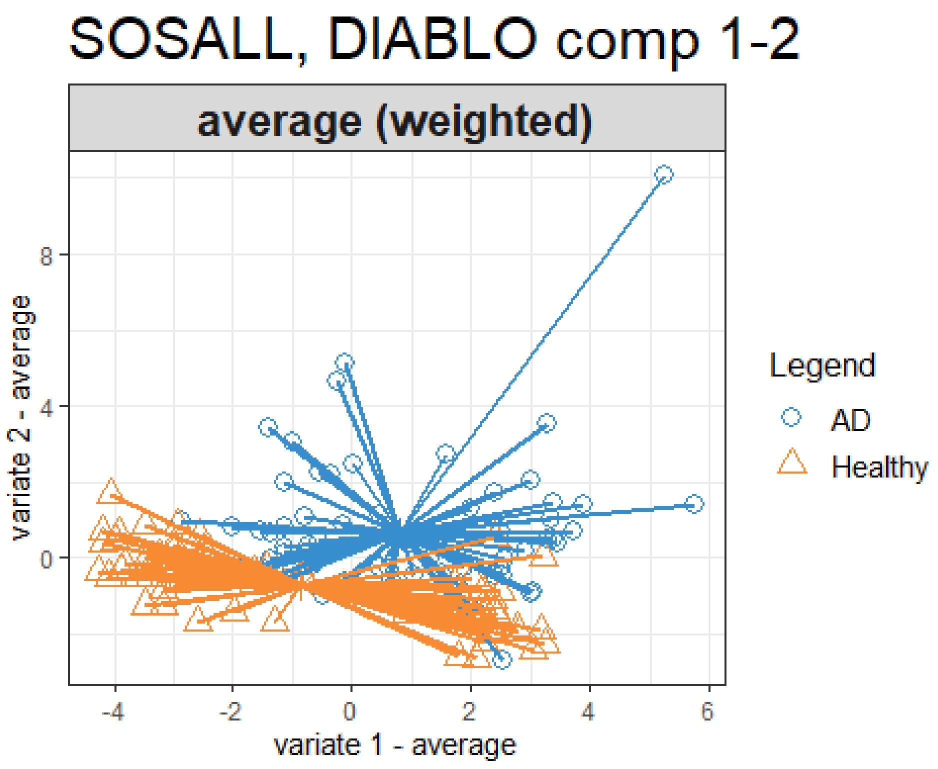


```
plotVar(diablo.sosall_full,
        var.names = FALSE,
        style = 'ggplot2',
        legend = TRUE,
        pch = c(15, 16, 17),
        cex = c(3,3,3),
        cutoff = 0.3,
        col = c('#D64933', 'orange','#0C7C59'),
        title = 'SOSALL, DIABLO comp 1 - 2')
```

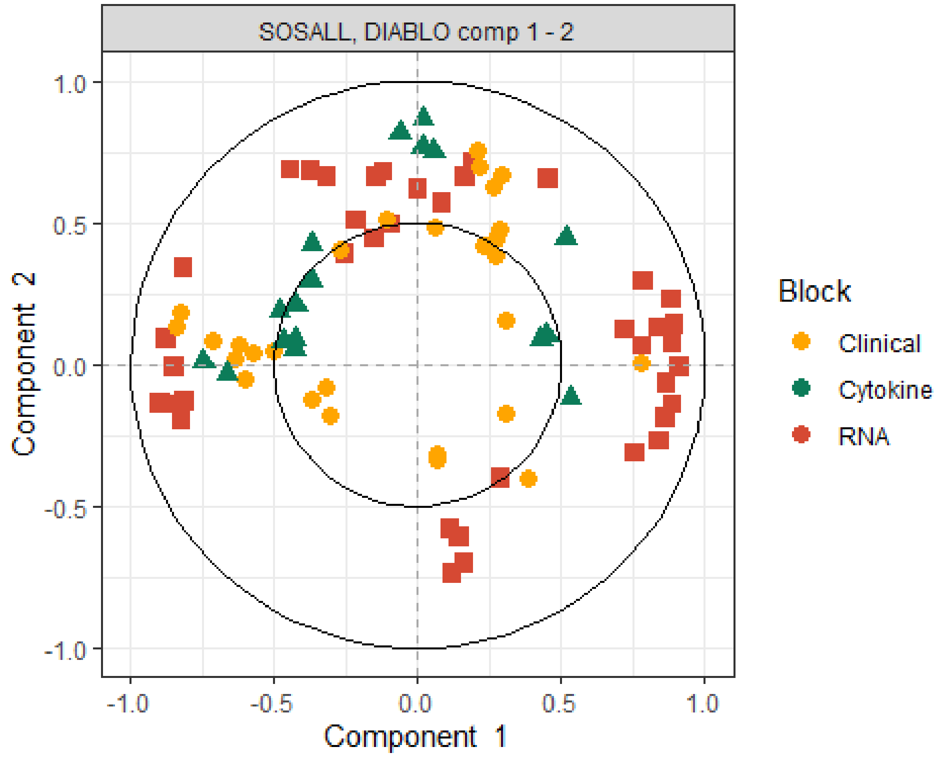


##### 3.3.4 Loadings Analysis

Finally, examining the loadings for components across datasets highlights the variables with the most significant contributions to each component. This analysis is key to understanding the model's focus and can guide further biological interpretation and validation efforts.

Throughout this process, your use of DIABLO in R Markdown not only facilitates a comprehensive analysis of complex multi-omics data but also ensures that the results are reproducible and well-documented, which is paramount in scientific research.

```r
plotLoadings(diablo.sosall_full,  comp = 1, block = 'RNA',contrib = 'max',
method = 'median', ndisplay = 20, plot = TRUE)
```

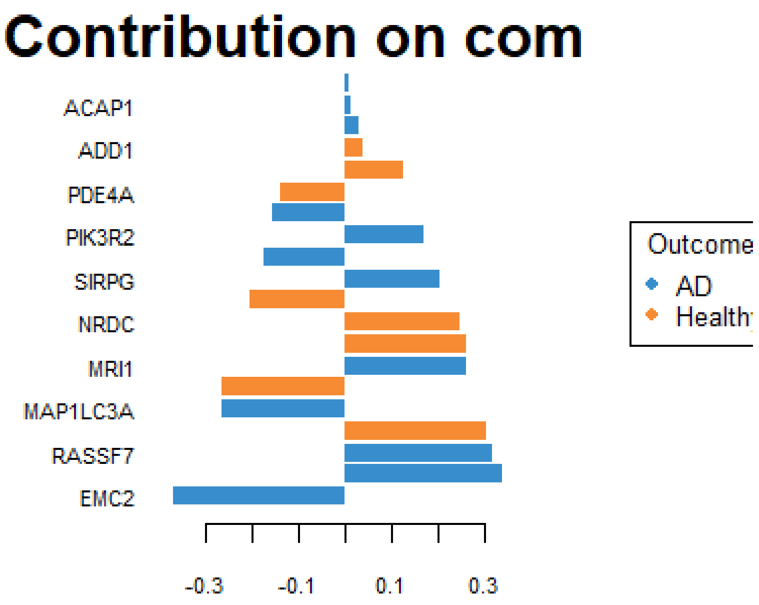


```r
plotLoadings(diablo.sosall_full,  comp = 2, block = 'RNA',contrib = 'max',
method = 'median', ndisplay = 20, plot = TRUE)
```

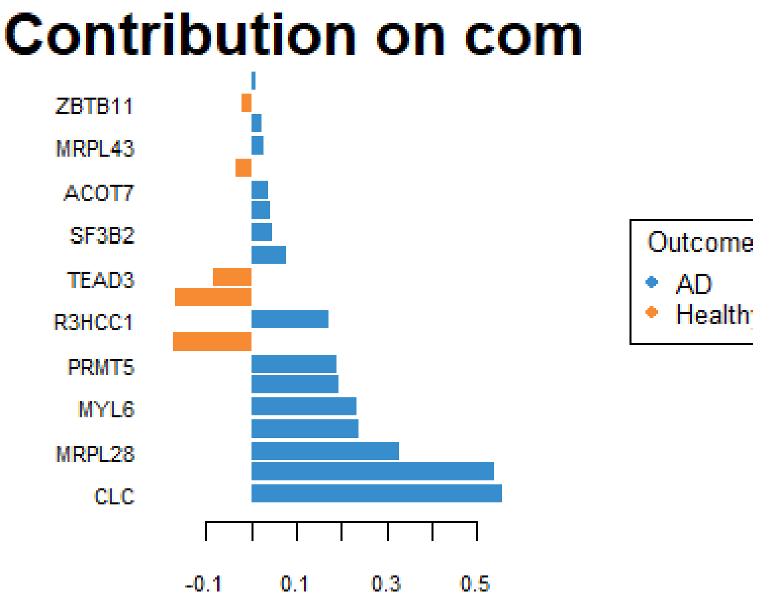

```r
plotLoadings(diablo.sosall_full,  comp = 1, block = 'Clinical',contrib =
'max', method = 'median', ndisplay = 20, plot = TRUE)
```

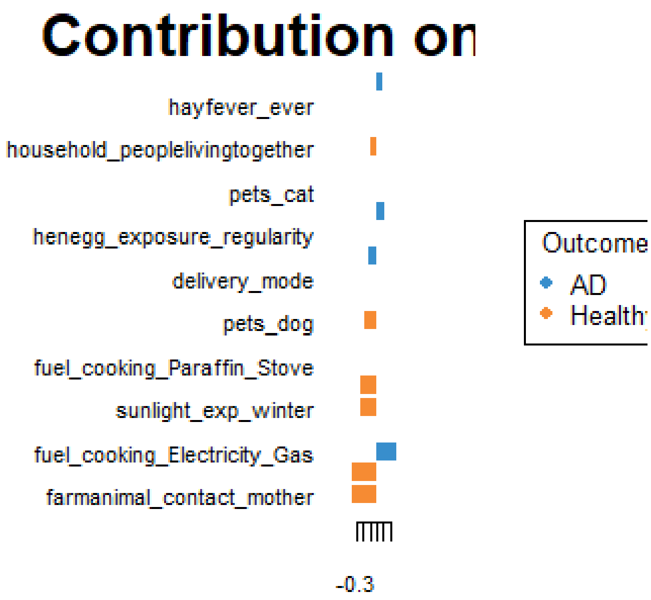


```r
plotLoadings(diablo.sosall_full,  comp = 2, block = 'Clinical',contrib =
'max', method = 'median', ndisplay = 20, plot = TRUE)
```

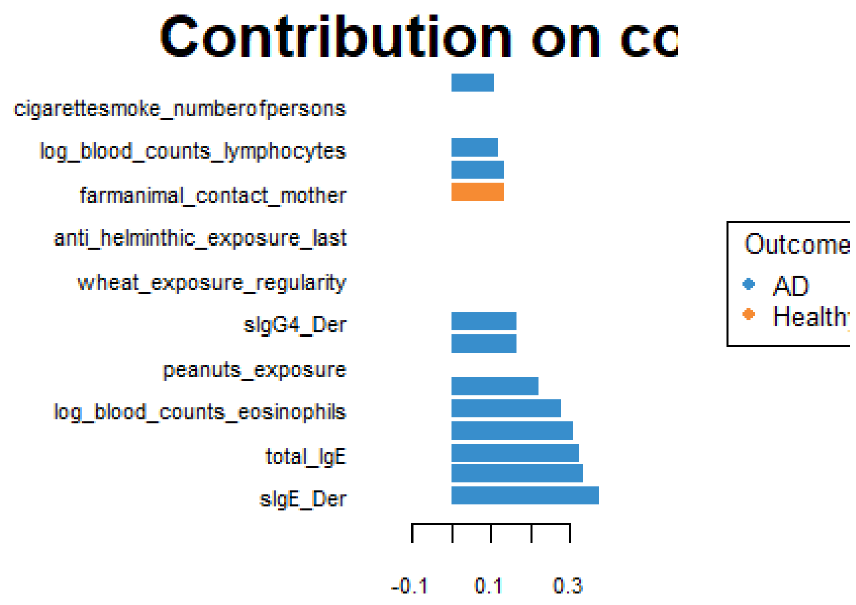


```
plotLoadings(diablo.sosall_full,  comp = 1, block = 'Cytokine',contrib =
'max', method = 'median', ndisplay = 20, plot = TRUE)

## 'ndisplay' value is larger than the number of selected variables! It has
been reseted to 15 for block Cytokine
```

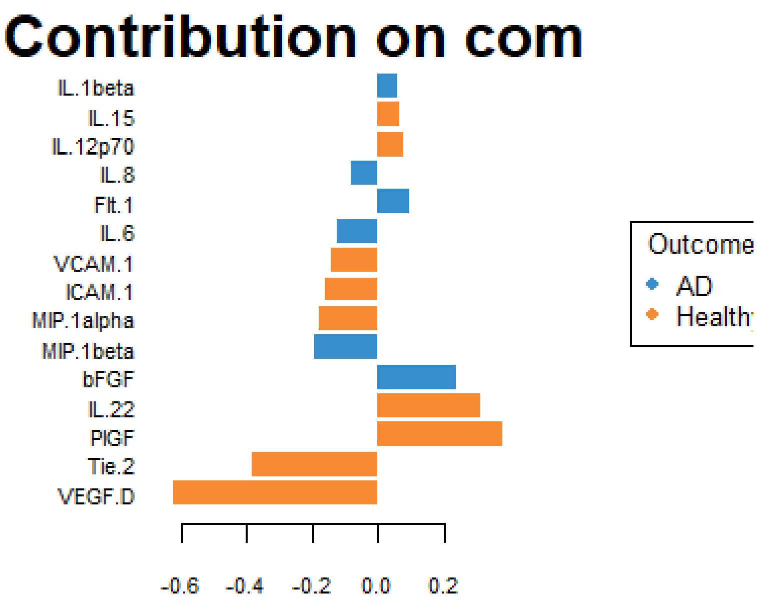


```
plotLoadings(diablo.sosall_full,  comp = 2, block = 'Cytokine',contrib =
'max', method = 'median', ndisplay = 20, plot = TRUE)

## 'ndisplay' value is larger than the number of selected variables! It has
been reseted to 5 for block Cytokine
```

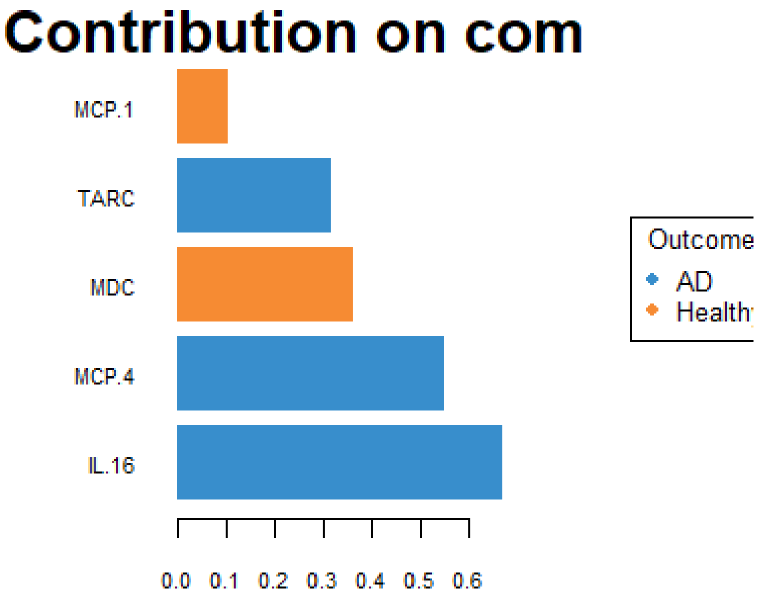

Contribution on com
MCP.1
TARC
MDC
MCP.4
IL.16
0.0 0.1 0.2 0.3 0.4 0.5 0.6
Outcome
AD
Health
